# Design and Analysis Heterogeneity in Observational Studies of COVID-19 Booster Effectiveness: A Review and Case Study

**DOI:** 10.1101/2023.06.22.23291692

**Authors:** Sabir Meah, Xu Shi, Lars G. Fritsche, Maxwell Salvatore, Abram Wagner, Emily T. Martin, Bhramar Mukherjeea

**Author notes:** Correspondence to: Bhramar Mukherjee, PhD, Department of Biostatistics, University of Michigan School of Public Health 1415 Washington Heights Ann Arbor, MI 48109.

## Abstract

**Background:** Observational vaccine effectiveness (VE) studies based on real-world data are a crucial supplement to initial randomized clinical trials of Coronavirus Disease 2019 (COVID-19) vaccines. However, there exists substantial heterogeneity in study designs and statistical methods for estimating VE. The impact of such heterogeneity on VE estimates is not clear.

**Methods:** We conducted a two-step literature review of booster VE: a literature search for first or second monovalent boosters on January 1, 2023, and a rapid search for bivalent boosters on March 28, 2023. For each study identified, study design, methods, and VE estimates for infection, hospitalization, and/or death were extracted and summarized via forest plots. We then applied methods identified in the literature to a single dataset from Michigan Medicine (MM), providing a comparison of the impact of different statistical methodologies on the same dataset.

**Results:** We identified 53 studies estimating VE of the first booster, 16 for the second booster. Of these studies, 2 were case-control, 17 were test-negative, and 50 were cohort studies. Together, they included nearly 130 million people worldwide. VE for all outcomes was very high (around 90%) in earlier studies (i.e., in 2021), but became attenuated and more heterogeneous over time (around 40%-50% for infection, 60%-90% for hospitalization, and 50%-90% for death). VE compared to the previous dose was lower for the second booster (10-30% for infection, 30-60% against hospitalization, and 50-90% against death). We also identified 11 bivalent booster studies including over 20 million people. Early studies of the bivalent booster showed increased effectiveness compared to the monovalent booster (VE around 50-80% for hospitalization and death).

Our primary analysis with MM data using a cohort design included 186,495 individuals overall (including 153,811 boosted and 32,684 with only a primary series vaccination), and a secondary test-negative design included 65,992 individuals tested for SARS-CoV-2. When different statistical designs and methods were applied to MM data, VE estimates for hospitalization and death were robust to analytic choices, with test-negative designs leading to narrower confidence intervals. Adjusting either for the propensity of getting boosted or directly adjusting for covariates reduced the heterogeneity across VE estimates for the infection outcome.

**Conclusion:** While the advantage of the second monovalent booster is not obvious from the literature review, the first monovalent booster and the bivalent booster appear to offer strong protection against severe COVID-19. Based on both the literature view and data analysis, VE analyses with a severe disease outcome (hospitalization, ICU admission, or death) appear to be more robust to design and analytic choices than an infection endpoint. Test-negative designs can extend to severe disease outcomes and may offer advantages in statistical efficiency when used properly.

## Introduction

The first emergency use authorization (EUA) of vaccines for Coronavirus Disease 2019 (COVID-19) in the US in December 2020 ushered in a new era of the pandemic, with mRNA vaccines having shown strong efficacy against both confirmed infection and severe outcomes in randomized clinical trials.^1^ After the EUA and broad administration of the vaccines, observational studies sought to measure real-world vaccine effectiveness (VE) against infection and severe disease outcomes such as hospitalization and death. Many early observational studies reported strong effectiveness against both infection and severe disease, consistent with clinical trials.^2^

However, in late 2021, early data on waning immunity motivated officials in the US and other countries to approve booster doses of COVID-19 vaccines.^3–5^ The landscape has further evolved with the approval and administration of additional booster doses,^6^ emergence of new and more immune-evasive severe acute respiratory syndrome coronavirus 2 (SARS-CoV-2) variants,^6–8^ bivalent boosters updated for such new variants,^9–11^ and general increase in vaccine availability worldwide.^12, 13^ In the US as of May 3, 2023, 81.4% of the population has received at least one dose of a COVID-19 vaccine, while 16.9% of the population has received a new bivalent booster.^14^ This dynamically evolving environment of vaccine-conferred immunity has led to an exploding number of papers and preprints on VE in 2022. Our sweep of all VE literature identified 380 papers and preprints in 2022, compared to 77 in 2021.

However, reported VEs for the first booster versus primary series vaccination in such studies have varied immensely, particularly for infection where estimates from as high as 93% to as low as negative effectiveness can be found.^15–17^ Differing epidemiological circumstances, including pre-existing natural immunity and contemporary variant circulation, may explain some of the differences between study results, but there still remains substantial heterogeneity in VE estimates reported by studies in the same country at around the same time, particularly in the US.^18, 19^.

Another factor that may explain some of the substantial differences in the results of VE studies is heterogeneity in study design and statistical methods. In a systematic review and meta-analysis of VE studies for primary series vaccination, Zeng et al. noted that “there is high heterogeneity between studies, and high statistical heterogeneity is also observed in most analysis.”^20^ The same has been observed in booster VE studies regarding both study design and statistical methods. For example, in Figure 1, we see in the latter half of 2022, of the 177 papers, 49 (28%) used a test-negative design, 118 (67%) used a cohort design, and 10 (6%) used a case-control design. In addition to differences in study design, studies made different choices regarding adjustment for confounding, stratification, and other aspects of statistical analysis. The effect of these choices on the ultimate VE estimates for infection, hospitalization, and death is not clear.

**Figure 1:**
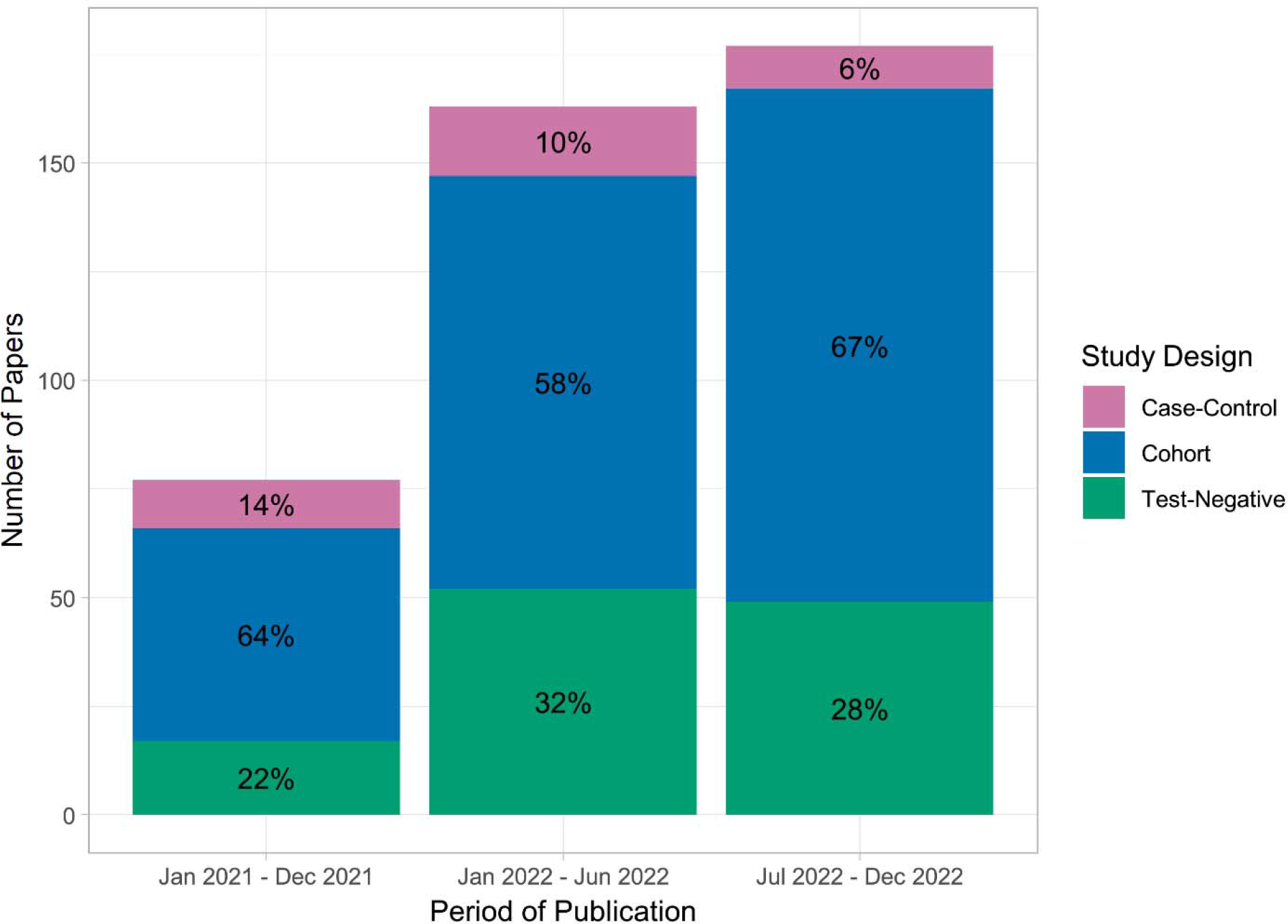
Number of published COVID-19 VE articles, preprints, or conference abstracts in each of 3 time periods: 2021 (January 2021 – December 2021), first half of 2022 (January 2022 – June 2022), and second half of 2022 (July 2022 – December 2022). Bars representing each time period are colored by class of study design, with the percentage of literature having each design in that time period indicated by the percentages given inside each shaded area.

In this study we attempt to analyze study design and analysis heterogeneity for COVID-19 booster VE studies using a two-step process. First, we conduct a literature review of studies estimating booster VE relative to primary series vaccination or second booster VE relative to first booster vaccination, summarizing VE estimates and methodology found in current work. We also report VE estimates from early studies of the bivalent booster. Second, we implement several different methods found in the literature on electronic health record (EHR) data from Michigan Medicine (MM), demonstrating how study design and analysis decisions can influence reported VE estimates and results within the same dataset.

## Methods

### Literature Review

#### Database Search

We performed a search in the PubMed and Embase databases on January 1, 2023, for published articles or preprints written in English after January 1, 2021, that provided a VE estimate of the first booster versus primary series, or second booster versus first booster, against infection, hospitalization, ICU admission, or death. The search used the following 4 groups of terms, requiring chosen articles to include a term from each group in their title and/or abstract: (1) terms for COVID-19 generally, (2) terms for indicating either the first or second booster, (3) terms for a VE metric, and (4) terms indicating real-life outcomes of interest (infection, hospitalization, ICU admission, and death). For detailed information on the search strategy, including specific verbiage for term blocks, see Supplementary Methods A1 and A2.

#### Article Selection Process

Database search results, along with several additional articles found from Google Scholar, were screened for potential duplicates by Rayyan (a web tool for literature and systematic reviews) and then removed if confirmed as a duplicate by manual title and abstract review.^21^ The remaining articles were subjected to a two-step review process of a title and abstract review followed by a full text review. As inclusion criteria, we required included articles to (1) study a vaccine approved for COVID-19 (in any country); (2) generate a VE metric (a proportion/percentage, odds ratio, risk ratio, or hazard ratio) against infection, hospitalization, ICU admission, and/or death; (3) generate the VE metric for the first booster (first dose after a primary series of vaccination, including both 1 and 2 dose regimens) compared to the primary series, or the second booster compared to the first booster (sometimes termed relative VE, or rVE, by papers). The first step of the title and abstract review was conducted to select articles that met the inclusion criteria in items (1) and (2), while the second step of the full text review selected articles that met final inclusion criteria in item (3). We also constructed a PRISMA diagram describing this process.^22^

We additionally conducted a rapid review of bivalent boosters (which were first approved in the US in August 2022) on March 28, 2023, by searching “covid-19 ‘bivalent’ booster ‘vaccine effectiveness’” in Google Scholar. We selected articles and CDC *Morbidity and Mortality Weekly Report* releases that generated bivalent booster (targeting either the BA.1 or BA.4/5 variants) VE estimates based on the title and information shown in the preview on the search page. Unlike the main search for monovalent boosters, we did not enforce the requirement of VE being calculated compared to a certain number of previous boosters, as many studies now calculate bivalent booster VE in comparison with any number of previous monovalent boosters.

#### Data Collection and Presentation

The study period, country of the study, general study design, dose and type of vaccine, and VE estimates (against infection, hospitalization, severe disease, and/or death for dose comparisons of interest) were collected in the full-text review of each included study. Study design was classified as one of test-negative, matched test-negative, case-control, cohort, or matched cohort. With the level of heterogeneity in vaccine brands used, study design, statistical methods, definitions of outcomes, and setting within our inclusion criteria, we did not perform any pooling of results or meta-analysis, with an emphasis on narrative synthesis instead. We simply visualized aggregate results by plotting VE estimates from the studies by using forest plots stratified by booster number (1st or 2nd), outcome (infection or hospitalization/severe disease or death), and time since vaccination (including a category for studies that did not stratify on time since vaccination). Studies in forest plots were displayed in (ascending) chronological order of last month in the study period, with publication date of the study used to break ties. For the bivalent booster forest plot, when the source studies were stratified by miscellaneous factors, we plotted results corresponding to only the oldest age, most recent variant, longest time since previous dose, and a 2-month time since boosted strata.

Forest plots were created in R, version 4.2.2.^23^ Aggregated data files collected in the literature review and code used to create plots can be found at https://github.com/smeah/COVID19BoosterVE.

### Michigan Medicine Data Analysis

#### Study Population

We implemented both cohort and test-negative designs with two different study populations. Study design and statistical methods for the test-negative analysis can be found in Supplementary Methods A3. Our cohort design study population for the main analysis consisted of 186,495 individuals after exclusion criteria (from 272,399 before exclusion criteria) who received two or three doses of the BNT162b2 (Pfizer) or mRNA-1273 (Moderna) vaccines and had at least two EHR entries at MM between October 1, 2021, and December 31, 2022. Individuals were eligible to contribute person-time from the start of the study period to their last EHR entry or the end of the study period (December 31, 2022), whichever occurred first. Any individuals with a documented infection prior to the start of the study period or missing values of any covariates were excluded from the analysis. We present analysis results for those who were boosted before or during October – December 2021 in the main text due to more complete information with longer follow-up data. Results for all other subsequent quarters are presented in the supplementary appendix (Figures A8-A9, A11, and A13 for the main study population).

#### Exposures

We considered three different exposure definitions (depending on booster dose) in this study in separate models. In our main analysis, we considered as exposure exactly three doses of the BNT162b2 or mRNA-1273 vaccines, and as reference two doses of BNT162b2 or mRNA-1273, both restricted to only the first 120 days of person-time, with individuals being censored after. We included both homologous and heterologous monovalent vaccine regimens, provided all doses were either BNT162b2 or mRNA-1273 (we did not include bivalent boosters). As secondary analyses, to understand the sequential benefit of serial boosting, we considered as exposure three or more doses of BNT162b2 or mRNA-1273 and four or more doses, with the reference groups being two doses and three doses, respectively.

#### Outcomes

We considered COVID-19 infection and severe COVID-19 disease as outcomes in different models. Infection was defined as a COVID-19 diagnosis or positive PCR test for SARS-CoV-2. Severe COVID-19 disease was defined as hospitalization or an ICU admission within 30 days of a positive PCR test or death within 60 days.

#### Statistical Analysis

We used Cox regression to model the association between each combination of outcome and exposure in our analysis.^24–26^ VE was calculated by exponentiating the coefficient for vaccination, giving the hazard ratio for the outcome for those boosted compared to those in the previous stage of vaccination, then subtracting it from one, i.e., 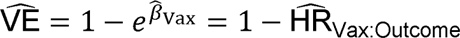 (see Supplementary Methods A4 for definitions of vaccination and outcome indicators). Aside from the outcome event, individuals were censored at the end of the study period, time of their last EHR entry (if before the end of the study period), death for any reason, and a positive PCR test (when infection was not the outcome event of interest).

We compared covariate adjustment, matching, and propensity score methods in the analysis. The (same) set of covariates was used for adjustment, matching, and propensity score generation: age, sex, BMI, race/ethnicity (non-Hispanic white, non-Hispanic black, other, and unknown), neighborhood development index (NDI),^27, 28^ population density (quartiles), healthcare worker status, whether the subject was receiving primary care at Michigan Medicine or not, Charlson Comorbidity Index (categories of 0, 1-2, 3-4, and >= 5),^29^ number of documented negative SARS-CoV-2 PCR tests from the subject at any time (categories of 0, 1, 2-3, 4-5, 6-10, >= 11). When used in exact matching, age was additionally categorized into three bins (18-49, 50-65, and >= 65), and BMI was categorized into four bins (underweight, healthy weight, overweight, and obese).

Matching and propensity score methods included exact matching, propensity score matching, regression adjustment for propensity score, and inverse probability weighting on propensity score (IPW). We calculated separate propensity scores for both receiving a booster and reporting a SARS-CoV-2 PCR test at any time. We implemented both unstratified and stratified Cox proportional hazard models for matched data. For propensity score methods, we performed analysis with and without regression adjustment of the covariate set. Supplementary Table A1 summarizes each of these methods and provides associated model statements.

All analyses were conducted in R, version 4.2.2.^23^ Code examples and code used for analysis can be found at https://github.com/smeah/COVID19BoosterVE. Data cannot be shared publicly due to patient confidentiality. The data underlying the results presented in the study are available from the University of Michigan Precision Health Analytics Platform at https://precisionhealth.umich.edu/tools-resources/data-access-tools/ for researchers who meet the criteria for access to confidential data.

## Results

### Literature Review

#### Included Studies

324 records were identified through the database search and 69 satisfied our inclusion criteria, with 53 of them estimating VE for the first booster, and 16 for the second booster. Figure A1 displays the PRISMA flow diagram describing the process of article selection. Tables A2-A4 present summaries of the types of study designs adopted in the included studies. Out of the 69 included studies, 17 used a test-negative design (4 with matching), 2 a non-test-negative case-control design (none with matching), and 50 used a cohort design (19 with matching). The proportion of cohort designs was higher for second booster studies compared to first booster studies (81% versus 70%, respectively). The 69 studies in the literature review included 129,368,483 individuals (or tests in designs where tests, not individuals, were the unit of observation) in total.

#### First Booster VE Against Infection

A forest plot summarizing first booster versus primary series VE against infection for studies not stratified on time since booster can be found in Figure 2, while VE stratified by 2 weeks, 1 month, 2 months, and 3 months since booster administration can be found in Figures A2-A3.

**Figure 2:**
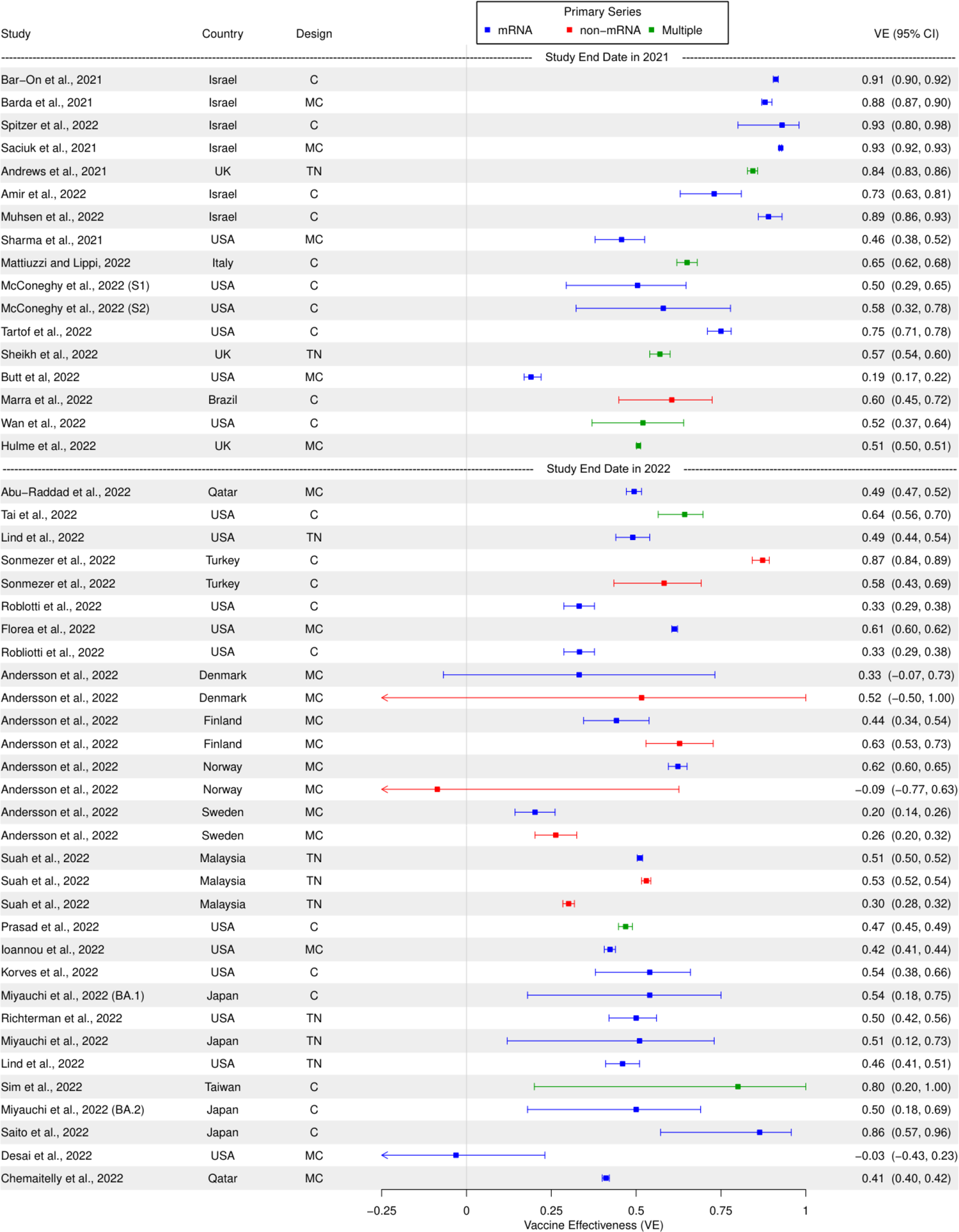
First booster VE compared to primary series against infection, in studies unstratified by time since booster administration. Studies are sorted in chronological order of last month included in the study period (with publication date being used to break ties). Year of study end date (noting that the end of 2021 was a time approximately corresponding to the dominance of the Omicron variant in many countries) is denoted by dashed lines in the figure, with studies completed after the year start below the respective year marker. Abbreviations: TN, test-negative; MTN, matched test-negative; CC, case-control (non-test-negative); MCC, matched case-control (non-test-negative); C, cohort; MC, matched cohort. S1/S2: System number for McConeghy et al.^48^

Infection VE for the first booster was high (80%-95%) for the earliest published studies in 2021, but then waned by the start of 2022 to around 50% (Figure 2). Similar patterns were noted in time-stratified studies (Figures A2-A3). In such studies, a clear pattern of waning immunity can be observed. Infection VE one month after vaccination ranged from 41%-59% in 2022 studies, then (except for one outlier reporting a VE of 69%) fell to −4%-44% (with most studies around 30%) three months after infection.

Cohort designs, including both with and without matching, were the majority class of study design (Figure 2). Test-negative designs however appeared to be popular for earlier studies stratifying on time since vaccination (Figures A2-A3). The earliest studies were predominantly from Israel, followed by studies in the US and Western Europe (particularly the UK), then followed by studies from Asia (although the US contributed many later studies as well). Perhaps related to the timing of studies, Israeli studies for the most part exhibited similar VE, while studies from the US or Europe displayed a high degree of heterogeneity of VE estimates.

There were no clear differences in booster VE when comparing study populations with mRNA or non-mRNA primary series vaccines (Figure 2). However, studies comparing the first booster to a non-mRNA primary series vaccination sometimes exhibited slightly lower VE compared to studies using an mRNA primary series, when from around the same time and when stratified by time since vaccination (Figures A2-A3).

#### First Booster VE Against Hospitalization/Severe Disease or Death

A forest plot summarizing first booster versus primary series VE against hospitalization or severe disease and against death can be found in Figure 3, while VE stratified by 1 month, 2 months, and 3 months since booster administration can be found in Figure A4 for hospitalization/severe disease and Figure A5 for death.

**Figure 3:**
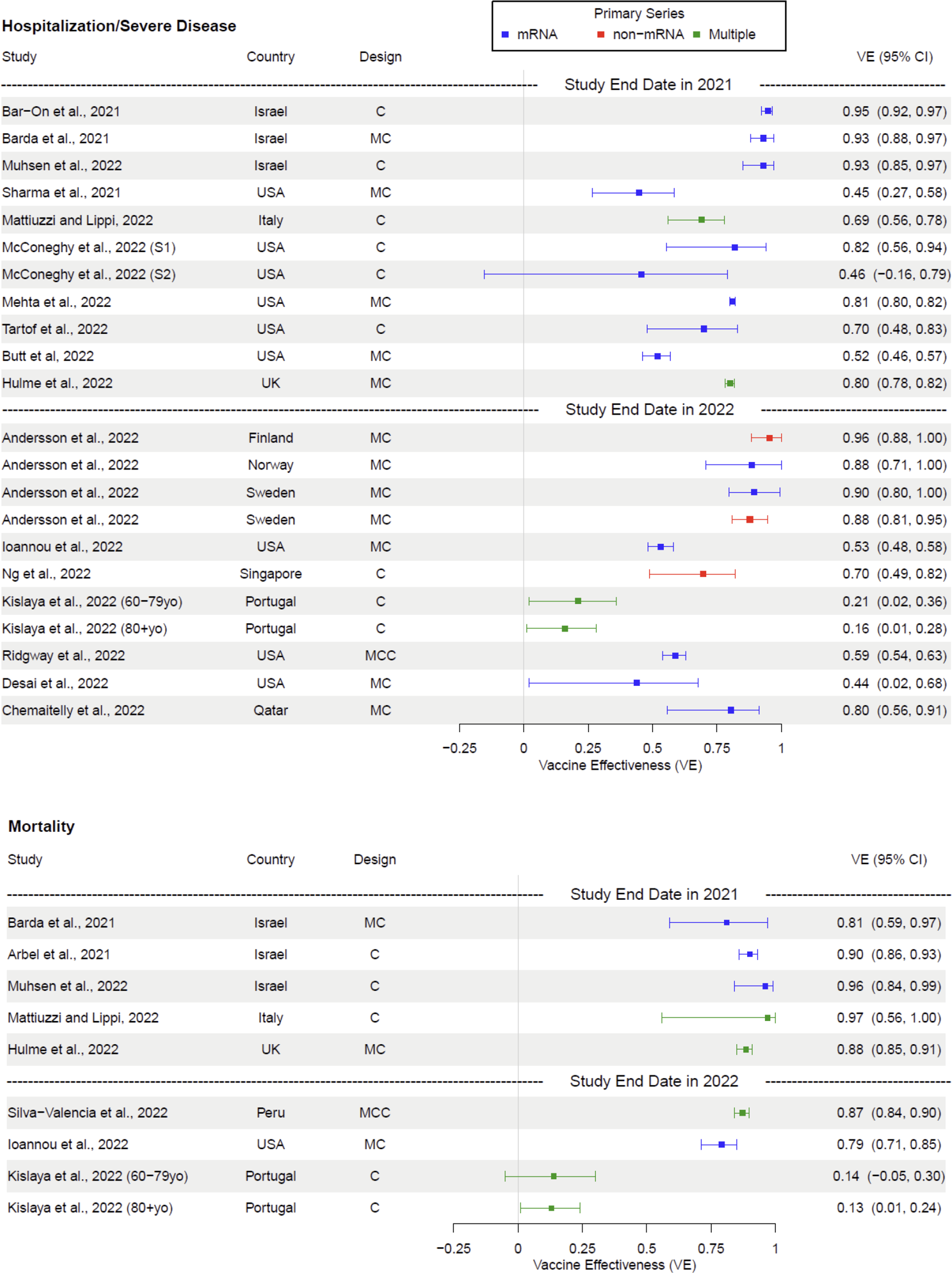
First booster VE compared to primary series against hospitalization or severe disease outcomes and mortality in separate panels, in studies unstratified by time since booster administration. Studies are sorted in chronological order of last month included in the study period (with publication date being used to break ties). Year of study end date (noting that the end of 2021 was a time approximately corresponding to the dominance of the Omicron variant in many countries) is denoted by dashed lines in the figure, with studies completed after the year start below the respective year marker. Abbreviations: TN, test-negative; MTN, matched test-negative; CC, case-control (non-test-negative); MCC, matched case-control (non-test-negative); C, cohort; MC, matched cohort.

Similar to infection VE, early studies examining VE against hospitalization or severe disease outcomes exhibited very high VE (81%-97%), particularly the earliest Israeli studies (Figures 3, A4). However, the drop in VE in later studies was not nearly as precipitous (with just Kislaya et al.^30^ outlying), declining to around 44%-80%. It also occurred later than with infection, in mid-2022 (Figures 3, A4). Similarly, VE against mortality was high in early studies (80%-90%) and dropped only slightly (again apart from Kislaya et al.^30^) in later studies to 62%-89% (Figures 3, A5).

When comparing studies stratified on time since vaccination, the effect of waning immunity for the outcomes of severe disease and death over 3 months since vaccination was much less than for infection. VE against hospitalization ranged from 64%-91% one month after vaccination in 2022 studies and from 48%-79% three months after vaccination in studies also ending in 2022.

The cohort study design was far more dominant in hospitalization/severe disease or mortality VE studies than infection studies (Figure 3). There were only two mortality studies that used a non-cohort design: one case-control and one test-negative (Figures 3, A5).

Like infection VE, hospitalization/severe disease studies did not display a clear divide in VE estimates between mRNA and non-mRNA primary series. For mortality outcomes, many studies used a mixed selection of primary series vaccinations, so it was impossible to disentangle differences in VE across these subgroups.

#### Second Booster VE

Forest plots summarizing second booster versus first booster VE against hospitalization/severe disease and mortality for studies can be found in Figure 4. An additional plot for second booster VE against infection can be found in Figure A6. Forest plots summarizing second booster VE stratified by time since vaccination are not included on account of the small number of studies for each time strata.

**Figure 4:**
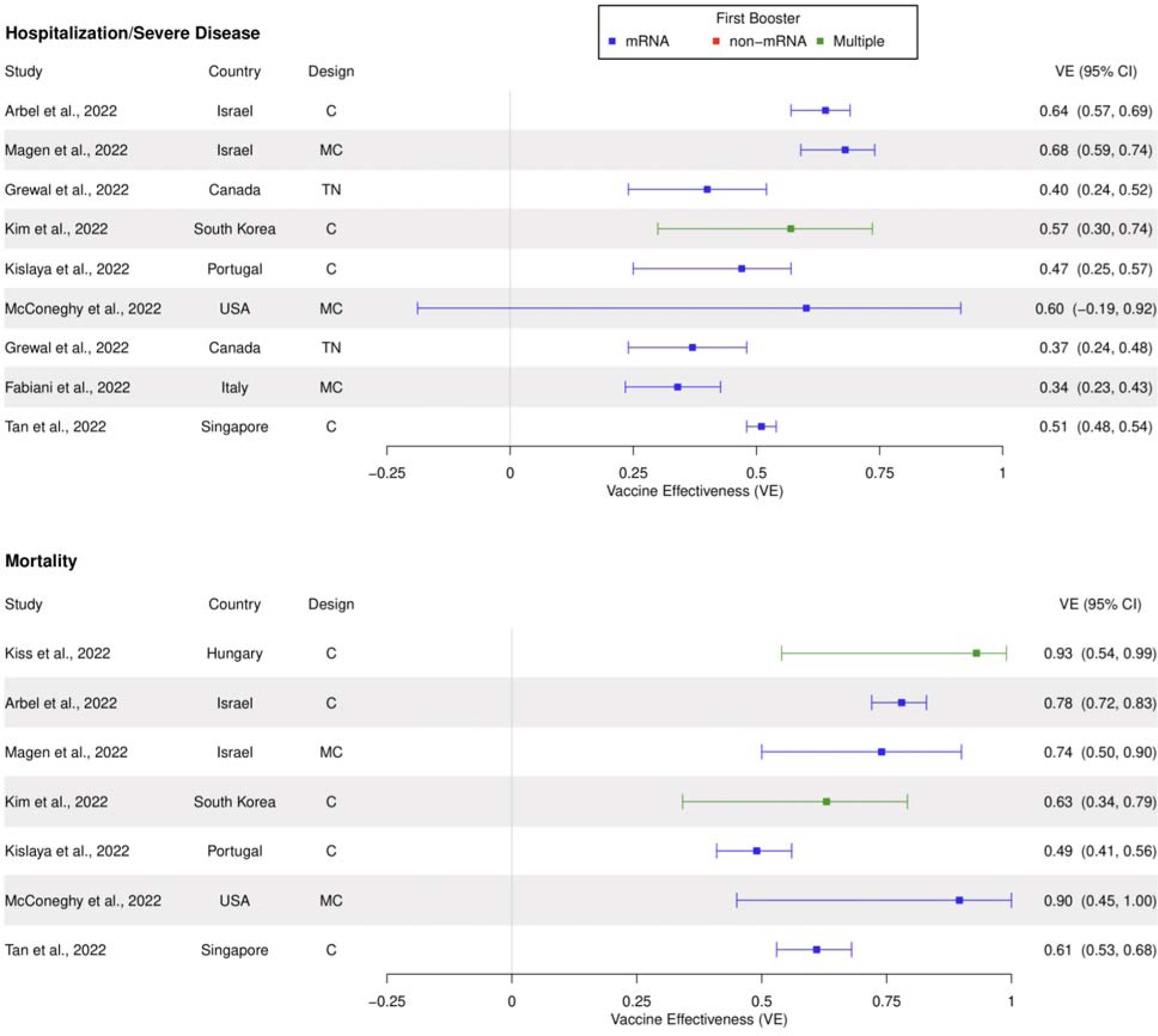
Second booster VE compared to primary series against hospitalization or severe disease outcomes and mortality in separate panels, in studies unstratified by time since booster administration. Studies are sorted in chronological order of last month included in the study period (with publication date being used to break ties). Abbreviations: TN, test-negative; MTN, matched test-negative; CC, case-control (non-test-negative); MCC, matched case-control (non-test-negative); C, cohort; MC, matched cohort.

Similar to first booster results, second booster versus first booster VE was higher for the early set of studies (30%-61% for infection, 40-68% for hospitalization/severe disease, 74%-93% for death), often in Israel, and dropped for later studies in other settings (Figures 4, A6). But overall, VE estimates across the board appeared to be lower for the second booster. The decline through later study dates was still more pronounced for infection than for hospitalization/severe disease and mortality. Infection VE in later studies fell to 14%-31% (Figure A6). VE against hospitalization after the early Israeli studies was in the range of 34%-60% (Figure 4). VE against death ranged from 49%-90%, with more later studies being closer to the lower bound (Figure 4).

Cohort designs were dominant for second booster studies. Every second booster mortality study in the review used a cohort design, and the test-negative design was used by just Grewal et al. for infection and severe disease outcomes.^31, 32^ Most second booster studies estimating infection VE used matching, though several hospitalization/severe disease studies and most mortality designs did not use matching.

There were no easily discernible patterns regarding first booster type (mRNA versus non-mRNA).

#### Bivalent Booster VE

Our Google Scholar query for bivalent booster VE returned 853 articles and we screened the first 40 of them (in addition to another article identified through the citations of these articles). We ultimately included 11 articles (including 4 CDC Morbidity and Mortality Weekly Reports) generating bivalent booster VE estimates. 4 of the studies used a test-negative design (none using matching), and 7 used a cohort design (1 using matching). Full study characteristics can be found in Table A5, and a forest plot summarizing the estimates can be found in Figure 5. The 11 studies included 23,215,145 individuals (or tests in designs where tests, not individuals, were the unit of observation) in total.

**Figure 5:**
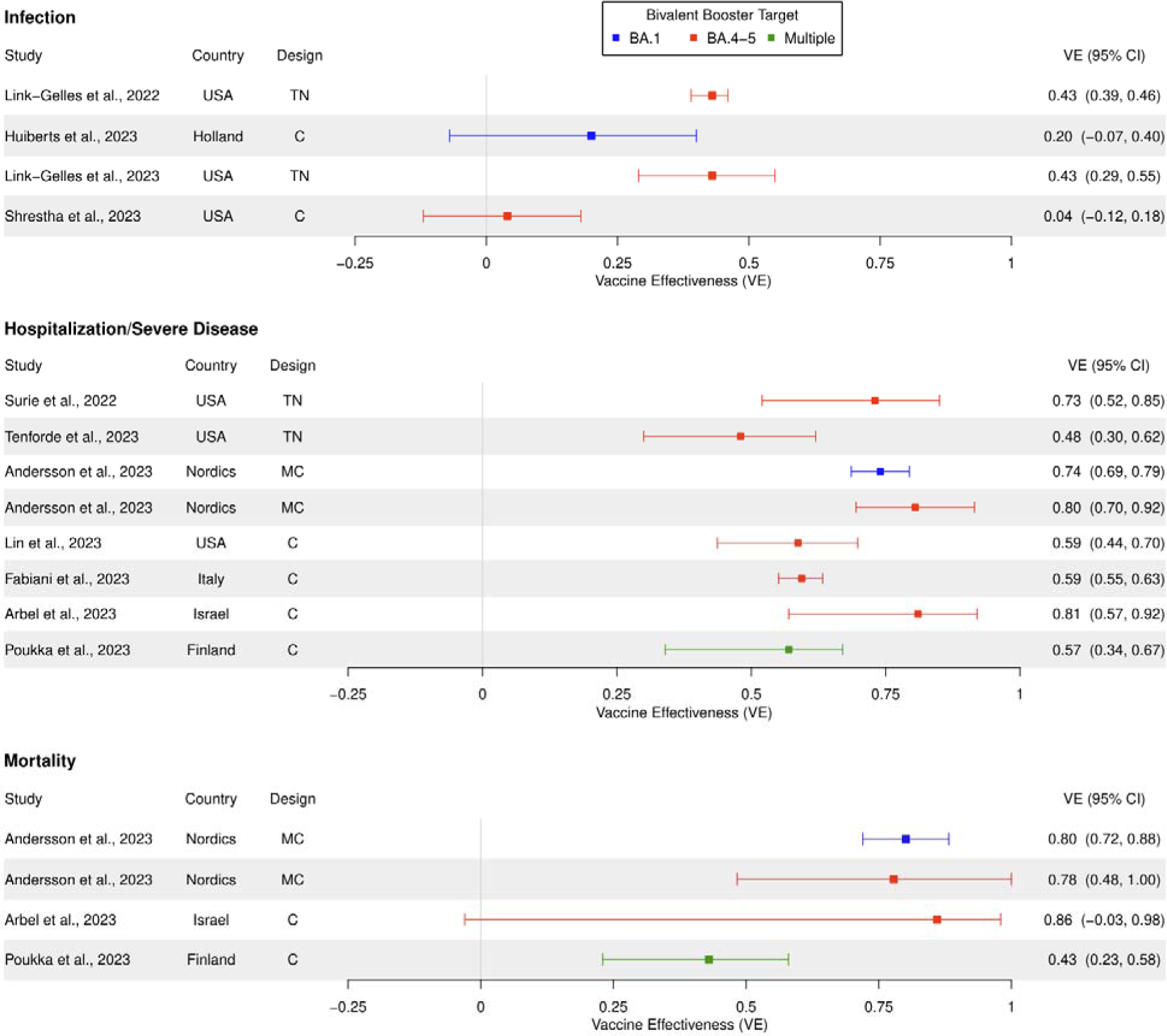
Bivalent booster (targeting either BA.1 or BA.4/5) VE compared to a previous dose against infection, hospitalization/severe disease, and mortality only in separate panels. Studies are sorted in chronological order of last month included in the study period (with publication date being used to break ties). For studies with stratified results, we displayed results corresponding to only the oldest age, most recent variant, longest time since previous dose, and a 2-month time since boosted strata. For Andersson et al., 2023,^49^ the Nordic countries included were Denmark, Finland, Norway, and Sweden (Iceland and Greenland were not included). Abbreviations: TN, test-negative; MTN, matched test-negative; CC, case-control (non-test-negative); MCC, matched case-control (non-test-negative); C, cohort; MC, matched cohort.

VE against infection was unclear, with two cohort studies failing to find a VE significantly different from zero, and two test-negative studies finding a VE of 43%. VE against hospitalization/severe disease however was much higher, in the range of 48%-81%. VE against death was close to 80% in most studies, but was also characterized by wide confidence intervals due to limited follow-up data.

### Michigan Medicine Data Analysis

#### Study Population

The MM EHR cohort corresponding to the main analysis (first booster versus primary series) included 186,495 individuals overall, with 153,811 boosted before or during Q4 2021, and 32,684 fully vaccinated but not boosted before the end of Q4 2021 (i.e., eligible to be included in the comparative primary series vaccination group). A full set of descriptive statistics for the cohort can be found in Table A6. Boosted individuals were generally older (57.0 vs. 47.6 average age for boosted and fully vaccinated, respectively) and had a higher percentage of non-Hispanic whites (77.5% vs. 68.6%) than individuals not boosted by Q4 2021. In addition, they had more comorbidities (1.01 vs 0.87 average Charlson Index) and were, on average, from less affluent neighborhoods (47.1% vs. 38.7% of individuals in the lowest quartile). Both groups were majority female (by self-reporting), but boosted individuals were a higher proportion male (42.9% vs. 39.9%). Rates of working in healthcare (1.6% vs. 2.3%) and number of negative tests (1.35 vs. 1.47 average) were slightly lower for boosted than fully vaccinated individuals. There were not notable differences between the boosted and fully vaccinated in BMI (28.7 vs. 28.9 average), population density of residence area (24.5% vs 25.0% in least dense quartile), and proportion receiving primary care at Michigan Medicine (51.2% vs. 51.6%). Study population characteristics for the test-negative analysis are given in Table A7 and display similar patterns.

In the cohort analysis, 7.77% of boosted and 6.55% of fully vaccinated individuals had a positive test in the follow-up period, while 0.40% of boosted and 0.38% of fully vaccinated individuals had severe disease. 5.21% of test-positives (of any vaccination status) developed severe disease.

#### Estimated Vaccine Effectiveness

VE estimates obtained by applying different statistical methodologies for infection and severe disease outcomes using the cohort design are shown in Figure 6. Associated Kaplan-Meier curves can be found in Figure A7. A summary of the propensity score models can be found in Table A10. Corresponding results using the test-negative design can be found in Figures A10-A12. Results using different study start dates can be found in Figures A8-A9 for the cohort design, and A11 and A13 for the test-negative design.

**Figure 6:**
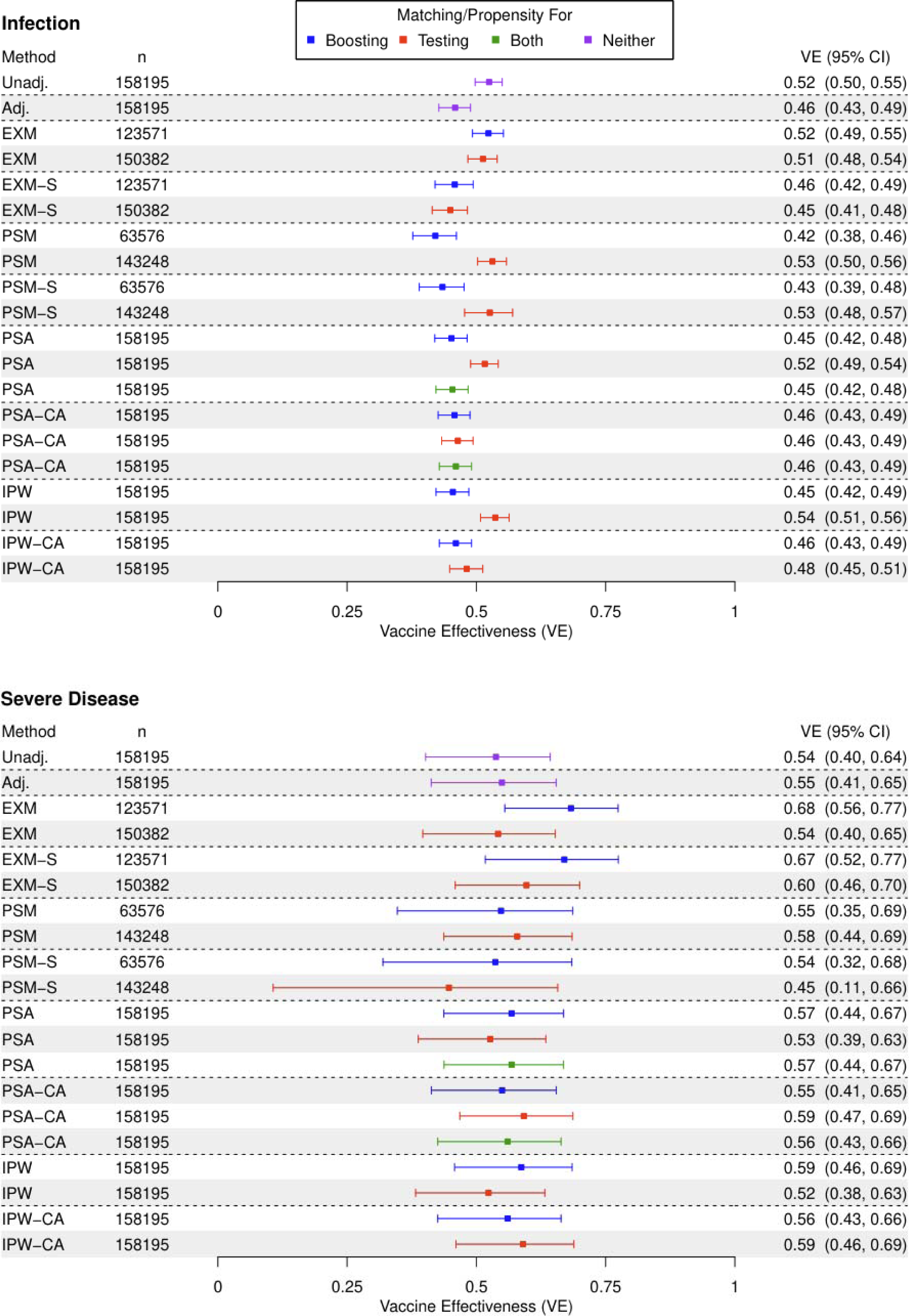
First booster/third dose vs. primary series/second dose VE against infection (first panel) and severe disease (second panel) with various matching designs during a study period of October 1, 2021 to December 31, 2022. Abbreviations: Unadj., unadjusted; Adj., adjusted for covariates; EXM, exact matching; EXM-S, exact matching with matching strata stratification in Cox regression; PSM, propensity score matching with a 0.2 caliper; PSM-S, propensity score matching with a 0.2 caliper and matching strata stratification in Cox regression; PSA; adjustment for propensity score; PSA-CA, adjustment for propensity score and for covariates; IPW, inverse probability weighting of propensity score; IPW-CA, inverse probability weighting of propensity score also adjusting for covariates.

Point estimates of infection VE ranged from about 42% to 54% across different statistical methods (Figure 6). We note that either regression adjustment for the full set of covariates or accounting (with either matching, weighting, or adjustment) for propensity of receiving a booster led to more conservative VE estimates (42%-48%).

Solely adjusting, matching, or weighting for propensity of testing led to higher VE estimates compared to other methods (52%-54%). For any matching strategy, stratification of the Cox proportional hazards model led to lower VE estimates (43%-46%). Matching, in general, led to wider confidence intervals for VE due to loss of observations.

VE estimates for severe disease fell in a range from about 45% to 68%, but generally had much wider confidence intervals than infection VE because of a limited number of events (Figure 6). Excluding matching methods (i.e., methods that discard observations) however, VE ranged only from 53% to 59%. Matching methods in general had wider confidence intervals, as with infection VE. All confidence intervals for severe disease were largely overlapping.

The test-negative analysis (Figures A10, A12) demonstrated a similar pattern: VE estimates against infection ranged from 40% to 57%, while VE against severe disease, aside from some erratic results with exact matching, ranged from 65% to 75%. With the test-negative results for the infection outcome, there were two clusters of VE estimates: one for methods that involved covariate adjustment (ranging from 53% to 57%) and another for methods that did not (ranging from 41% to 47%). However, one noteworthy observation was the reduction in the length of the confidence interval by using a test-negative design, particularly when using IPW (by the propensity of boosting) and covariate adjustment. While the severity VE for the cohort design corresponding to the same method was 56% (95% CI: 43%-66%), the test-negative design VE was 67% (95% CI: 62%-72%), a much narrower confidence interval.

A necessary consideration for the use of the test-negative design to estimate severity VE is the choice of control group.^33^ Within the same method, we observed only slight variation in point estimates depending upon the choice of the control group, which could be one of test-negatives, those without severe disease regardless of test results, and test-positives without severe disease, as the control group. In all methods except exact matching, where small sample sizes produced erratic results, VE estimates from using each of the three control groups all were within a 6-percentage point interval or smaller (Figure A12).

Results for the secondary study populations can be found in the Supplementary Appendix. Both cohort and test-negative results for the vaccination group of one or more boosters can be found in Figures A14-A23, while cohort and test-negative results for the vaccination group of two or more boosters can be found in Figures A24-A33.

## Discussion

### Literature Review

#### Choice of Boosters: VE of First, Second, and Bivalent Boosters

##### First Booster

We found that the preponderance of the literature suggests that, relative to a primary series vaccination, a first booster provides strong protection against both hospitalization and death from COVID-19. This protection remained strong even in studies conducted later in 2022 (44%-80% for hospitalization, 62%-89% for death) (Figures 3, A4-A5). We however caution that we did not examine longer periods of time since vaccination beyond 3 months.

Protection against infection was much more fragile, shown most clearly in our forest plots stratified on time since vaccination, where the decrease in VE as time progressed followed a relatively linear pattern (Figure A3). These plots also conveyed a decline in protection against infection between one- and three-months post-booster (41%-59% to −4%-44%) (Figure A3).

We acknowledge that the first booster VE estimates from Kislaya et al.^30^ are outliers that we have chosen to largely ignore in our description of trends (Figure 3). In addition to producing estimates much lower than any other study in our review, this study produced much higher VE estimates for the second booster relative to the first booster than the first booster relative to primary series vaccination. We suspect selection bias between vaccination groups in this study’s cohort, which was EHR-based and limited only to older residents.

##### Second Booster

We observed that a second monovalent booster provided additional protection over the first booster against hospitalization and death (Figure 4). However, this protection was notably lower than the protection afforded by the first booster over primary series vaccination for all outcome measures. Also, protection against infection was very low in general (around 10%-30% for later studies) (Figure A6).

The VE results for the second monovalent booster suggest that there may be diminishing returns of continually boosting with the same vaccine targeting the ancestral Wuhan variant, particularly as SARS-CoV-2 continues to evolve further away from this variant. Fortunately, recent vaccine development has given the world a new option.

##### Bivalent Booster

Our primary literature search occurred at a time where there was little bivalent booster literature formally published in scientific journals, thus limiting its scope to monovalent boosters targeting the ancestral SARS-CoV-2 strain. However, many developed countries have now shifted their vaccination rollout exclusively to bivalent boosters targeting Omicron variants. While the specific VE estimates of the bivalent booster could be much different than that of the monovalent boosters that we included in our literature review, our findings on the design and analysis of VE studies are just as applicable to bivalent booster VE studies.

Our rapid review in late March 2023 offers a glimpse into the early evidence of bivalent booster effectiveness. The current results seem promising, suggesting increased effectiveness of bivalent boosters compared to monovalent boosters, at least for severe disease outcomes. Many of the more recent second monovalent booster studies reported a VE against hospitalization or severe disease of around 50% or lower, with multiple studies below 40% (Figure 4). In contrast, out of the 6 bivalent booster studies with hospitalization/severe disease VE estimates, all reported a VE of at least 48%, and the second lowest VE reported was 57% (Figure 5). Of particular interest is Lin et al.,^10^ who calculated VE of a monovalent and bivalent booster in the same study, finding a much higher VE against severe disease for the bivalent booster (61.8%, 95% CI: 48.2%-71.8%) compared to the monovalent (24.2%, 95% CI: 1.4%-42.8%). We note that not all the studies on the bivalent booster are encouraging: Poukka et al.^34^ reported sharply waning immunity, with a severe disease VE of 26% for both a hospitalization (95% CI: −9%-50%) and mortality (95% CI: −13%-51%) outcome between 61-90 days after booster administration. The jury is still out on longer follow-up time and whether a recommended annual booster is sufficient to ensure protection in the endemic stage of the pandemic.

#### Choice of Designs: Cohort and/or Test-Negative

##### Cohort Design

Cohort studies were usually viewed in a time-to-event paradigm, where the time since study start or booster administration to an outcome event of interest or censoring event was compared between fully vaccinated and boosted groups. This comparison was done using Cox regression, often adjusted for a set of covariates, or performed on a matched cohort, sometimes using stratified Cox regression with the matched sets as strata (with the final hazard ratio pooled between strata). VE was calculated as the hazard ratio from Cox regression subtracted from one, i.e., 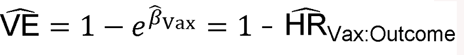.

[Censoring criteria for studies using Cox regression typically included documented COVID-19 infection (for non-infection endpoints), the end of the study, death for non-COVID-related reasons, or the administration of an additional dose of the COVID-19 vaccine. Individuals in the referent group for VE calculation were often allowed to re-enter the cohort as a member of the other group after receiving the next dose of the vaccine. A small number of studies used this post-vaccination incubation period as an alternative referent group for the effectiveness of this dose.^35^ Cohort studies using matching would often censor both sides of the matched pair when the one in the referent group received the next dose.^36^

Less commonly, cohort studies were analyzed in terms of counts of individuals at risk. In this situation, VE was calculated through a Poisson regression (where 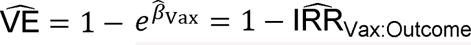). These models included calendar time as a covariate. For example, Bar-On et al. fit a model of person-days at risk for COVID-19 and adjusted for calendar day.^5^

##### Test-Negative Design

Statistical methods were very similar between test-negative designs for infection. Test-negative designs used logistic regression to estimate VE, or conditional logistic regression if they employed matching. The response variable was the test result, and explanatory variables were the vaccination status of the individual at the time of the test, along with other adjustment covariates. VE was calculated as the implied odds ratio for the vaccination status term subtracted from 1, i.e., 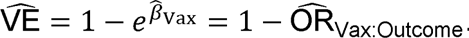.

Test-negative designs were not very common for outcomes besides infection, as testing data essentially serves as a case-control for infection, but not other outcomes. Most severe disease endpoints were evaluated with a cohort design. Nonetheless, a few test-negative studies tackled severe disease endpoints, though the reference group for severe disease often varied between those who tested negative and any who did not experience severe disease, regardless of test results. We give a more detailed overview of this choice of control group in Supplementary Methods A3.

As an alternative strategy, a few test-negative studies used a secondary matched non-test-negative case-control design to estimate VE against severe disease endpoints.^37^ A few studies that evaluated VE against hospitalization but not infection tackled this control-group problem by designing a case-control sample among those hospitalized, not those presenting for infection, with matching used to ensure the appropriateness of controls.^38^ Another case-control design was used by Silva-Valencia et al.^39^ where cases were those dead from COVID-19, and controls were risk-set sampled and matched from the healthy at-risk population at the time of the case’s death.

#### Choice of Analysis: Covariate Adjustment, Matching, Weighting, and Stratification

##### Choice of Adjustment or Matching Covariates

Regression adjustment covariates (or matching covariates in studies using matching and not adjusting further on these covariates) differed somewhat between studies, but generally followed similar patterns. Most studies adjusted or matched on age and sex, and many also on one or more covariates related to the subject’s economic status (including race/ethnicity and living conditions), along with comorbidities, as either a comorbidity index or a set of several covariates each responding to individual comorbidities. Studies with longer follow-up periods had adjustment for time periods at risk. Test-negative studies often adjusted for the time of the test. Some studies adjusted for exposure to disease in healthcare settings. Without the convenient built-in control that a test-negative design offered, many cohort studies also adjusted for the number of negative PCR tests taken either before the study or in total before the study end date, accounting in some sense for inclination for testing.^40^

There were several studies included in this review that presented completely unadjusted VE metrics.^17, 41–43^ These were often shorter articles or preprints, often from countries or populations with little prior (English-language) booster VE literature.

##### Matching and Weighting

Matching was a strategy used in some but not all test-negative and cohort designs. It served as an alternative way to adjust for covariates beyond the standard regression adjustment used in unmatched studies. Some matched studies used matching in lieu of regression adjustment entirely, having entirely unadjusted regressions after matching.

Others matched on some covariates and adjusted for a few remaining ones. A small number of studies both matched on and regression-adjusted for numeric covariates, typically binned into several categories in the matching step and then either treated as a continuous variable or modeled with splines within the categories in the regression step.^44^

The most popular type of matching performed was exact matching, where only individuals with exact matches in all considered covariates were matched. This generally consisted of binning variables into several bins or categories before performing exact matching. Observations were matched either in pairs or unbalanced ratios of every subject sharing the same exact matching values depending on the study. Observations not receiving an exact match were discarded.

A less common method found in the literature was propensity score matching. This consisted first of the generation of a propensity for receiving a booster through the regression of the covariates on the vaccination status variable. Pairs of subjects with different exposure statuses were then matched with a nearest neighbor algorithm (Table 1). Some studies introduced a caliper, or maximum allowable distance between propensity scores for matching observations, with matches with a greater distance discarded. Previous literature including Austin^45^ has suggested a caliper of 0.2 standard deviations of the logit of the propensity score as a recommended value, and this was the value typically used by studies using propensity score matching.^46^

Even less common in the literature was the use of a propensity score not via matching, but with IPW instead.^16^ This involved weighting observations in the study sample by the inverse of their propensity for vaccination/booster (more detail is included in Table 1).

Compared to propensity score matching with a caliper, IPW provides potential efficiency benefits by not fully excluding any observations.

##### Stratification

Stratification was a key area where different strategies were employed by different studies. Many studies did not stratify their results at all, but those that did often chose different stratification variables. One of the most popular stratification conditions was time since booster vaccination (for a test or contributed person-time), a condition that could capture the process of waning immunity. A parallel consideration was stratification or inclusion criteria on time since primary series vaccination for the referent group; shorter amounts of time provide a comparison of two fresh vaccine regimens, while a longer period of time provides a more useful policy decision on the utility of vaccination for those booster-eligible (in some cases both were provided; Mattiuzzi and Lippi^43^ provided strata before and after booster eligibility). Another popular stratification condition, at least in countries with different widely used vaccinations, was brand of booster, primary series, or entire vaccination series, with mRNA boosters sometimes (but often not) being combined in one stratum. A type of stratification used in studies with longer follow-up time was stratification on type of variant (usually done on a date of variant dominance, as opposed to genetic testing of cases), done most commonly in studies that spanned both Delta and Omicron periods.^47^

##### Limitations

There are several notable limitations of our review that may prevent it from being truly representative of the entirety of booster VE literature and provide a word of disclaimer to its interpretation. First and foremost, we limited our search to English-language articles only. While our search was still able to include articles from a broad selection of countries, this may have systematically excluded literature from certain areas of the world. Also, we chose to limit the scope of the review to articles that estimated VE of a booster relative to the previous dose in the vaccine regimen only. Many booster VE articles, particularly for the first booster, evaluated VE as relative to no vaccination, and such articles (including 47 of 116 VE articles in our search) were excluded from our search (Figure A1). Additionally, our forest plots, even after stratification by time since booster, grouped together in the same plot articles with major differences in design and even their target estimand (e.g., prospective versus retrospective studies, symptomatic versus all types of infection, etc.), which may affect the interpretation of such plots.

### Michigan Medicine Data Analysis

The VE estimates from our MM data analysis provided results that were relatively consistent with the literature review for the US during the relevant time period, a reassuring occurrence.

Nevertheless, the main utility of the data analysis is not the specific VE estimates, but that it allowed us to apply many of the methods we identified through our literature review (and a few new ones) to the same healthcare system (with the caveat that the cohort and test-negative design have different definitions of study cohorts).

It should be noted that the methods we examined even under the same design do not all target the same estimand. First and foremost, exact and propensity score matching alter the study population by matching on the treated units and discarding observations without satisfactory matches. Secondly, we included methods that estimated either marginal or conditional relationships. A comparison between different matching/propensity score methods is therefore not able to decompose differences in the true parameter or estimator. While we keep this fact in mind and refer to it when relevant, we note however that such a distinction is not crucial to the purposes of this comparison: we intended to document how different analysis decisions can change VE estimates, regardless of what side of the decomposition leads to such changes. As observed in the literature review, VE studies in practice have targeted either marginal or conditional estimands, and have certainly differed, often systematically, in target population, so such a comparison across different estimands serves only to better reflect the landscape in the literature.

For multiple methods, the infection VE using only propensity for testing was notably higher than the VE using propensity for boosting or adjustment for covariates. In fact, the VE using only propensity for testing was closer to an unadjusted estimate. This may indicate, with an infection outcome, accounting for propensity of testing alone is not sufficient to control confounding bias.

However, using a propensity score for either boosting or testing did not make a notable difference for a severe disease outcome. No statistical method had a non-overlapping confidence interval for severity VE. This is in part because confidence intervals were generally wider with a limited number of events, but, particularly excluding matching methods that discard observations, point estimates were also largely in agreement.

Generally, our results suggest that inference regarding a severe disease outcome is more robust to changes in statistical methodology than an infection outcome, even between models targeting conditional and marginal average treatment effects.

The takeaway message from our analysis of the test-negative design suggests that, for infection VE, we may need to adjust for an expanded set of covariates despite the design attempting to control for confounding due to healthcare seeking behavior. Also, the test-negative design with weighting and covariate adjustment led to narrower confidence intervals for the severity VE compared to the cohort design. Finally, applying the test-negative design to severe disease involves a choice of several different control groups, but this only seems to lead to slight differences in results.

## Conclusion

As we advance into the endemic stage of the COVID-19 pandemic, infections are less likely to be reported. Our study indicates severity endpoints are more robust to choice of statistical methodology than infection endpoints. However, since severe COVID-19 (as defined as hospitalization or death) is a rare outcome in a world with hybrid immunity,^14^ test-negative designs, which can be extended to severe disease outcomes, may offer more efficiency when properly used. But when using test-negative designs for infection, one must adjust for an expanded set of relevant covariates instead of purely relying on the design properties to mitigate confounding. Finally, our work is not just relevant to COVID-19 vaccines, but potentially may apply to other vaccination assessments as well.

## Data Availability

Aggregated data files collected in the literature review and code used for visualization and analysis are available online at https://github.com/smeah/COVID19BoosterVE. Data from the Michigan Medicine vaccine effectiveness analysis cannot be shared publicly due to patient confidentiality. The data are available from the University of Michigan Precision Health Analytics Platform at https://precisionhealth.umich.edu/tools-resources/data-access-tools/ for researchers who meet the criteria for access to confidential data.

## Acknowledgements

We acknowledge the Precision Health at the University of Michigan and the University of Michigan Medical School Data Office for Clinical and Translational Research for providing data storage, management, processing, and distribution services. We thank Judith Smith from the University of Michigan Library for assistance with developing the keyword blocks for the database search and methods for the literature review. We also thank Dr. Yair Goldberg for providing comments on a previous draft of the manuscript. The study was supported by NSF DMS 1712933 and NIH/NIGMS R01GM139926.

## References

1. Polack FP, Thomas SJ, Kitchin N, et al. Safety and Efficacy of the BNT162b2 mRNA Covid-19 Vaccine. N Engl J Med. 2020;383(27):2603–2615. doi:10.1056/NEJMoa2034577

2. Thompson MG, Burgess JL, Naleway AL, et al. Prevention and Attenuation of Covid-19 with the BNT162b2 and mRNA-1273 Vaccines. N Engl J Med. 2021;385(4):320–329. doi:10.1056/NEJMoa2107058

3. Tenforde MW, Self WH, Naioti EA, et al. Sustained Effectiveness of Pfizer-BioNTech and Moderna Vaccines Against COVID-19 Associated Hospitalizations Among Adults — United States, March–July 2021. Morb Mortal Wkly Rep. 2021;70(34):1156-1162. doi:10.15585/mmwr.mm7034e2

4. Goldberg Y, Mandel M, Bar-On YM, et al. Waning Immunity after the BNT162b2 Vaccine in Israel. N Engl J Med. 2021;385(24):e85. doi:10.1056/NEJMoa2114228

5. Bar-On YM, Goldberg Y, Mandel M, et al. Protection of BNT162b2 Vaccine Booster against Covid-19 in Israel. N Engl J Med. 2021;385(15):1393–1400. doi:10.1056/NEJMoa2114255

6. Barouch DH. Covid-19 Vaccines — Immunity, Variants, Boosters. N Engl J Med. 2022;387(11):1011–1020. doi:10.1056/NEJMra2206573

7. Wise J. Covid-19: New coronavirus variant is identified in UK. BMJ. 2020;371:m4857. doi:10.1136/bmj.m4857

8. Mahase E. Covid-19: What new variants are emerging and how are they being investigated? BMJ. 2021;372:n158. doi:10.1136/bmj.n158

9. Surie D. Early Estimates of Bivalent mRNA Vaccine Effectiveness in Preventing COVID-19–Associated Hospitalization Among Immunocompetent Adults Aged ≥65 Years — IVY Network, 18 States, September 8–November 30, 2022. MMWR Morb Mortal Wkly Rep. 2022;71. doi:10.15585/mmwr.mm715152e2

10. Lin DY, Xu Y, Gu Y, et al. Effectiveness of Bivalent Boosters against Severe Omicron Infection. N Engl J Med. 2023;388(8):764–766. doi:10.1056/NEJMc2215471

11. Arbel R, Peretz A, Sergienko R, et al. Effectiveness of the Bivalent mRNA Vaccine in Preventing Severe COVID-19 Outcomes: An Observational Cohort Study. Published online January 3, 2023. doi:10.2139/ssrn.4314067

12. Chen Z, Zheng W, Wu Q, et al. Global diversity of policy, coverage, and demand of COVID-19 vaccines: a descriptive study. BMC Med. 2022;20(1):130. doi:10.1186/s12916-022-02333-0

13. Wong MK, Brooks DJ, Ikejezie J, et al. COVID-19 Mortality and Progress Toward Vaccinating Older Adults — World Health Organization, Worldwide, 2020–2022. Morb Mortal Wkly Rep. 2023;72(5):113-118. doi:10.15585/mmwr.mm7205a1

14. CDC. COVID Data Tracker. Centers for Disease Control and Prevention. Published May 4, 2023. Accessed May 4, 2023. https://covid.cdc.gov/covid-data-tracker

15. Spitzer A, Angel Y, Marudi O, et al. Association of a Third Dose of BNT162b2 Vaccine With Incidence of SARS-CoV-2 Infection Among Health Care Workers in Israel. JAMA. 2022;327(4):341–349. doi:10.1001/jama.2021.23641

16. Andersson NW, Thiesson EM, Baum U, et al. Comparative effectiveness of heterologous booster schedules with AZD1222, BNT162b2, or mRNA-1273 vaccines against COVID-19 during omicron predominance in the Nordic countries. medRxiv. Published online 2022. doi:10.1101/2022.11.24.22282651

17. Desai A, Deepak P, Cross RK, et al. Effect of 2 vs 3 Doses of COVID-19 Vaccine in Patients With Inflammatory Bowel Disease: A Population-based Propensity Matched Analysis. Inflamm Bowel Dis. Published online 2022. doi:10.1093/ibd/izac252

18. Tartof SY, Slezak JM, Puzniak L, et al. Effectiveness of a third dose of BNT162b2 mRNA COVID-19 vaccine in a large US health system: A retrospective cohort study. Lancet Reg Health Am. 2022;9:100198. doi:10.1016/j.lana.2022.100198

19. Butt AA, Talisa VB, Shaikh OS, Omer SB, Mayr FB. Relative Vaccine Effectiveness of a SARS-CoV-2 mRNA Vaccine Booster Dose Against the Omicron Variant. Clin Infect Dis Off Publ Infect Dis Soc Am. Published online May 2022:ciac328. doi:10.1093/cid/ciac328

20. Zeng B, Gao L, Zhou Q, Yu K, Sun F. Effectiveness of COVID-19 vaccines against SARS-CoV-2 variants of concern: a systematic review and meta-analysis. BMC Med. 2022;20(1):200. doi:10.1186/s12916-022-02397-y

21. Ouzzani M, Hammady H, Fedorowicz Z, Elmagarmid A. Rayyan—a web and mobile app for systematic reviews. Syst Rev. 2016;5(1):210. doi:10.1186/s13643-016-0384-4

22. Page MJ, McKenzie JE, Bossuyt PM, et al. The PRISMA 2020 statement: an updated guideline for reporting systematic reviews. BMJ. 2021;372:n71. doi:10.1136/bmj.n71

23. R Core Team. R: A language and environment for statistical computing. https://www.r-project.org/

24. Abu-Raddad LJ, Chemaitelly H, Ayoub HH, et al. Effect of mRNA Vaccine Boosters against SARS-CoV-2 Omicron Infection in Qatar. N Engl J Med. 2022;386(19):1804–1816. doi:10.1056/NEJMoa2200797

25. Arbel R, Hammerman A, Sergienko R, et al. BNT162b2 Vaccine Booster and Mortality Due to Covid-19. N Engl J Med. 2021;385(26):2413–2420. doi:10.1056/NEJMoa2115624

26. Lin DY, Gu Y, Xu Y, et al. Association of Primary and Booster Vaccination and Prior Infection With SARS-CoV-2 Infection and Severe COVID-19 Outcomes. JAMA. 2022;328(14):1415–1426. doi:10.1001/jama.2022.17876

27. Clarke P, Melendez R. National Neighborhood Data Archive (NaNDA): Neighborhood Socioeconomic and Demographic Characteristics of Census Tracts, United States, 2000-2010. Published online August 28, 2019. doi:10.3886/E111107V1

28. Wu P, Gifford A, Meng X, et al. Mapping ICD-10 and ICD-10-CM Codes to Phecodes: Workflow Development and Initial Evaluation. JMIR Med Inform. 2019;7(4):e14325. doi:10.2196/14325

29. Charlson ME, Charlson RE, Peterson JC, Marinopoulos SS, Briggs WM, Hollenberg JP. The Charlson comorbidity index is adapted to predict costs of chronic disease in primary care patients. J Clin Epidemiol. 2008;61(12):1234–1240. doi:10.1016/j.jclinepi.2008.01.006

30. Kislaya I, Machado A, Magalhães S, et al. COVID-19 mRNA vaccine effectiveness (second and first booster dose) against hospitalisation and death during Omicron BA.5 circulation: cohort study based on electronic health records, Portugal, May to July 2022. Eurosurveillance. 2022;27(37). doi:10.2807/1560-7917.ES.2022.27.37.2200697

31. Grewal R, Kitchen SA, Nguyen L, et al. Effectiveness of a fourth dose of covid-19 mRNA vaccine against the omicron variant among long term care residents in Ontario, Canada: test negative design study. BMJ. 2022;378:e071502. doi:10.1136/bmj-2022-071502

32. Grewal R, Nguyen L, Buchan SA, Wilson SE, Costa AP, Kwong JC. Effectiveness and Duration of Protection of a Fourth Dose of COVID-19 mRNA Vaccine among Long-Term Care Residents in Ontario, Canada. medRxiv. Published online 2022. doi:10.1101/2022.09.29.22280526

33. Shi X, Li KQ, Mukherjee B. Current Challenges With the Use of Test-Negative Designs for Modeling COVID-19 Vaccination and Outcomes. Am J Epidemiol. 2023;192(3):328–333. doi:10.1093/aje/kwac203

34. Poukka E, Goebeler S, Nohynek H, Leino T, Baum U. Bivalent booster effectiveness against severe COVID-19 outcomes in Finland, September 2022 — January 2023. Published online March 5, 2023:2023.03.02.23286561. doi:10.1101/2023.03.02.23286561

35. Korves C, Izurieta HS, Smith J, et al. Relative effectiveness of booster vs. 2-dose mRNA Covid-19 vaccination in the Veterans Health Administration: Self-controlled risk interval analysis. medRxiv. Published online 2022. doi:10.1101/2022.03.17.22272555

36. Magen O, Waxman JG, Makov-Assif M, et al. Fourth Dose of BNT162b2 mRNA Covid-19 Vaccine in a Nationwide Setting. N Engl J Med. 2022;386(17):1603–1614. doi:10.1056/NEJMoa2201688

37. Patalon T, Gazit S, Pitzer VE, Prunas O, Warren JL, Weinberger DM. Short Term Reduction in the Odds of Testing Positive for SARS-CoV-2; a Comparison Between Two Doses and Three doses of the BNT162b2 Vaccine. Published online August 31, 2021:2021.08.29.21262792. doi:10.1101/2021.08.29.21262792

38. Ridgway JP, Tideman S, French T, et al. Odds of Hospitalization for COVID-19 After 3 vs 2 Doses of mRNA COVID-19 Vaccine by Time Since Booster Dose. JAMA. 2022;328(15):1559–1561. doi:10.1001/jama.2022.17811

39. Silva-Valencia J, Soto-Becerra P, Escobar-Agreda S, et al. Relative vaccine effectiveness of the booster dose of COVID-19 vaccine for preventing death in individuals with a primary regimen based on the BBIBP-CorV, ChAdOx1-S, or BNT162b2 vaccines during the Omicron wave in Peru: A nested case-control study using national population data. Vaccine. 2022;40(45):6512-6519. doi:10.1016/j.vaccine.2022.09.066

40. Barda N, Dagan N, Cohen C, et al. Effectiveness of a third dose of the BNT162b2 mRNA COVID-19 vaccine for preventing severe outcomes in Israel: an observational study. Lancet Lond Engl. 2021;398(10316):2093–2100. doi:10.1016/S0140-6736(21)02249-2

41. Saito Y, Yamamoto K, Takita M, Kami M, Tsubokura M, Shibuya K. Effectiveness of the Booster of SARS-CoV-2 Vaccine among Japanese Adolescents: A Cohort Study. Vaccines. 2022;10(11). doi:10.3390/vaccines10111914

42. Tai CG, Maragakis LL, Connolly S, et al. Booster protection against Omicron infection in a highly vaccinated cohort. medRxiv. Published online 2022. doi:10.1101/2022.02.24.22271347

43. Mattiuzzi C, Lippi G. Primary COVID-19 vaccine cycle and booster doses efficacy: analysis of Italian nationwide vaccination campaign. Eur J Public Health. 2022;32(2):328–330. doi:10.1093/eurpub/ckab220

44. Ranzani OT, Hitchings MDT, de Melo RL, et al. Effectiveness of an Inactivated Covid-19 Vaccine with Homologous and Heterologous Boosters against the Omicron (B.1.1.529) Variant. medRxiv. Published online 2022. doi:10.1101/2022.03.30.22273193

45. Austin PC. Optimal caliper widths for propensity-score matching when estimating differences in means and differences in proportions in observational studies. Pharm Stat. 2011;10(2):150–161. doi:10.1002/pst.433

46. Ioannou GN, Bohnert ASB, O’Hare AM, et al. Effectiveness of mRNA COVID-19 vaccine boosters against infection, hospitalization and death: A target trial emulation in the omicron (B.1.1.529) variant era. medRxiv. Published online 2022. doi:10.1101/2022.06.15.22276466

47. Suah JL, Tng BH, Tok PSK, et al. Real-world effectiveness of homologous and heterologous BNT162b2, CoronaVac, and AZD1222 booster vaccination against Delta and Omicron SARS-CoV-2 infection. Emerg Microbes Infect. 2022;11(1):1343-1345. doi:10.1080/22221751.2022.2072773

48. McConeghy KW, Bardenheier B, Huang AW, et al. Effectiveness of a SARS-CoV-2 mRNA vaccine booster dose for prevention of infection, hospitalization or death in two nation-wide nursing home systems. medRxiv. Published online 2022. doi:10.1101/2022.01.25.22269843

49. Andersson NW, Thiesson EM, Baum U, et al. Comparative effectiveness of the bivalent BA.4–5 and BA.1 mRNA-booster vaccines in the Nordic countries. Published online January 19, 2023:2023.01.19.23284764. doi:10.1101/2023.01.19.23284764

50. Accorsi EK, Britton A, Fleming-Dutra KE, et al. Association Between 3 Doses of mRNA COVID-19 Vaccine and Symptomatic Infection Caused by the SARS-CoV-2 Omicron and Delta Variants. JAMA. 2022;327(7):639–651. doi:10.1001/jama.2022.0470

51. Patalon T, Gazit S, Pitzer VE, Prunas O, Warren JL, Weinberger DM. Odds of Testing Positive for SARS-CoV-2 Following Receipt of 3 vs 2 Doses of the BNT162b2 mRNA Vaccine. JAMA Intern Med. 2022;182(2):179–184. doi:10.1001/jamainternmed.2021.7382

52. Andrews N, Stowe J, Kirsebom F, Gower C, Ramsay M, Bernal JL. Effectiveness of BNT162b2 (Comirnaty, Pfizer-BioNTech) COVID-19 booster vaccine against covid-19 related symptoms in England: test negative case-control study. Published online November 15, 2021:2021.11.15.21266341. doi:10.1101/2021.11.15.21266341

53. Bar-On YM, Goldberg Y, Mandel M, et al. Protection against Covid-19 by BNT162b2 Booster across Age Groups. N Engl J Med. 2021;385(26):2421–2430. doi:10.1056/NEJMoa2115926

54. Patalon T, Saciuk Y, Peretz A, et al. Waning effectiveness of the third dose of the BNT162b2 mRNA COVID-19 vaccine. Nat Commun. 2022;13(1):3203. doi:10.1038/s41467-022-30884-6

55. Prasad N, Derado G, Nanduri SA, et al. Effectiveness of a COVID-19 Additional Primary or Booster Vaccine Dose in Preventing SARS-CoV-2 Infection Among Nursing Home Residents During Widespread Circulation of the Omicron Variant - United States, February 14-March 27, 2022. MMWR Morb Mortal Wkly Rep. 2022;71(18):633-637. doi:10.15585/mmwr.mm7118a4

56. Marra AR, Miraglia JL, Malheiros DT, et al. Effectiveness of heterologous COVID-19 vaccine booster dosing in Brazilian healthcare workers, 2021. Clin Infect Dis Off Publ Infect Dis Soc Am. Published online May 2022. doi:10.1093/cid/ciac430

57. Amir O, Goldberg Y, Mandel M, et al. Protection following BNT162b2 booster in adolescents substantially exceeds that of a fresh 2-dose vaccine. Nat Commun. 2022;13(1):1971. doi:10.1038/s41467-022-29578-w

58. Andrews N, Stowe J, Kirsebom F, et al. Effectiveness of COVID-19 booster vaccines against COVID-19-related symptoms, hospitalization and death in England. Nat Med. 2022;28(4):831–837. doi:10.1038/s41591-022-01699-1

59. Lind ML, Robertson AJ, Silva J, et al. Effectiveness of Primary and Booster COVID-19 mRNA Vaccination against Omicron Variant SARS-CoV-2 Infection in People with a Prior SARS-CoV-2 Infection. medRxiv. Published online 2022. doi:10.1101/2022.04.19.22274056

60. Monge S, Rojas-Benedicto A, Olmedo C, et al. Effectiveness of mRNA vaccine boosters against infection with the SARS-CoV-2 omicron (B.1.1.529) variant in Spain: a nationwide cohort study. Lancet Infect Dis. 2022;22(9):1313-1320. doi:10.1016/S1473-3099(22)00292-4

61. Muhsen K, Maimon N, Mizrahi AY, et al. Association of BNT162b2 Vaccine Third Dose Receipt With Incidence of SARS-CoV-2 Infection, COVID-19-Related Hospitalization, and Death Among Residents of Long-term Care Facilities, August to October 2021. JAMA Netw Open. 2022;5(7):e2219940. doi:10.1001/jamanetworkopen.2022.19940

62. Richterman A, Behrman A, Brennan PJ, O’Donnell JA, Snider CK, Chaiyachati KH. Durability of SARS-CoV-2 mRNA Booster Vaccine Protection Against Omicron Among Health Care Workers with a Vaccine Mandate. Clin Infect Dis Off Publ Infect Dis Soc Am. Published online June 2022. doi:10.1093/cid/ciac454

63. Saciuk Y, Kertes J, Shamir Stein N, Ekka Zohar A. Effectiveness of a Third Dose of BNT162b2 mRNA Vaccine. J Infect Dis. 2022;225(1):30–33. doi:10.1093/infdis/jiab556

64. Sharma A, Oda G, Holodniy M. Effectiveness of a third dose of BNT162b2 or mRNA-1273 vaccine for preventing post-vaccination COVID-19 infection: an observational study. medRxiv. Published online 2021. doi:10.1101/2021.11.29.21266777

65. Sheikh A, Kerr S, Woolhouse M, McMenamin J, Robertson C. Severity of omicron variant of concern and effectiveness of vaccine boosters against symptomatic disease in Scotland (EAVE II): a national cohort study with nested test-negative design. Lancet Infect Dis. 2022;22(7):959–966. doi:10.1016/S1473-3099(22)00141-4

66. Wan J, Cazer CL, Clarkberg ME, et al. Boosters protect against SARS-CoV-2 infections in young adults during an Omicron-predominant period. medRxiv. Published online 2022. doi:10.1101/2022.03.08.22272056

67. Miyauchi S, Hiyama T, Nakano Y, et al. Real-World Effectiveness of a Booster Dose of the COVID-19 Vaccines among Japanese University Students. Vaccines. 2022;10(8). doi:10.3390/vaccines10081283

68. Hulme WJ, Williamson EJ, Horne E, et al. Effectiveness of BNT162b2 booster doses in England: an observational study in OpenSAFELY-TPP. medRxiv. Published online 2022. doi:10.1101/2022.06.06.22276026

69. Ng OT, Marimuthu K, Lim N, et al. Analysis of COVID-19 Incidence and Severity Among Adults Vaccinated With 2-Dose mRNA COVID-19 or Inactivated SARS-CoV-2 Vaccines With and Without Boosters in Singapore. JAMA Netw Open. 2022;5(8):e2228900. doi:10.1001/jamanetworkopen.2022.28900

70. Robilotti EV, Whiting K, Lucca A, et al. Effectiveness of mRNA booster vaccine among health Care workers in New York City during the omicron surge, December 2021-January 2022. Clin Microbiol Infect Off Publ Eur Soc Clin Microbiol Infect Dis. Published online August 2022:S1198-743X(22)00385-8. doi:10.1016/j.cmi.2022.07.017

71. Chemaitelly H, Ayoub HH, Tang P, et al. Long-term COVID-19 booster effectiveness by infection history and clinical vulnerability and immune imprinting. medRxiv. Published online 2022. doi:10.1101/2022.11.14.22282103

72. Florea A, Sy LS, Qian L, et al. Effectiveness of mRNA-1273 vaccine booster against COVID-19 in immunocompetent adults. Clin Infect Dis Off Publ Infect Dis Soc Am. Published online 2022. doi:10.1093/cid/ciac785

73. González S, Olszevicki S, Gaiano A, et al. Protection of homologous and heterologous boosters after primary schemes of rAd26-rAd5, ChAdOx1 nCoV-19 and BBIBP-CorV during the Omicron outbreak in adults of 50 years and older in Argentina: a test-negative case-control study. medRxiv. Published online 2022. doi:10.1101/2022.09.25.22280341 control+study.&stitle=medRxiv&title=medRxiv&volume=&issue=&spage=&epage=& aulast=Gonz%C3%A1lez&aufirst=Soledad&auinit=S.&aufull=Gonz%C3%A1lez+S.& coden=&isbn=&pages=-&date=2022&auinit1=S&auinitm=

74. Laake I, Skodvin SN, Blix K, et al. Effectiveness of mRNA Booster Vaccination Against Mild, Moderate, and Severe COVID-19 Caused by the Omicron Variant in a Large, Population-Based, Norwegian Cohort. J Infect Dis. 2022;226(11):1924-1933. doi:10.1093/infdis/jiac419

75. Mehta HB, Li S, Goodwin JS. Effectiveness of COVID-19 Booster on the Risk of Hospitalization Among Medicare Beneficiaries. Mayo Clin Proc. 2022;97(10):1780–1793. doi:10.1016/j.mayocp.2022.06.029

76. Sim JY, Wu PS, Cheng CF, Chao YC, Yu CH. Effectiveness of Booster and Influenza Vaccines against COVID-19 among Healthcare Workers, Taiwan. Emerg Infect Dis. 2022;28(10):2126–2130. doi:10.3201/eid2810.221134

77. Sonmezer MC, Dizman GT, Erul E, et al. Relative Vaccine Effectiveness of the Third Dose of CoronaVac or BNT162b2 Following a Two-Dose CoronaVac Regimen: A Prospective Observational Cohort Study from an Adult Vaccine Center in Turkey. Vaccines. 2022;10(7). doi:10.3390/vaccines10071140

78. Stirrup O, Shrotri M, Adams NL, et al. Clinical effectiveness of SARS-CoV-2 booster vaccine against Omicron infection in residents and staff of Long-Term Care Facilities: a prospective cohort study (VIVALDI). medRxiv. Published online 2022. doi:10.1101/2022.08.08.22278532

79. Regev-Yochay G, Gonen T, Gilboa M, et al. Efficacy of a Fourth Dose of Covid-19 mRNA Vaccine against Omicron. N Engl J Med. 2022;386(14):1377–1380. doi:10.1056/NEJMc2202542

80. Arbel R, Sergienko R, Friger M, et al. Effectiveness of a second BNT162b2 booster vaccine against hospitalization and death from COVID-19 in adults aged over 60 years. Nat Med. 2022;28(7):1486–1490. doi:10.1038/s41591-022-01832-0

81. Bar-On YM, Goldberg Y, Mandel M, et al. Protection by a Fourth Dose of BNT162b2 against Omicron in Israel. N Engl J Med. 2022;386(18):1712–1720. doi:10.1056/NEJMoa2201570

82. Cohen MJ, Oster Y, Moses AE, et al. Effectiveness of the BNT162b vaccine fourth dose in reducing SARS-CoV-2 infection among healthcare workers in Israel, a multi-center cohort study. medRxiv. Published online 2022. doi:10.1101/2022.04.11.22273327

83. Gazit S, Saciuk Y, Perez G, Peretz A, Pitzer VE, Patalon T. Short term, relative effectiveness of four doses versus three doses of BNT162b2 vaccine in people aged 60 years and older in Israel: retrospective, test negative, case-control study. BMJ. 2022;377:e071113. doi:10.1136/bmj-2022-071113

84. Nordström P, Ballin M, Nordström A. Effectiveness of a fourth dose of mRNA COVID-19 vaccine against all-cause mortality in long-term care facility residents and in the oldest old: A nationwide, retrospective cohort study in Sweden. Lancet Reg Health Eur. Published online July 2022:100466. doi:10.1016/j.lanepe.2022.100466

85. Kiss Z, Wittmann I, Polivka L, et al. Nationwide Effectiveness of First and Second SARS-CoV2 Booster Vaccines during the Delta and Omicron Pandemic Waves in Hungary (HUN-VE 2 Study). medRxiv. Published online 2022. doi:10.1101/2022.03.27.22273000

86. Barda N, Canetti M, Gilboa M, et al. Comparing immunogenicity and efficacy of two different mRNA-based COVID-19 vaccines as a fourth dose; six-month follow-up, Israel, 27 December 2021 to 24 July 2022. Eurosurveillance. 2022;27(39). doi:10.2807/1560-7917.ES.2022.27.39.2200701

87. Fabiani M, Mateo-Urdiales A, Sacco C, et al. Relative effectiveness of a 2nd booster dose of COVID-19 mRNA vaccine up to four months post administration in individuals aged 80 years or more in Italy: A retrospective matched cohort study. Vaccine. 2023;41(1):76–84. doi:10.1016/j.vaccine.2022.11.013

88. Kim YY, Choe YJ, Kim J, et al. Effectiveness of Second mRNA COVID-19 Booster Vaccine in Immunocompromised Persons and Long-Term Care Facility Residents. Emerg Infect Dis. 2022;28(11):2165–2170. doi:10.3201/eid2811.220918

89. McConeghy KW, White EM, Blackman C, et al. Effectiveness of a Second COVID-19 Vaccine Booster Dose Against Infection, Hospitalization, or Death Among Nursing Home Residents - 19 States, March 29-July 25, 2022. MMWR Morb Mortal Wkly Rep. 2022;71(39):1235-1238. doi:10.15585/mmwr.mm7139a2

90. Tan CY, Chiew CJ, Lee VJ, Ong B, Lye DC, Tan KB. Comparative effectiveness of 3 or 4 doses of mRNA and inactivated whole-virus vaccines against COVID-19 infection, hospitalization and severe outcomes among elderly in Singapore. Lancet Reg Health West Pac. 2022;29:100654. doi:10.1016/j.lanwpc.2022.100654

91. Tenforde MW. Early Estimates of Bivalent mRNA Vaccine Effectiveness in Preventing COVID-19–Associated Emergency Department or Urgent Care Encounters and Hospitalizations Among Immunocompetent Adults — VISION Network, Nine States, September–November 2022. MMWR Morb Mortal Wkly Rep. 2023;71. doi:10.15585/mmwr.mm7153a1

92. Shrestha NK, Burke PC, Nowacki AS, Simon JF, Hagen A, Gordon SM. Effectiveness of the Coronavirus Disease 2019 (COVID-19) Bivalent Vaccine. Published online March 22, 2023:2022.12.17.22283625. doi:10.1101/2022.12.17.22283625

93. Link-Gelles R. Effectiveness of Bivalent mRNA Vaccines in Preventing Symptomatic SARS-CoV-2 Infection — Increasing Community Access to Testing Program, United States, September–November 2022. Morb Mortal Wkly Rep. 2022;71. doi:10.15585/mmwr.mm7148e1

94. Link-Gelles R. Early Estimates of Bivalent mRNA Booster Dose Vaccine Effectiveness in Preventing Symptomatic SARS-CoV-2 Infection Attributable to Omicron BA.5– and XBB/XBB.1.5–Related Sublineages Among Immunocompetent Adults — Increasing Community Access to Testing Program, United States, December 2022–January 2023. Morb Mortal Wkly Rep. 2023;72. doi:10.15585/mmwr.mm7205e1

95. Fabiani M, Mateo-Urdiales A, Sacco C, et al. Protection against severe COVID-19 after second booster dose of adapted bivalent (original/Omicron BA.4-5) mRNA vaccine in persons ≥ 60 years, by time since infection, Italy, 12 September to 11 December 2022. Eurosurveillance. 2023;28(8):2300105. doi:10.2807/1560-7917.ES.2023.28.8.2300105

96. Huiberts AJ, Gier B de, Hoeve CE, et al. Effectiveness of bivalent mRNA booster vaccination against SARS-CoV-2 Omicron infection, the Netherlands, September to December 2022. Eurosurveillance. 2023;28(7):2300087. doi:10.2807/1560-7917.ES.2023.28.7.2300087

