## Supplementary Methods and Figures for "Design and Analysis Heterogeneity in Observational Studies of COVID-19 Booster Effectiveness: A Review and Case Study"

**Methods A1**: Search terms for PubMed Search

Date searched: 1/1/2023

Number of results: 217

Date filter: January 1, 2021 to present

Language filter: English

Search Blocks

1. "covid-19"[Text Word] OR "COVID19"[Text Word] OR "COVID-19"[Supplementary Concept] OR "SARS-CoV-2"[Text Word] OR "SARS-CoV2"[Text Word] OR "severe acute respiratory syndrome coronavirus 2"[Supplementary Concept] OR "severe acute respiratory syndrome coronavirus 2"[Text Word] OR "2019-nCoV"[Text Word] OR "2019nCoV"[Text Word] OR "coronavirus"[Text Word] OR "coronavirus"[MeSH Terms]
2. "booster"[Title] OR "boosters"[Title] OR "third dose"[Title] OR "fourth dose"[Title] OR "3 doses"[Title] OR "4 doses"[Title]
3. "effectiveness"[Title/Abstract] OR "effective"[Title/Abstract] OR “effect”[Title] OR “effects”[Title] OR "odds"[Title] OR "protection"[Title] OR “association”[Title] OR “comparison”[Title] OR “comparing”[Title]
4. "infection"[Title/Abstract] OR "hospitalization"[Title/Abstract] OR "COVID-19 outcomes"[Title/Abstract] OR "COVID19 outcomes"[Title/Abstract] OR "covid 19 outcomes"[Title/Abstract] OR "severe COVID-19"[Title/Abstract] OR "severe COVID19"[Title/Abstract] OR "severe covid 19"[Title/Abstract] OR "confirmed COVID-19"[Title/Abstract] OR "confirmed COVID19"[Title/Abstract] OR "confirmed covid 19"[Title/Abstract] OR “ICU”[Title/Abstract] OR “intensive care”[Title/Abstract] OR “mortality”[Title/Abstract] OR “death”[Title/Abstract]

**Methods A2**: Search terms for Embase Search

Date Searched: 1/1/2023

Number of Results: 96

Date filter: January 1, 2021 to present

Language filter: English

Other filters applied: exclude MedLine (only Embase and preprints), only articles and preprints (exclude conference abstracts and reviews)

Search Blocks:

1. 'covid 19':ti,ab OR covid19:ti,ab OR 'sars cov 2':ti,ab OR 'sars cov2':ti,ab OR 'severe acute respiratory syndrome coronavirus 2':ti,ab OR '2019 ncov':ti,ab OR 2019ncov:ti,ab OR coronavirus:ti,ab OR 'coronavirinae'/exp OR 'coronavirus disease 2019'/exp OR covid:ti,ab
2. 'booster':ti OR 'boosters':ti OR 'third dose':ti OR 'fourth dose':ti OR '3 doses':ti OR '4 doses':ti
3. 'effectiveness':ti,ab OR 'effective':ti,ab OR 'effect':ti OR 'effects':ti OR 'odds':ti OR 'protection':ti OR 'association':ti OR 'comparison':ti OR 'comparing':ti
4. "infection":ti,ab OR "hospitalization":ti,ab OR "COVID-19 outcomes":ti,ab OR "COVID19 outcomes":ti,ab OR "covid 19 outcomes":ti,ab OR "severe COVID-19":ti,ab OR "severe COVID19":ti,ab OR "severe covid 19":ti,ab OR "confirmed COVID-19":ti,ab OR "confirmed COVID19":ti,ab OR "confirmed covid 19":ti,ab OR "ICU":ti,ab OR "intensive care":ti,ab OR "mortality":ti,ab OR "death":ti,ab

**Methods A3:** Test-negative methods

**Study Population**

Our study population for the main analysis consisted of 65,992 individuals (75,645 individuals for the two or more booster secondary analysis) after exclusion criteria (from 115,660 individuals before exclusion criteria) who received two or three doses of the BNT162b2 (Pfizer) or mRNA-1273 (Moderna) vaccines and were tested for (including both positive and negative results) or diagnosed with COVID-19 at Michigan medicine between October 1, 2021 and December 31, 2022. We allowed each individual to contribute up to one test per quarter, treating tests as the unit of observation in models. Any tests taken after a previous positive test or documented infection were excluded. We also restricted the data to complete cases of all the covariates used in analysis, which are given below.

**Exposures**

We used the same exposures as in the cohort analysis.

**Outcomes**

We considered the same outcomes as in the cohort analysis.

**Statistical Analysis**

For each combination of study population and outcome, we fit several models using a variety of different adjustment, matching, or propensity score strategies to the data. As a secondary analysis, we, also for each combination of study population and outcome, fit unadjusted and unadjusted models (not including matching or propensity scores) using different quarters of study start dates (only including tests that occurred during or after the study start quarter) between Q4 2021 and Q4 2022.

Models in the test-negative analysis used logistic regression, formulated as $\text{logit}\left[ P\left( Y_{\text{outcome}}=1 | I\left( \text{Vaccine} \right),\text{Covariates} \right) \right]=\beta_{0}+\beta_{\text{Vax}}I\left( \text{Vaccine} \right)+\beta_{\text{Cov}}\text{Covariates}$ (see Methods A4 for definitions of vaccination and outcome indicators). Some models utilizing matching used conditional logistic regression, an extension of logistic regression with matching stratum-specific intercepts, formulated as $\text{logit}\left[ P\left( Y_{\text{outcome}}=1 | I\left( \text{Vaccine} \right),\text{Covariates} \right) \right]=\beta_{0,j|i}+\beta_{\text{Vax}}I\left( \text{Vaccine} \right)+\beta_{\text{Cov}}\text{Covariates}$. VE was calculated by exponentiating the coefficient for vaccination, giving the odds ratio for the outcome for those boosted compared to those in the previous stage of vaccination, then subtracting it from one, i.e., $\hat{\text{VE}}=1-e^{\hat{\beta}_{\mathrm{Vax}}}=1-{\hat{\text{OR}}}_{\text{Vax:Outcome}}$. For infection models, cases were considered as individuals who tested positive or were diagnosed with COVID-19 and controls those who tested negative.

For severe disease models, the choice of control group is multifaceted. To demonstrate, we can represent the study population as a 2x2 table of severe disease versus test results:

|  | Severe Disease | No Severe Disease |
| --- | --- | --- |
| Test-Positive | Case | Potential Control A |
| Test-Negative | Potential Control B | Potential Control C |

We can express each of these control groups in terms of a 2x2 table (also including a potential control group 4 that was not examined in our analysis):

| Control Group | Table 4 Potential Controls |
| --- | --- |
| 1 - Test-negative | B, C |
| 2 - Non-hospitalized | A, C |
| 3 - Test-positive non-hospitalized | A |
| 4 - Test-negative non-hospitalized | C |

We considered control groups 1, 2, and 3 shown above. In sum, cases were considered those with severe disease defined above, and we fit models with different three choices of controls: those who tested negative (control group 1), those who had no severe disease regardless of test results (control group 2), and those who tested positive but had no severe disease (control group 3). For the last model, final VE was calculated as $\hat{\text{VE}}=1-{\hat{\text{OR}}}_{\text{Vax:Infection}}*{\hat{\text{OR}}}_{\text{Vax:Severe Disease}}$, where the odds ratio for infection is obtained from a model using the same design otherwise with infection and not severe disease as the outcome variable in logistic regression.

We adjusted for the same set of covariates as done in the cohort models, except with the addition of quarter of test.

Unadjusted logistic regression and adjusted logistic regression for the covariates were fit to the data. In addition, we fit models using the same matching and propensity score methods used with the cohort data (exact matching, propensity score caliper matching, models adjusted for propensity for boosting, and models with IPW for propensity for boosting) and explained further in Table A1 (except note that unconditional logistic regression was used in lieu of unstratified Cox regression, and conditional logistic regression was used in lieu of stratified Cox regression).

**Methods A4**: Definitions of terms in models (vaccination and outcome indicators)

Exposures:

For first booster models:

$$I\left( \text{Vaccine} \right)=\left\{ \begin{aligned} 0, fully vaccinated \\ 1, \mathrm{boosted} \end{aligned} \right.$$

For second booster models:

$$I\left( \text{Vaccine} \right)=\left\{ \begin{aligned} 0, once boosted \\ 1, boosted twice or more \end{aligned} \right.$$

Outcomes:

For infection models:

$$I\left( \text{Outcome} \right)=\left\{ \begin{aligned} 0, tested negative \\ 1, tested positive \end{aligned} \right.$$

For severe disease models:

$$I\left( \text{Outcome} \right)=\left\{ \begin{aligned} 0, no severe disease \\ 1, severe disease \end{aligned} \right.$$

**Supplementary Figures**

**Literature Review**

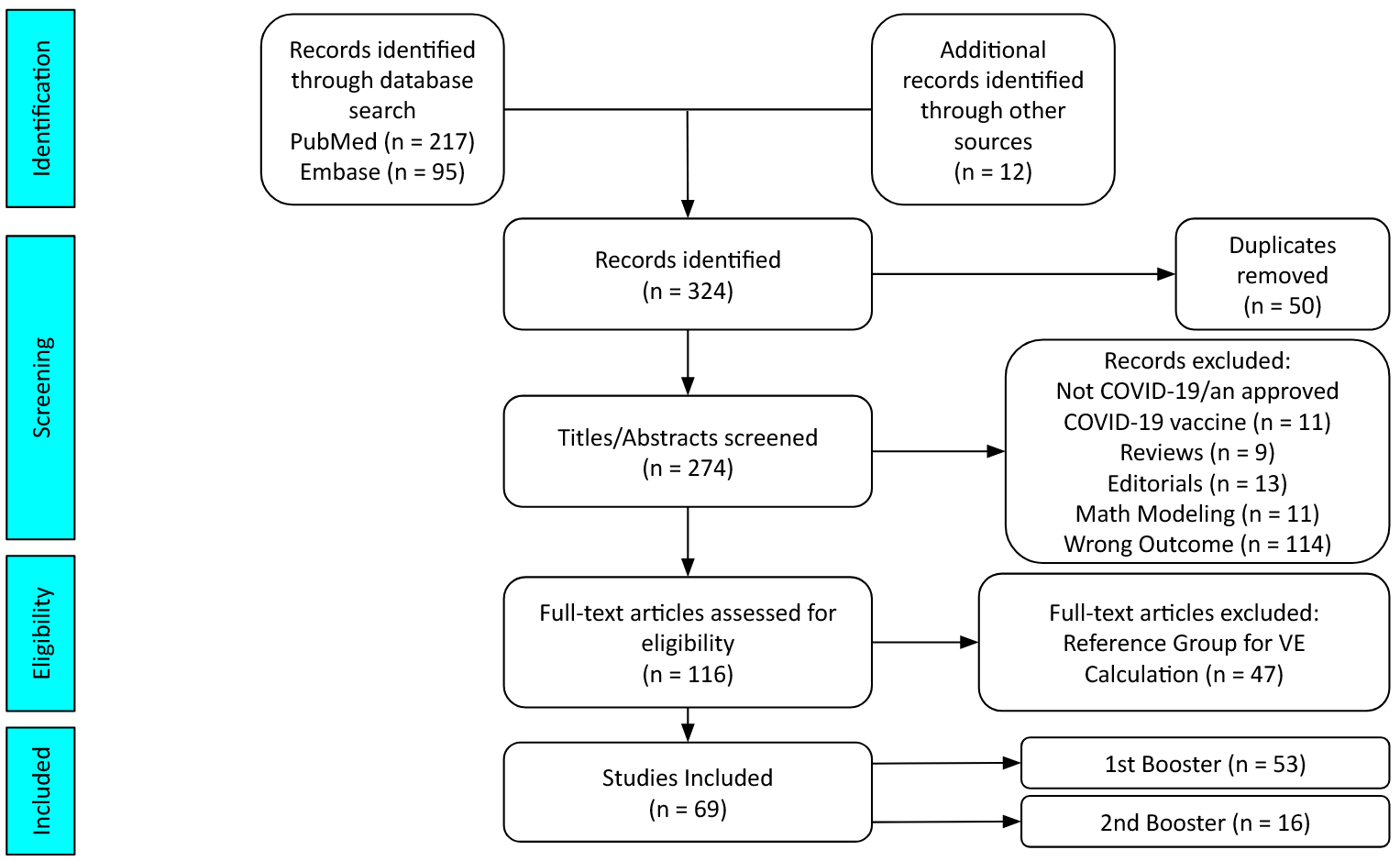

**Figure A1**: PRISMA flow diagram for the selection process of studies in the literature review.

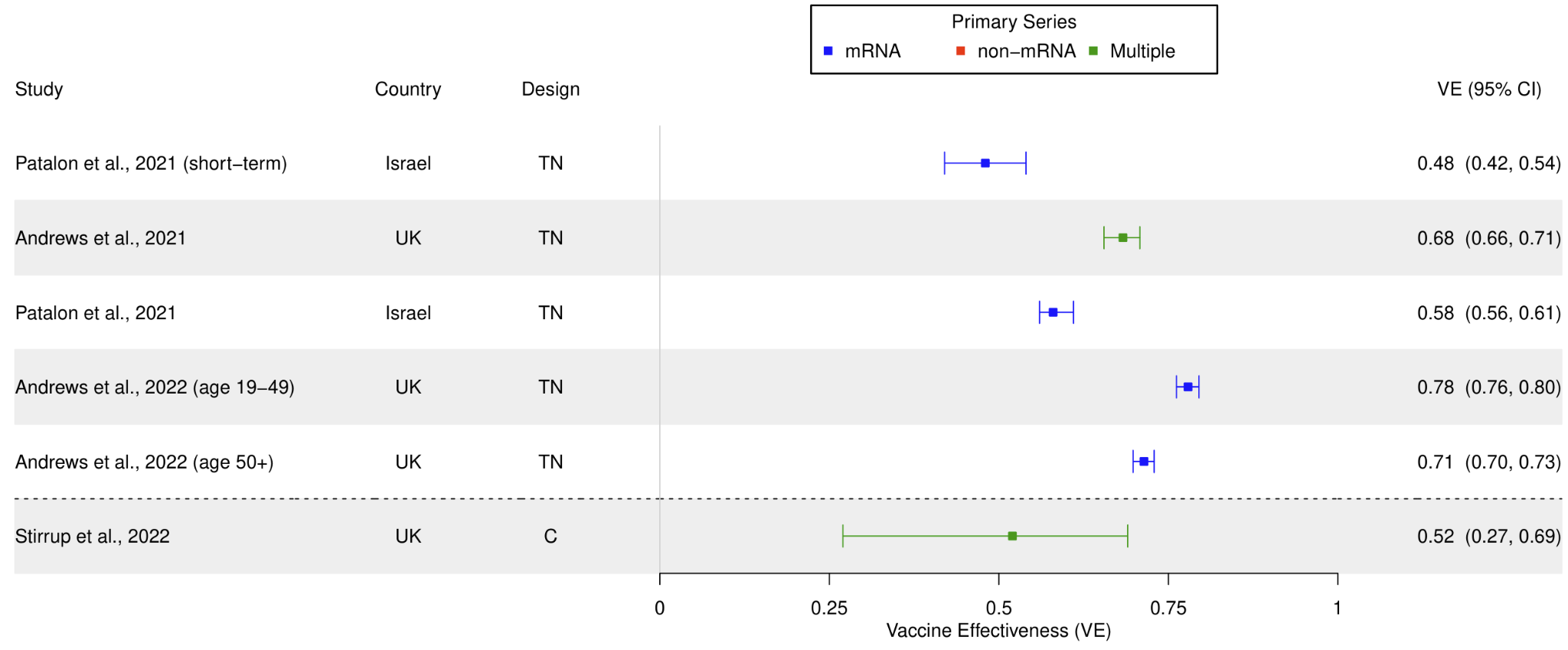

**Figure A2**: First booster VE compared to primary series against infection between 1 and 2 weeks post-booster vaccination. Studies are sorted in chronological order of last month included in the study period (with publication date being used to break ties). The dotted line indicates a division between studies ending in 2021 and ending in 2022 (noting that Omicron became the dominant variant in many countries around the end of 2021).

Abbreviations: TN, test-negative; MTN, matched test-negative; CC, case-control (non-test-negative); MCC, matched case-control (non-test-negative); C, cohort; MC, matched cohort.

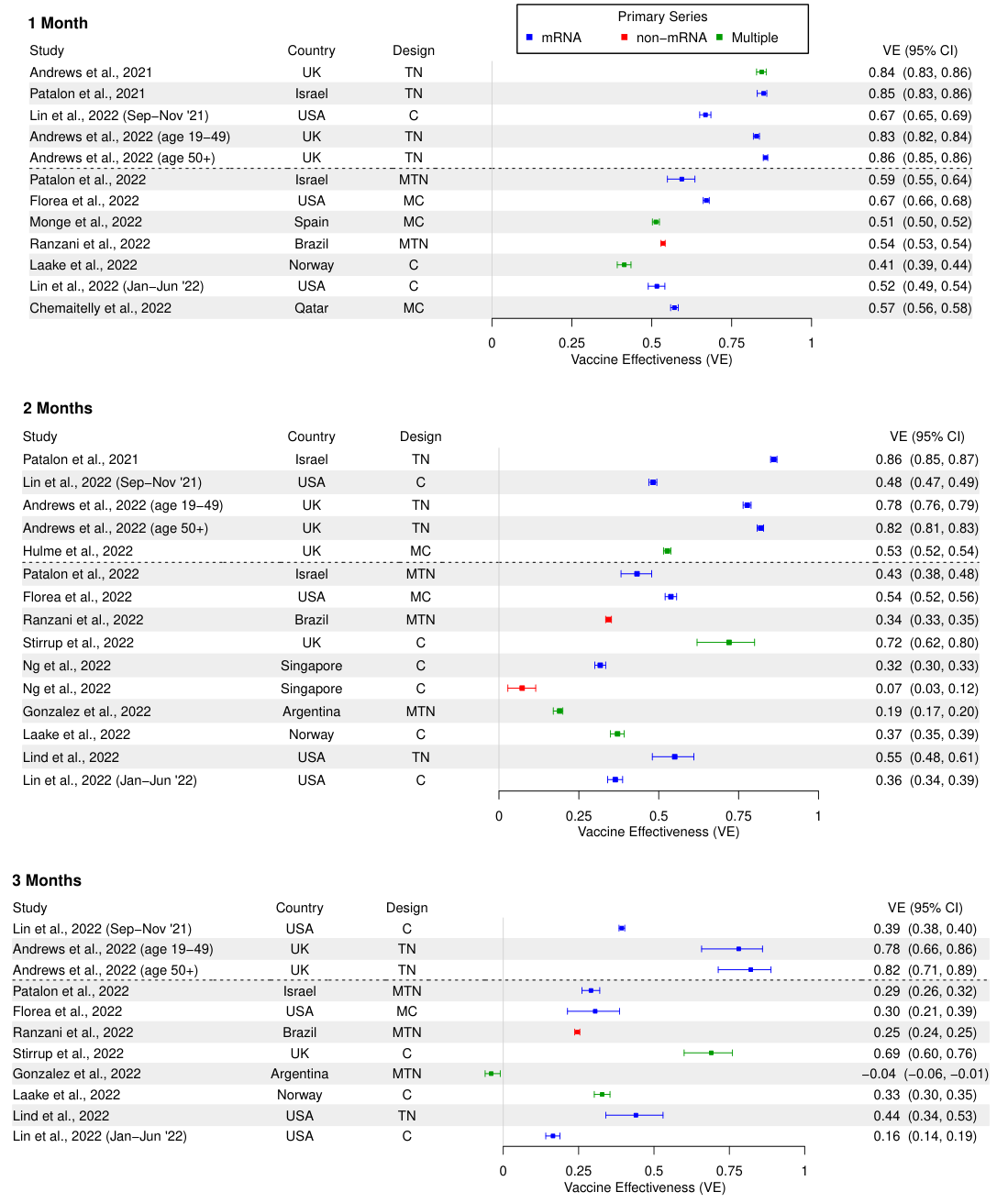

**Figure A3**: First booster VE compared to primary series against infection at 1, 2, and 3 months after booster vaccination. Studies are sorted in chronological order of last month included in the study period (with publication date being used to break ties). The dotted line indicates a division between studies ending in 2021 and ending in 2022 (noting that Omicron became the dominant variant in many countries around the end of 2021).

Abbreviations: TN, test-negative; MTN, matched test-negative; CC, case-control (non-test-negative); MCC, matched case-control (non-test-negative); C, cohort; MC, matched cohort.

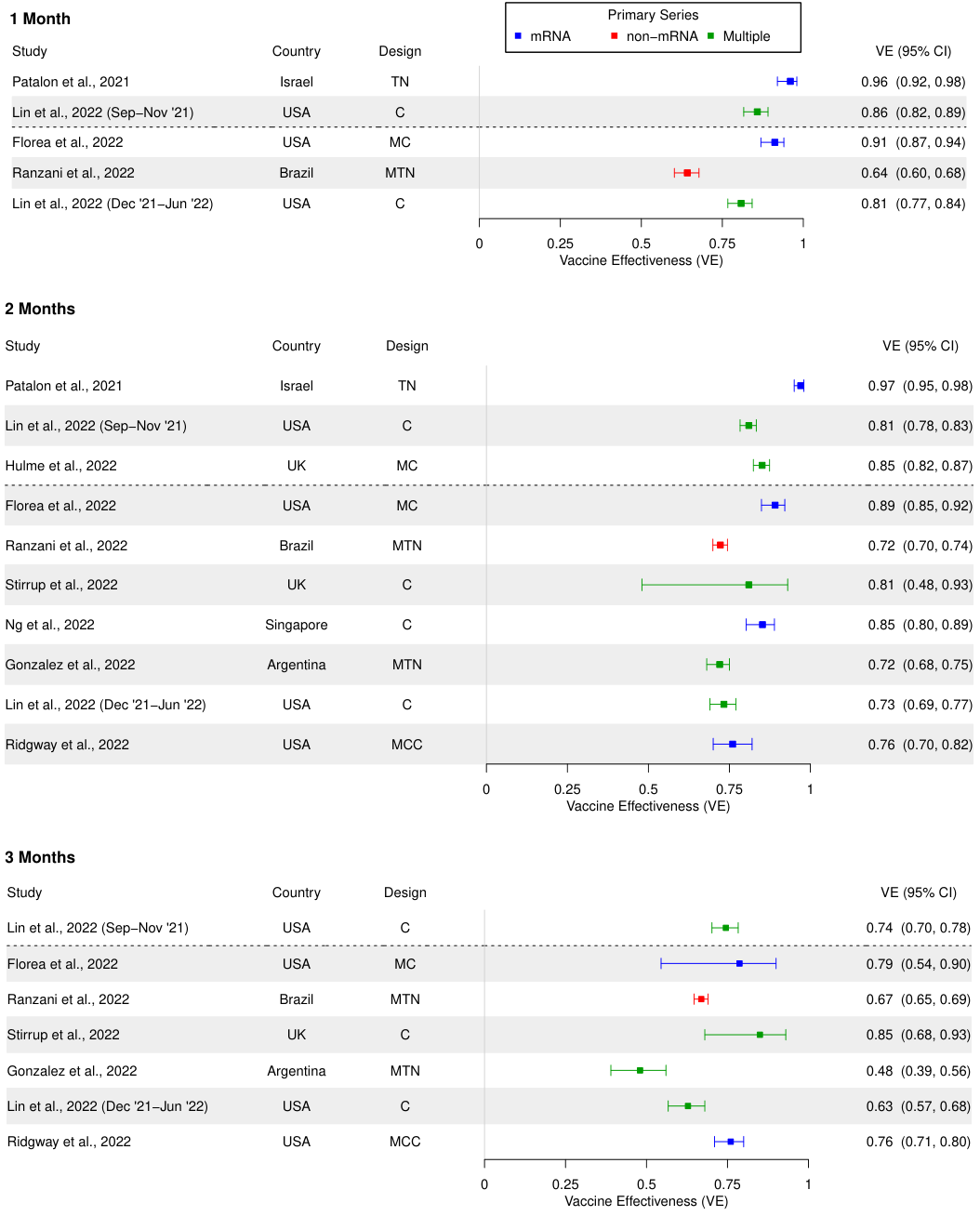

**Figure A4**: First booster VE compared to primary series against hospitalization or severe disease outcomes at 1, 2, and 3 months after booster vaccination. Studies are sorted in chronological order of last month included in the study period (with publication date being used to break ties). The dotted line indicates a division between studies ending in 2021 and ending in 2022 (noting that Omicron became the dominant variant in many countries around the end of 2021).

Abbreviations: TN, test-negative; MTN, matched test-negative; CC, case-control (non-test-negative); MCC, matched case-control (non-test-negative); C, cohort; MC, matched cohort.

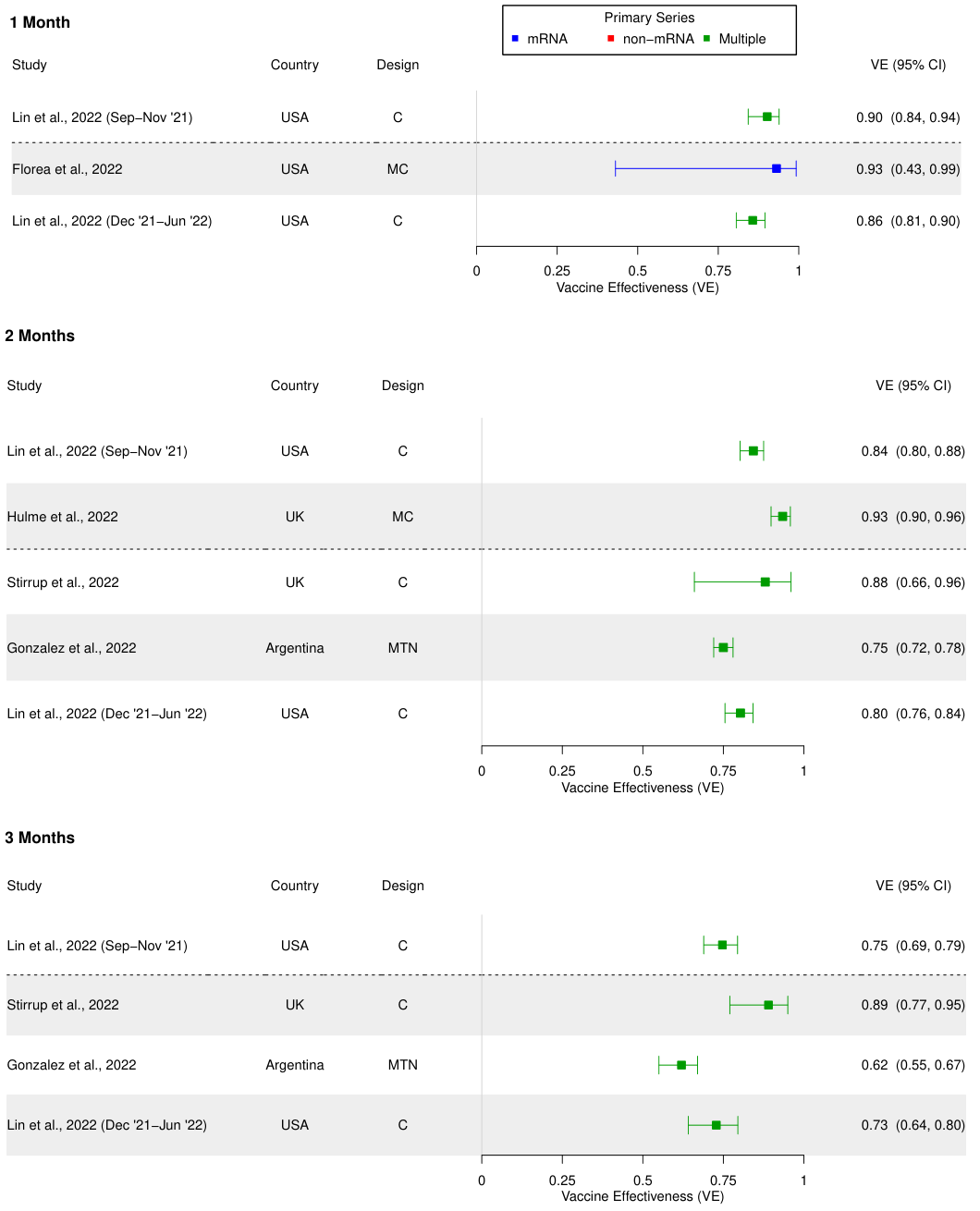

**Figure A5**: First booster VE compared to primary series against mortality at 1 month after booster vaccination. Studies are sorted in chronological order of last month included in the study period (with publication date being used to break ties). The dotted line indicates a division between studies ending in 2021 and ending in 2022 (noting that Omicron became the dominant variant in many countries around the end of 2021).

Abbreviations: TN, test-negative; MTN, matched test-negative; CC, case-control (non-test-negative); MCC, matched case-control (non-test-negative); C, cohort; MC, matched cohort.

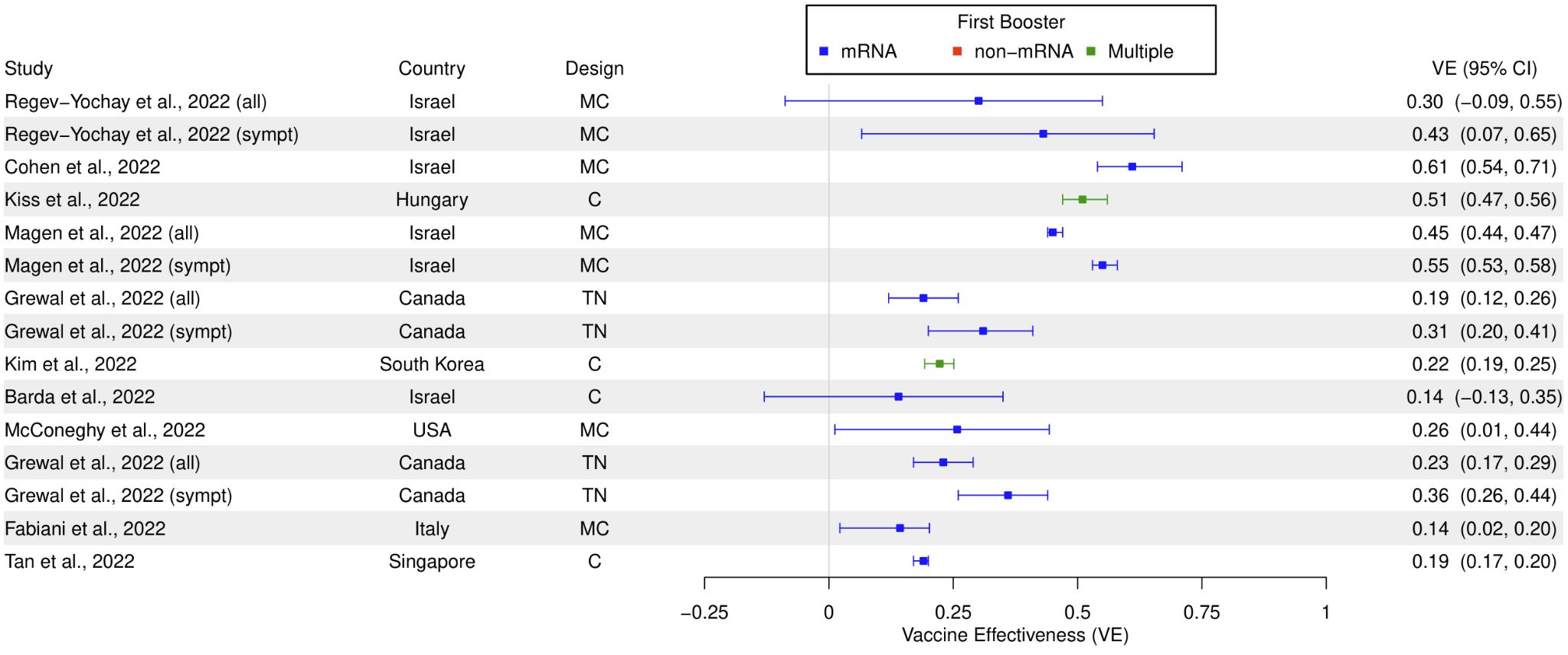

**Figure A6**: Second booster VE compared to first booster against infection unstratified by time since booster vaccination. Studies are sorted in chronological order of last month included in the study period (with publication date being used to break ties). Both symptomatic and overall results are included for studies that produced estimates for both, indicated by “(sympt)” and “(all)”, respectively.

Abbreviations: TN, test-negative; MTN, matched test-negative; CC, case-control (non-test-negative); MCC, matched case-control (non-test-negative); C, cohort; MC, matched cohort.

**First Booster vs. Primary Series**

**Cohort Infection and Severe Disease – First Booster vs. Primary Series**

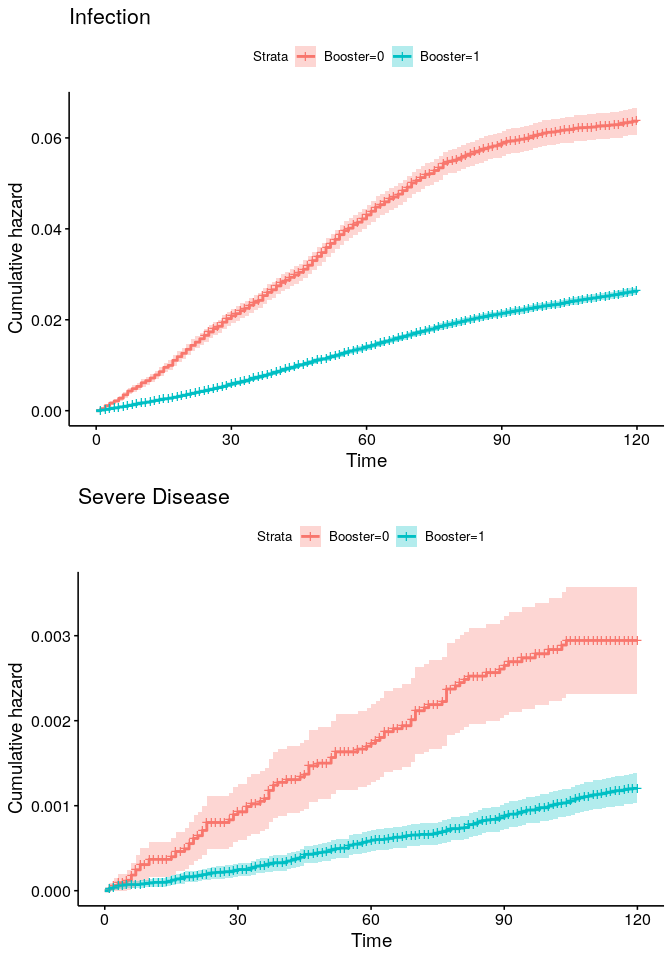

**Figure A7:** Kaplan-Meier curves for first booster/third dose vs full vaccination/second dose VE against infection and severe disease during a study period of October 1, 2021 to December 31, 2022

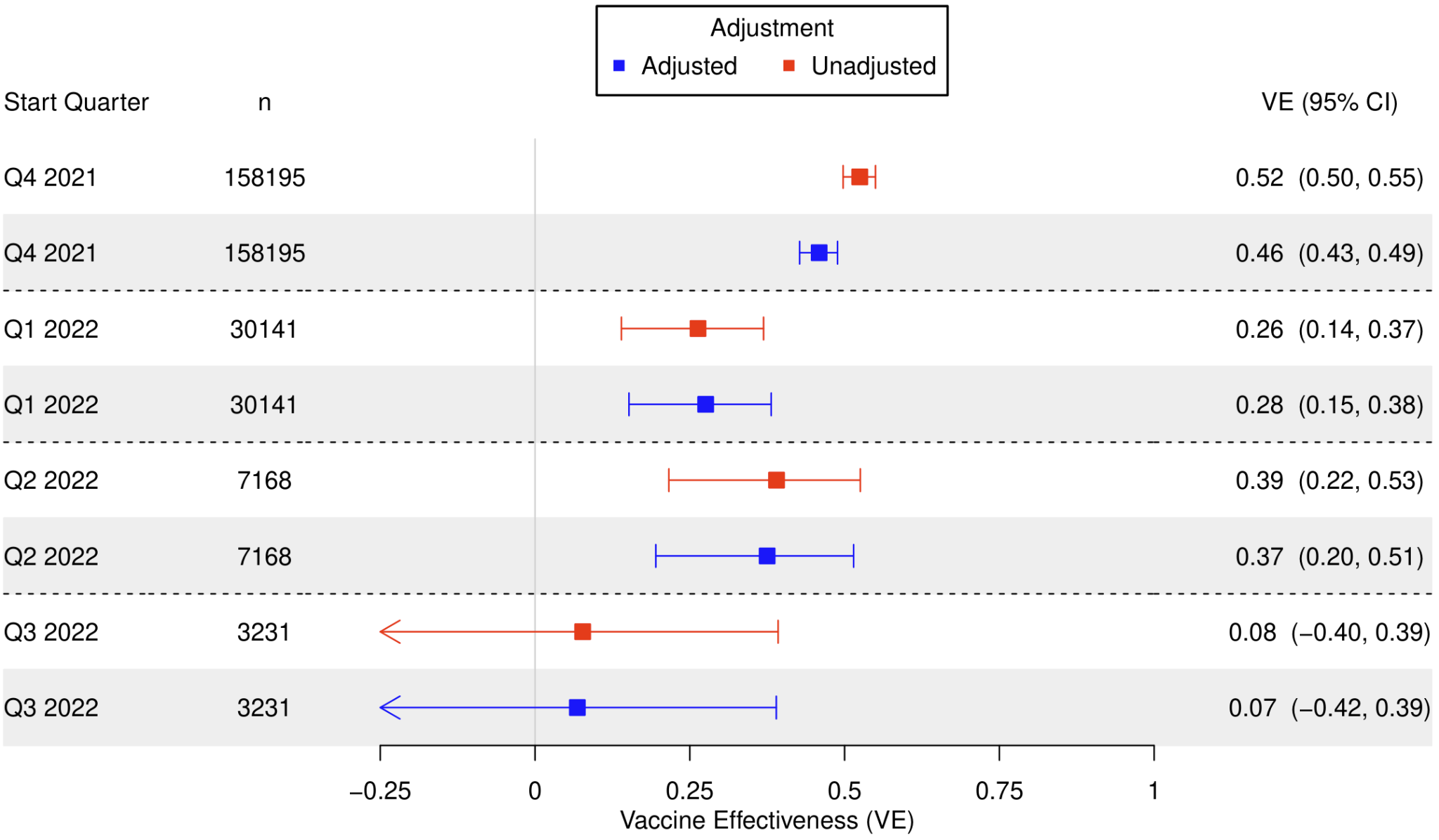

**Figure A8**: Cohort first booster/third dose vs full vaccination/second dose VE against infection, with various study start (index) dates and with an end date of Dec 31, 2022.

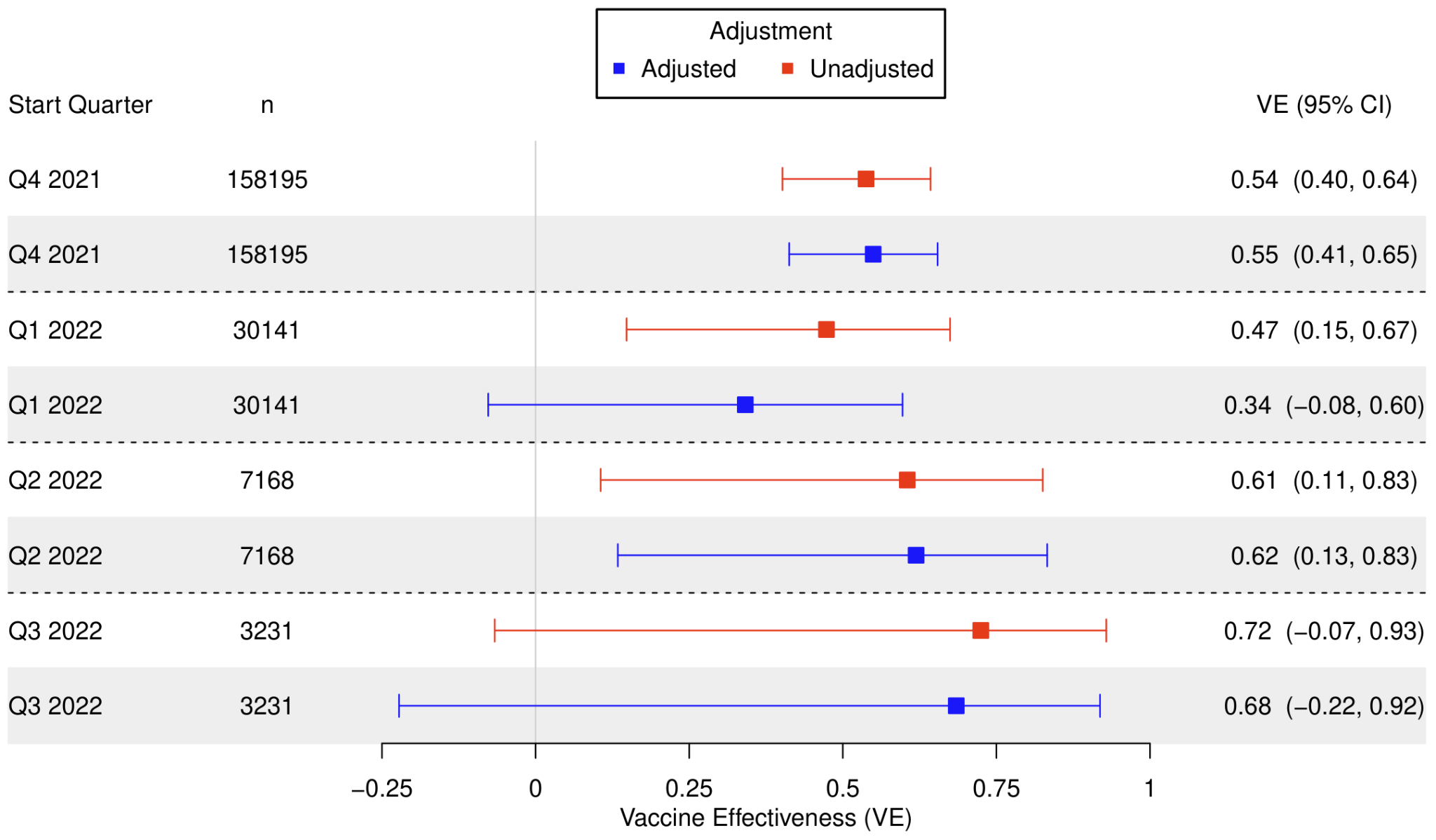

**Figure A9**: Cohort first booster/third dose vs full vaccination/second dose VE against severe disease, with various study start (index) dates and with an end date of Dec 31, 2022.

**Test-Negative Infection – First Booster vs. Primary Series**

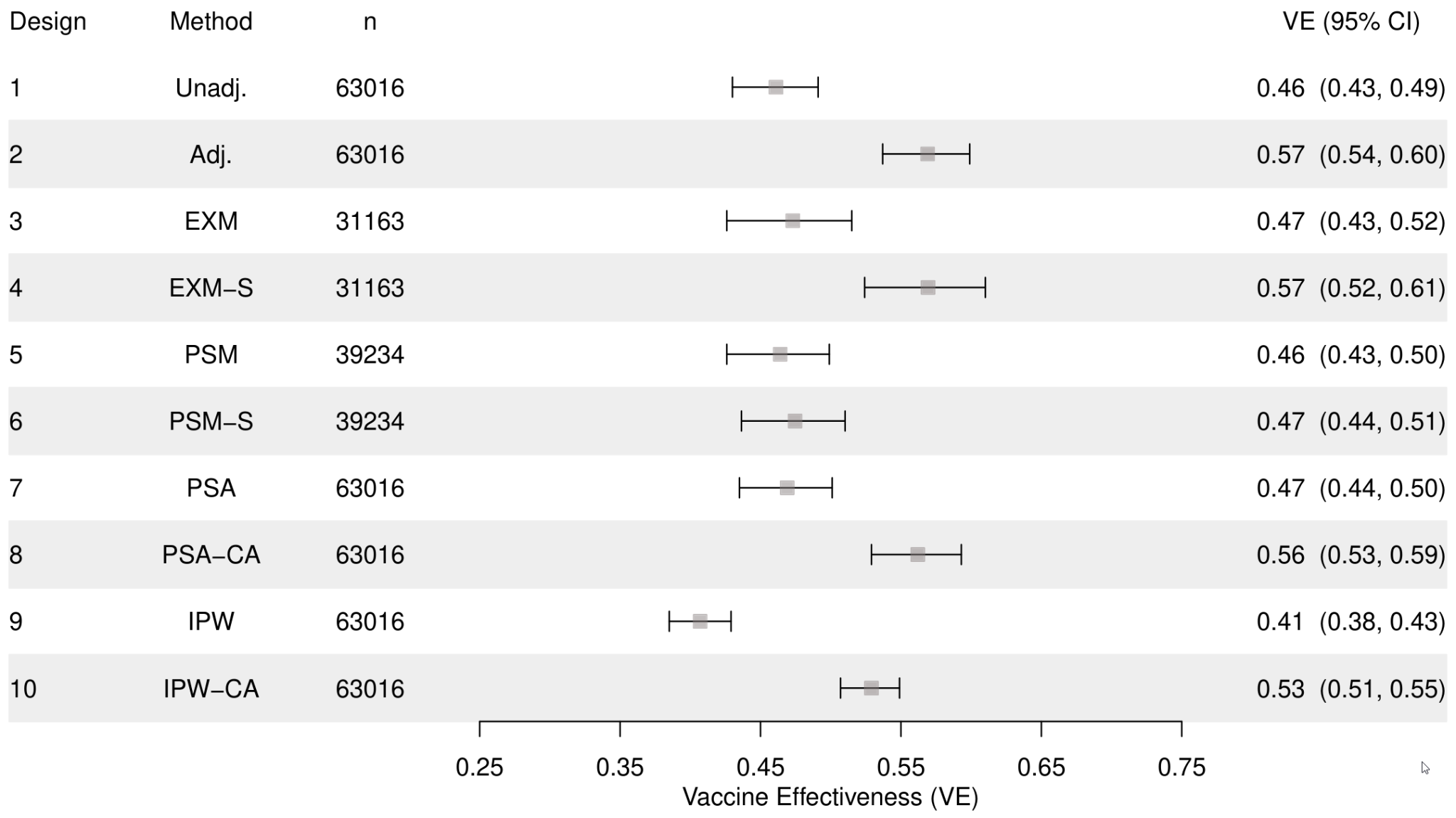

**Figure A10**: Test-negative first booster/third dose up to 2 quarters since vaccination vs full vaccination/second dose VE against infection, with various matching designs during a study period of October 1, 2021 to December 31, 2022.

Abbreviations: Unadj., unadjusted; Covar. Adj., adjusted for covariates; Exact, exact matching; Exact Strat., exact matching accounting for matching strata in conditional logistic regression; Prop. Cal., propensity score matching with a 0.2 caliper; Prop. Cal. Strat, propensity score matching with a 0.2 caliper accounting for matching strata in conditional logistic regression; Prop. Adj.; adjustment for propensity score; Prop. + Covar. Adj., adjustment for propensity score and for covariates; IPW, inverse probability weighting of propensity score; IPW Covar. Adj., inverse probability weighting of propensity score also adjusting for covariates.

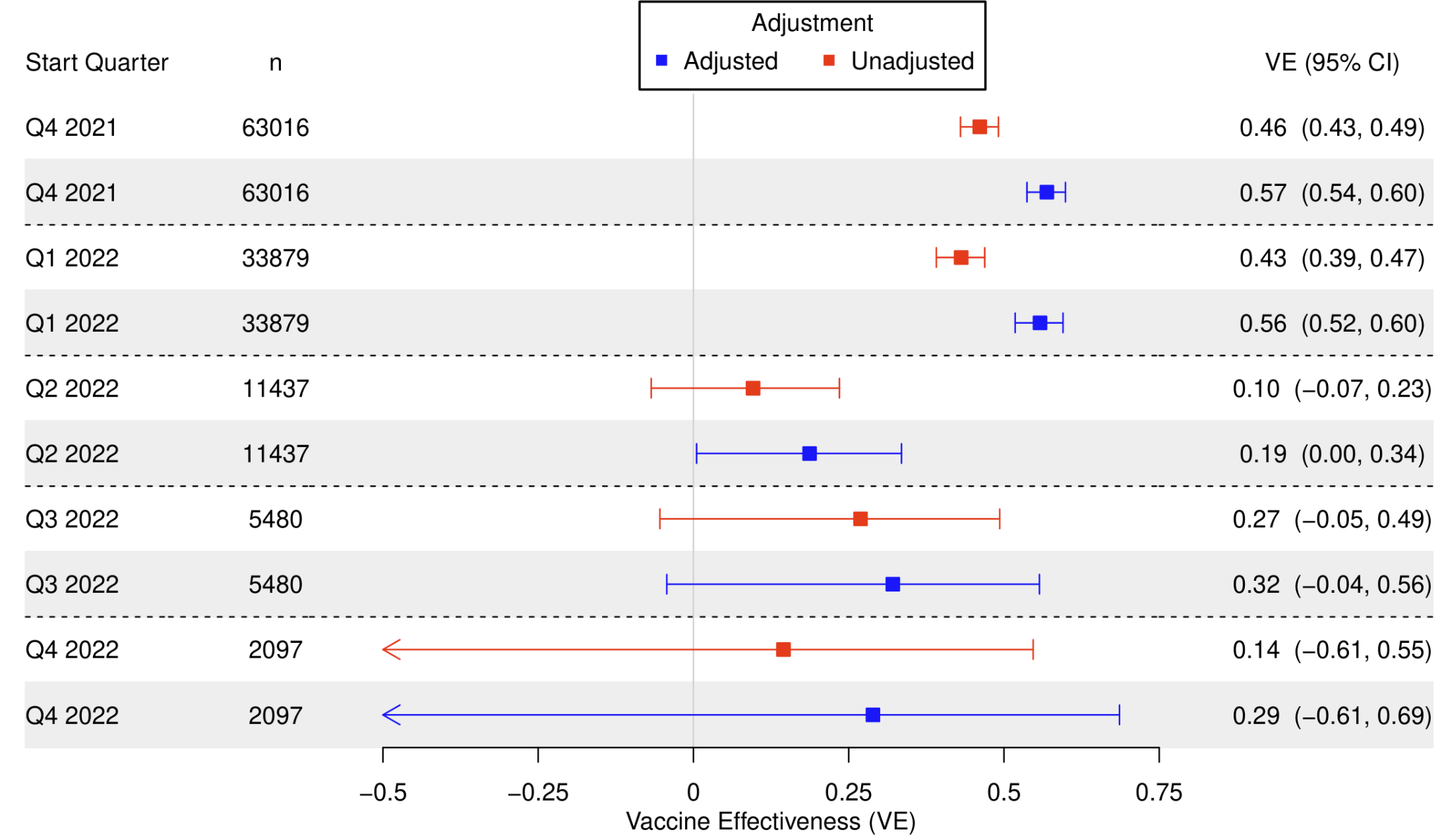

**Figure A11**: Test-negative first booster/third dose up to 2 quarters since vaccination vs full vaccination/second dose VE against infection, with various study start (index) dates and with an end quarter of Q4 2022.

**Test-Negative Severe Disease – First Booster vs. Primary Series**

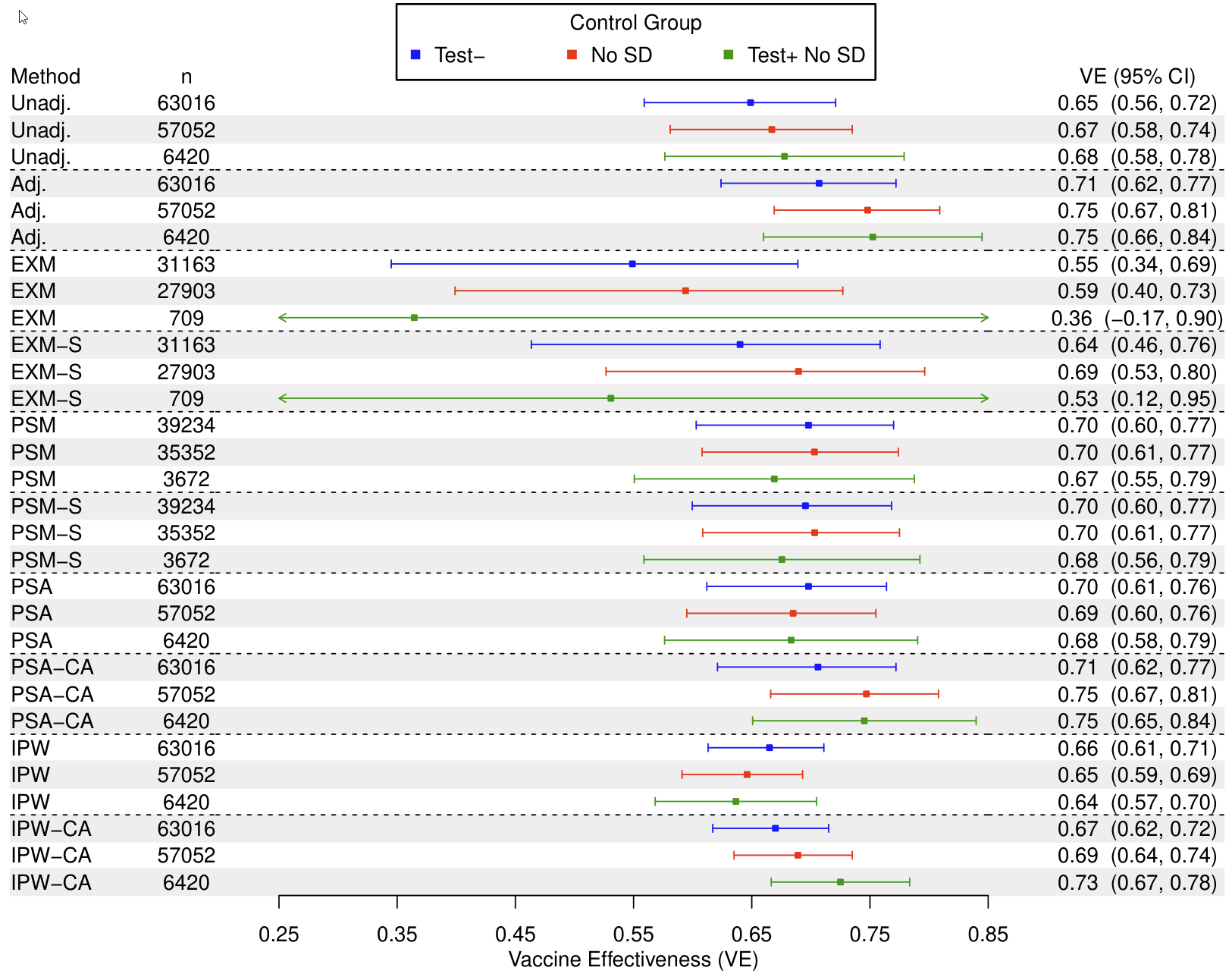

**Figure A12**: Test-negative first booster/third dose up to 2 quarters since vaccination vs full vaccination/second dose VE against severe disease, with various matching designs during a study period of October 1, 2021 to December 31, 2022.

Control Groups: (1) Test-, test-negatives; (2) No SD, no severe disease; (3) Test+ No SD, test-positives without severe disease.

Abbreviations: Unadj., unadjusted; Adj., adjusted for covariates; EXM, exact matching; EXM-S, exact matching accounting for matching strata in conditional logistic regression; PSM, propensity score matching with a 0.2 caliper; PSM-S, propensity score matching with a 0.2 caliper accounting for matching strata in conditional logistic regression; PSA; adjustment for propensity score; PSA-CA, adjustment for propensity score and for covariates; IPW, inverse probability weighting of propensity score; IPW-CA, inverse probability weighting of propensity score also adjusting for covariates.

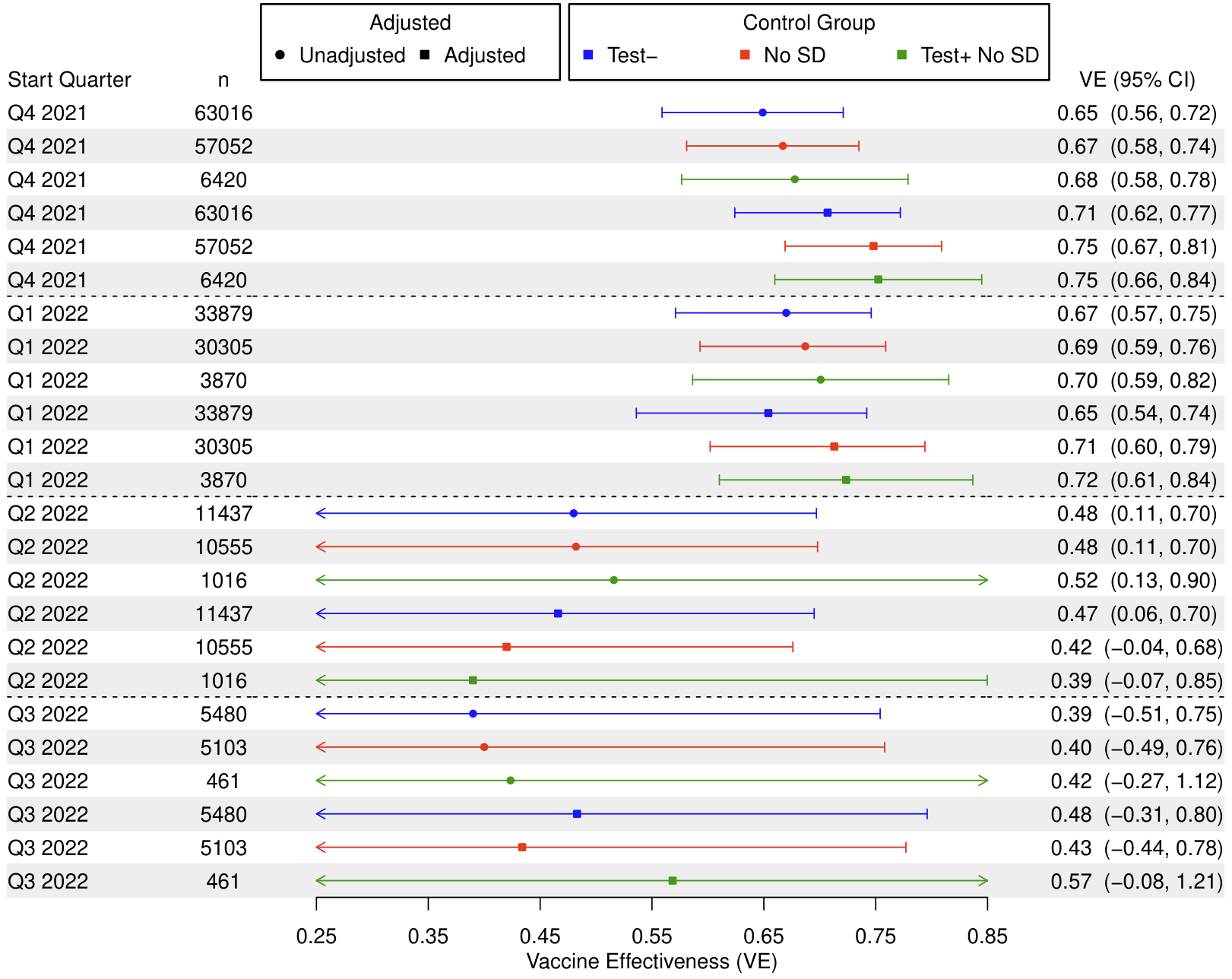

**Figure A13**: Test-negative first booster/third dose up to 2 quarters since vaccination vs full vaccination/second dose VE against severe disease, with various study start (index) dates and choices of control group and with an end quarter of Q4 2022.

Control Groups: (1) Test-, test-negatives; (2) No SD, no severe disease; (3) Test+ No SD, test-positives without severe disease.

**One or More Boosters vs. Primary Series**

**Cohort Infection – One or More Boosters vs. Primary Series**

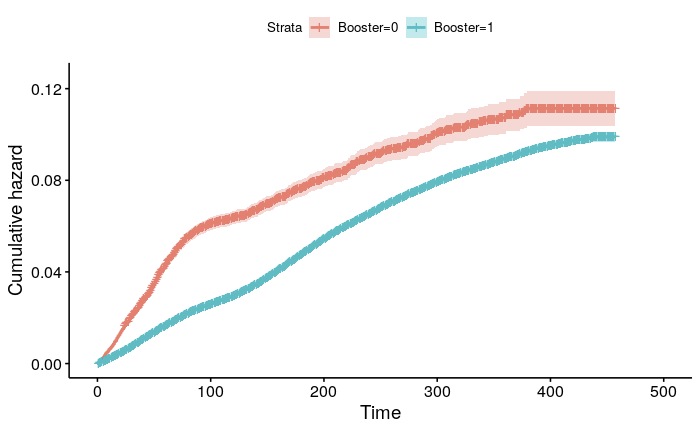

**Figure A14**: Kaplan-Meier curve for cohort boosted (3 or more doses) vs full vaccination/second dose VE against infection during a study period of October 1, 2021 to December 31, 2022.

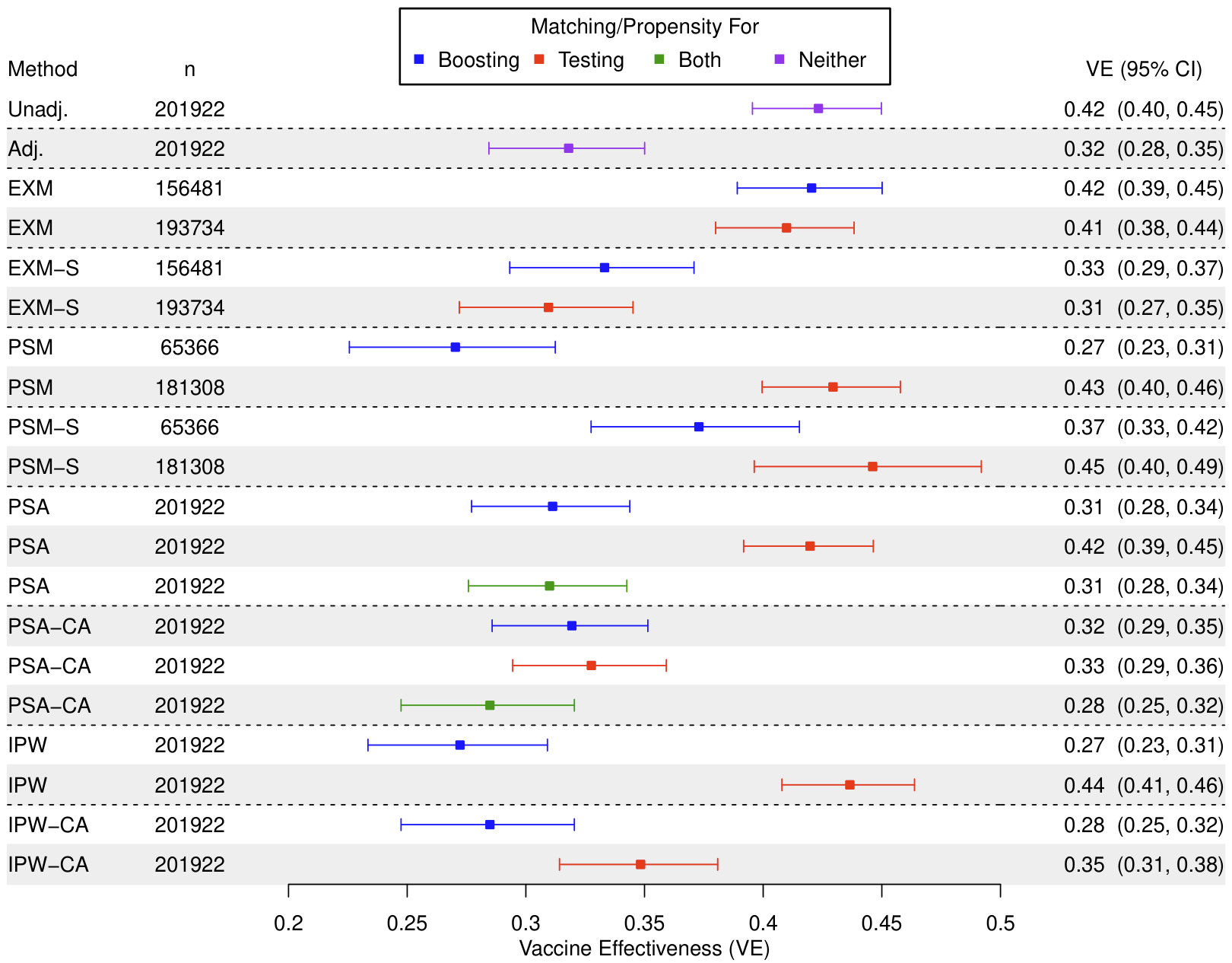

**Figure A15**: Cohort boosted (3 or more doses) vs full vaccination/second dose VE against infection, with various matching designs during a study period of October 1, 2021 to December 31, 2022.

Abbreviations: Unadj., unadjusted; Adj., adjusted for covariates; EXM, exact matching; EXM-S, exact matching with matching strata stratification in Cox regression; PSM, propensity score matching with a 0.2 caliper; PSM-S, propensity score matching with a 0.2 caliper and matching strata stratification in Cox regression; PSA; adjustment for propensity score; PSA-CA, adjustment for propensity score and for covariates; IPW, inverse probability weighting of propensity score; IPW-CA, inverse probability weighting of propensity score also adjusting for covariates.

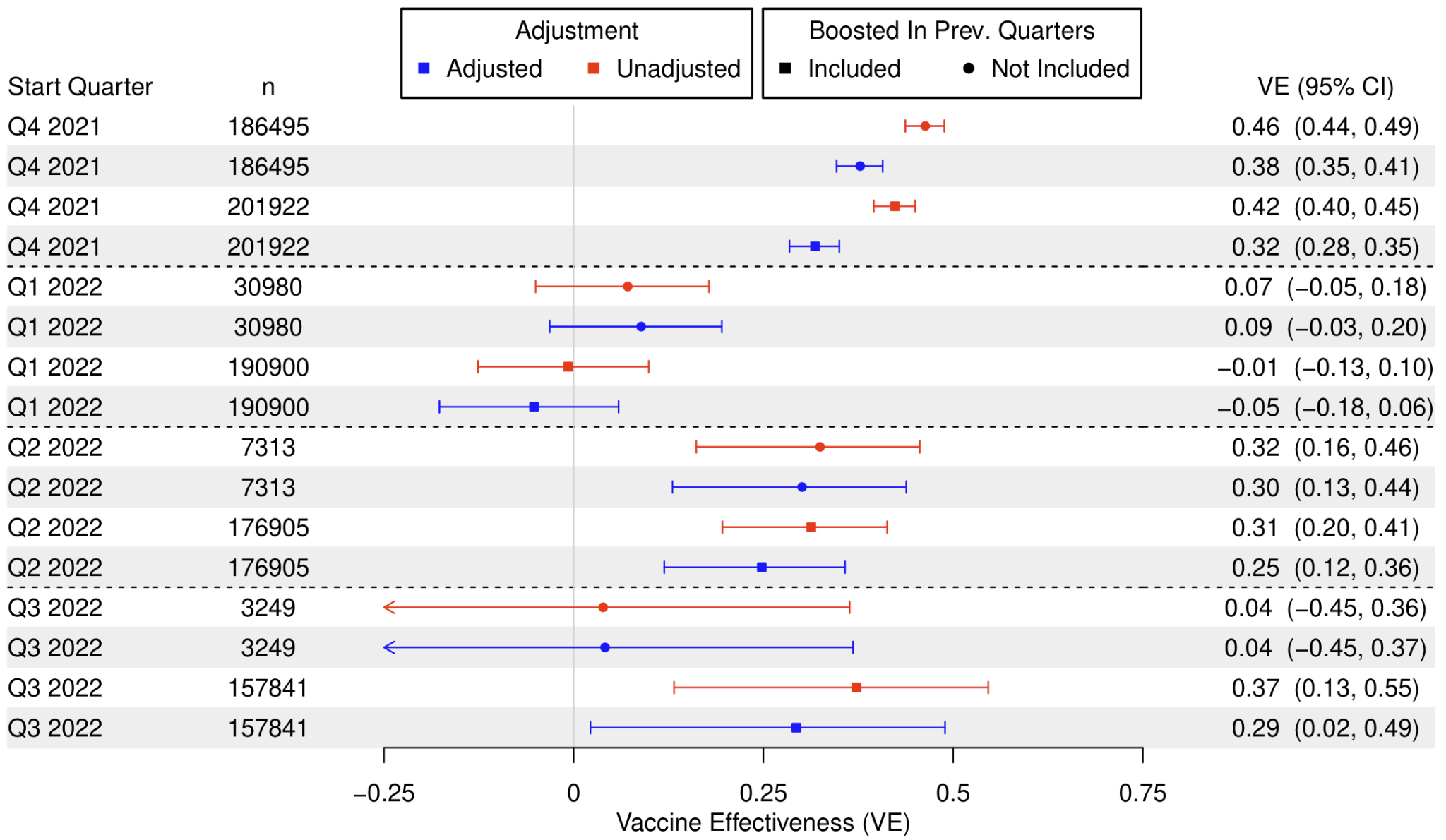

**Figure A16**: Cohort boosted (3 or more doses) vs full vaccination/second dose VE against infection, with various study start (index) dates and with an end quarter of Q4 2022.

Abbreviations: Boost in prior Q, if design includes in the boosted group only those boosted in the start quarter (N) or those boosted before or during the start quarter (Y).

**Cohort Severe Disease – One or More Boosters vs. Primary Series**

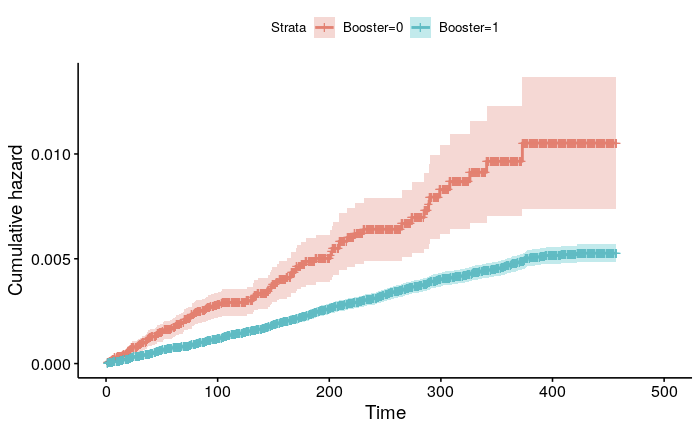

**Figure A17:** Kaplan-Meier curve for cohort boosted (3 or more doses) vs full vaccination/second dose VE against severe disease during a study period of October 1, 2021 to December 31, 2022.

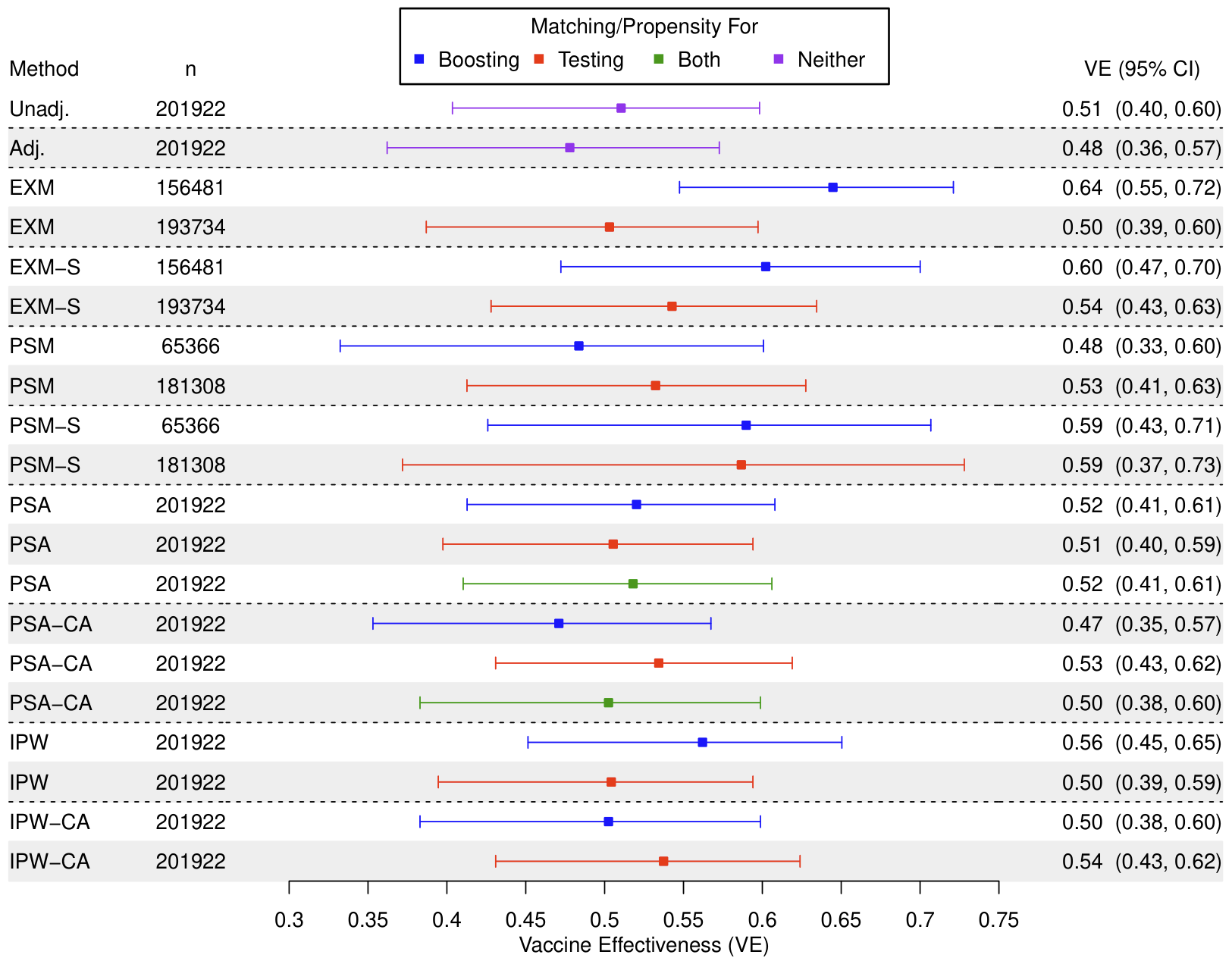

**Figure A18**: Cohort boosted (3 or more doses) vs full vaccination/second dose VE against severe disease, with various matching or propensity score designs during a study period of October 1, 2021 to December 31, 2022.

Abbreviations: Unadj., unadjusted; Adj., adjusted for covariates; EXM, exact matching; EXM-S, exact matching with matching strata stratification in Cox regression; PSM, propensity score matching with a 0.2 caliper; PSM-S, propensity score matching with a 0.2 caliper and matching strata stratification in Cox regression; PSA; adjustment for propensity score; PSA-CA, adjustment for propensity score and for covariates; IPW, inverse probability weighting of propensity score; IPW-CA, inverse probability weighting of propensity score also adjusting for covariates.

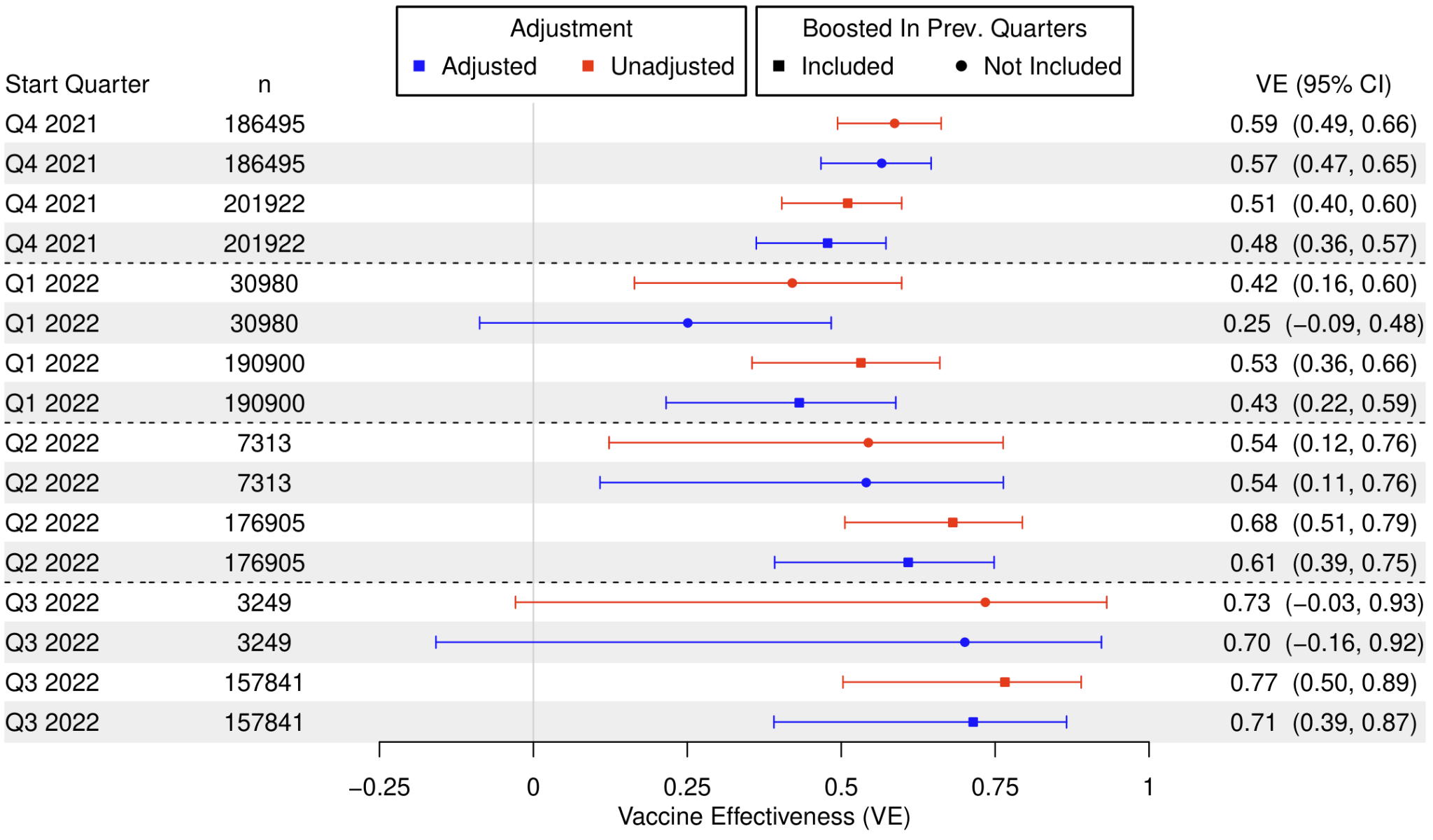

**Figure A19**: Cohort boosted (3 or more doses) vs full vaccination/second dose VE against severe disease, with various study start (index) dates and choices of control group and with an end quarter of Q4 2022.

Abbreviations: Boost in prior Q, if design includes in the second boosted group only those boosted in the start quarter (N) or those boosted before or during the start quarter (Y).

**Test-Negative Infection – One or More Boosters vs. Primary Series**

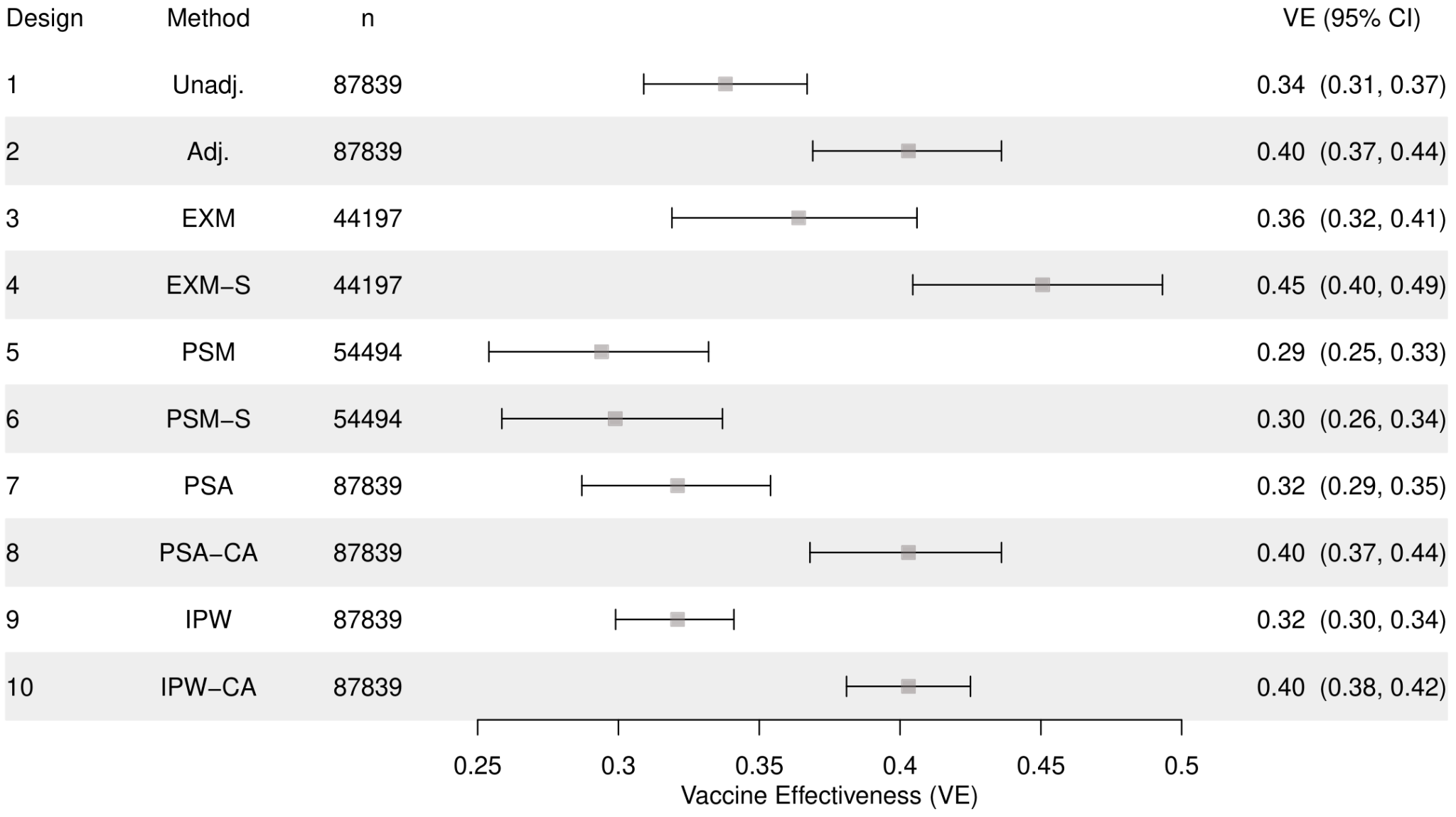

**Figure A20**: Test-negative boosted (3 or more doses) vs full vaccination/second dose VE against infection, with various matching designs during a study period of October 1, 2021 to December 31, 2022.

Abbreviations: Unadj., unadjusted; Adj., adjusted for covariates; EXM, exact matching; EXM-S, exact matching accounting for matching strata in conditional logistic regression; PSM, propensity score matching with a 0.2 caliper; PSM-S, propensity score matching with a 0.2 caliper accounting for matching strata in conditional logistic regression; PSA; adjustment for propensity score; PSA-CA, adjustment for propensity score and for covariates; IPW, inverse probability weighting of propensity score; IPW-CA, inverse probability weighting of propensity score also adjusting for covariates.

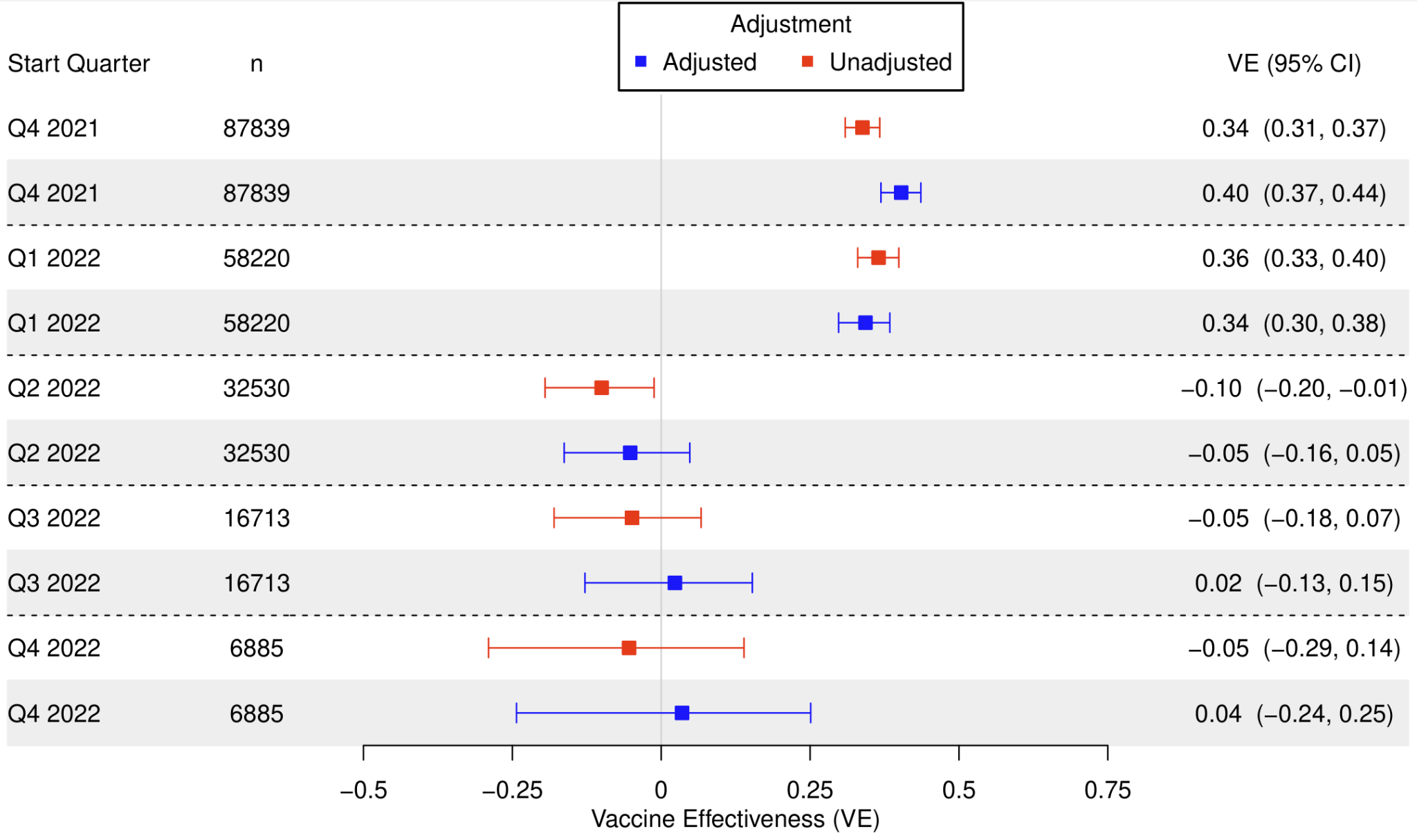

**Figure A21**: Test-negative boosted (3 or more doses) vs full vaccination/second dose VE against infection, with various study start (index) dates and with an end quarter of Q4 2022.

**Test-Negative Severe Disease – One or More Boosters vs. Primary Series**

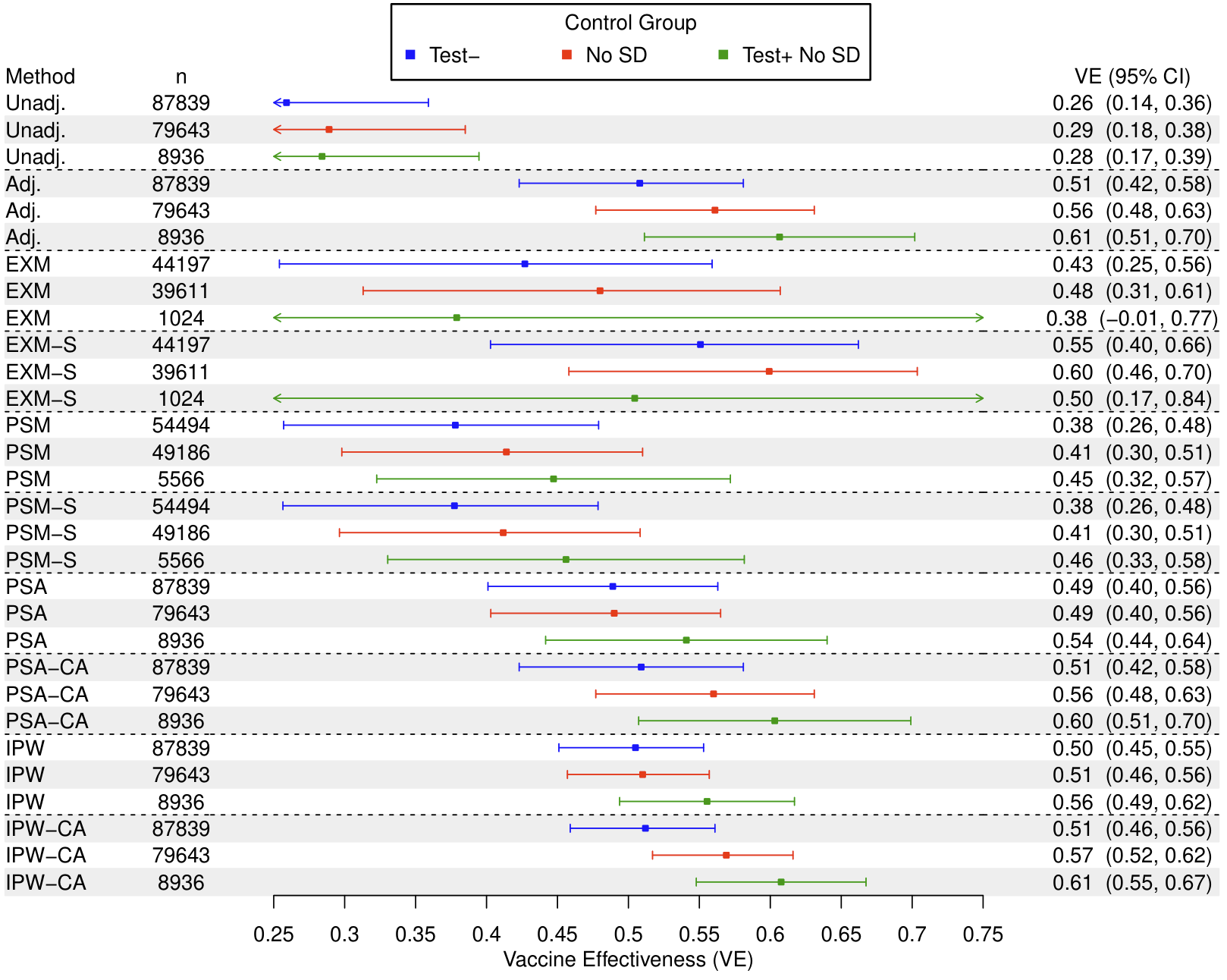

**Figure A22**: Test-negative boosted (3 or more doses) vs full vaccination/second dose VE against severe disease, with various matching or propensity score designs during a study period of October 1, 2021 to December 31, 2022.

Control Groups: (1) Test-, test-negatives; (2) No SD, no severe disease; (3) Test+ No SD, test-positives without severe disease.

Abbreviations: Unadj., unadjusted; Adj., adjusted for covariates; EXM, exact matching; EXM-S, exact matching accounting for matching strata in conditional logistic regression; PSM, propensity score matching with a 0.2 caliper; PSM-S, propensity score matching with a 0.2 caliper accounting for matching strata in conditional logistic regression; PSA; adjustment for propensity score; PSA-CA, adjustment for propensity score and for covariates; IPW, inverse probability weighting of propensity score; IPW-CA, inverse probability weighting of propensity score also adjusting for covariates.

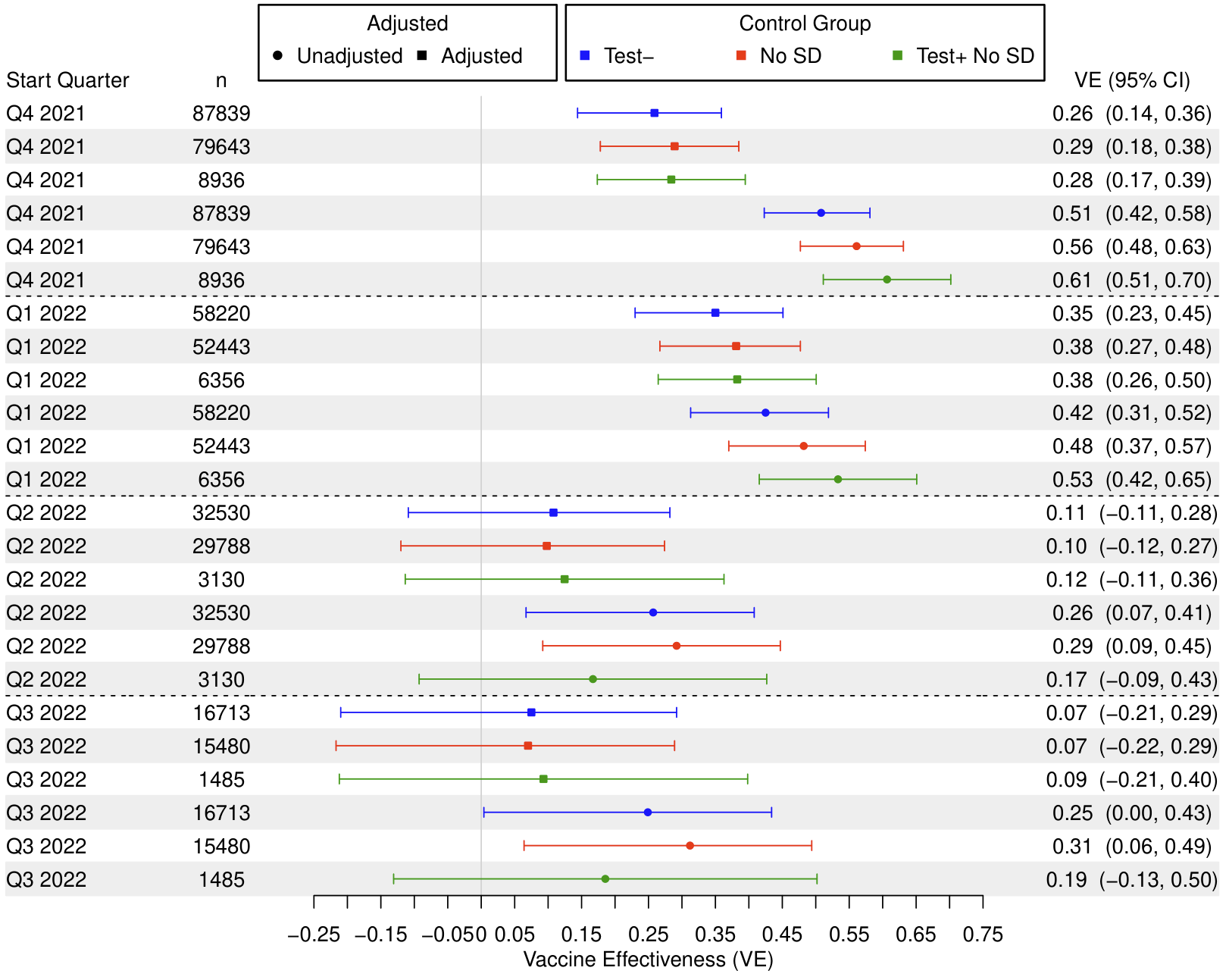

**Figure A23**: Test-negative boosted (3 or more doses) vs full vaccination/second dose VE against severe disease, with various study start (index) dates and choices of control group and with an end quarter of Q4 2022.

Control Groups: (1) Test-, test-negatives; (2) No SD, no severe disease; (3) Test+ No SD, test-positives without severe disease.

**Two or More Boosters vs. One Booster**

**Cohort Infection – Two or More Boosters vs. One Booster**

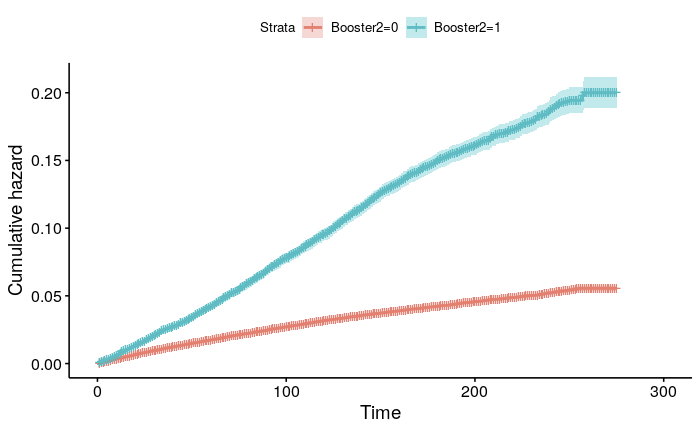

**Figure A24:** Kaplan-Meier curve for cohort double boosted or more (4 or more doses) vs single boosted (3 doses) VE against infection.

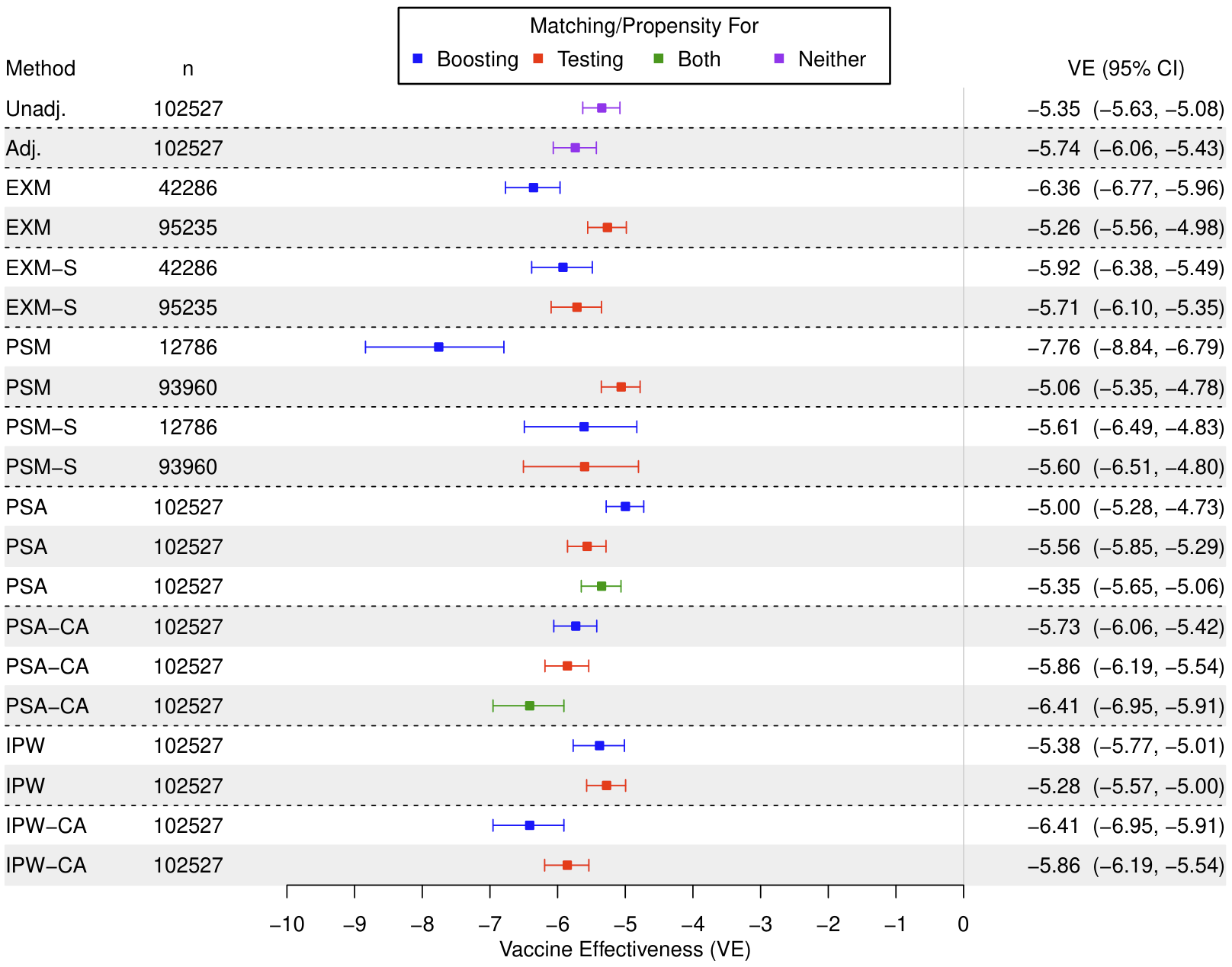

**Figure A25**: Cohort double boosted or more (4 or more doses) vs single boosted (3 doses) VE against infection, with various matching designs during a study period of April 1, 2022 to December 31, 2022.

Abbreviations: Unadj., unadjusted; Adj., adjusted for covariates; EXM, exact matching; EXM-S, exact matching with matching strata stratification in Cox regression; PSM, propensity score matching with a 0.2 caliper; PSM-S, propensity score matching with a 0.2 caliper and matching strata stratification in Cox regression; PSA; adjustment for propensity score; PSA-CA, adjustment for propensity score and for covariates; IPW, inverse probability weighting of propensity score; IPW-CA, inverse probability weighting of propensity score also adjusting for covariates.

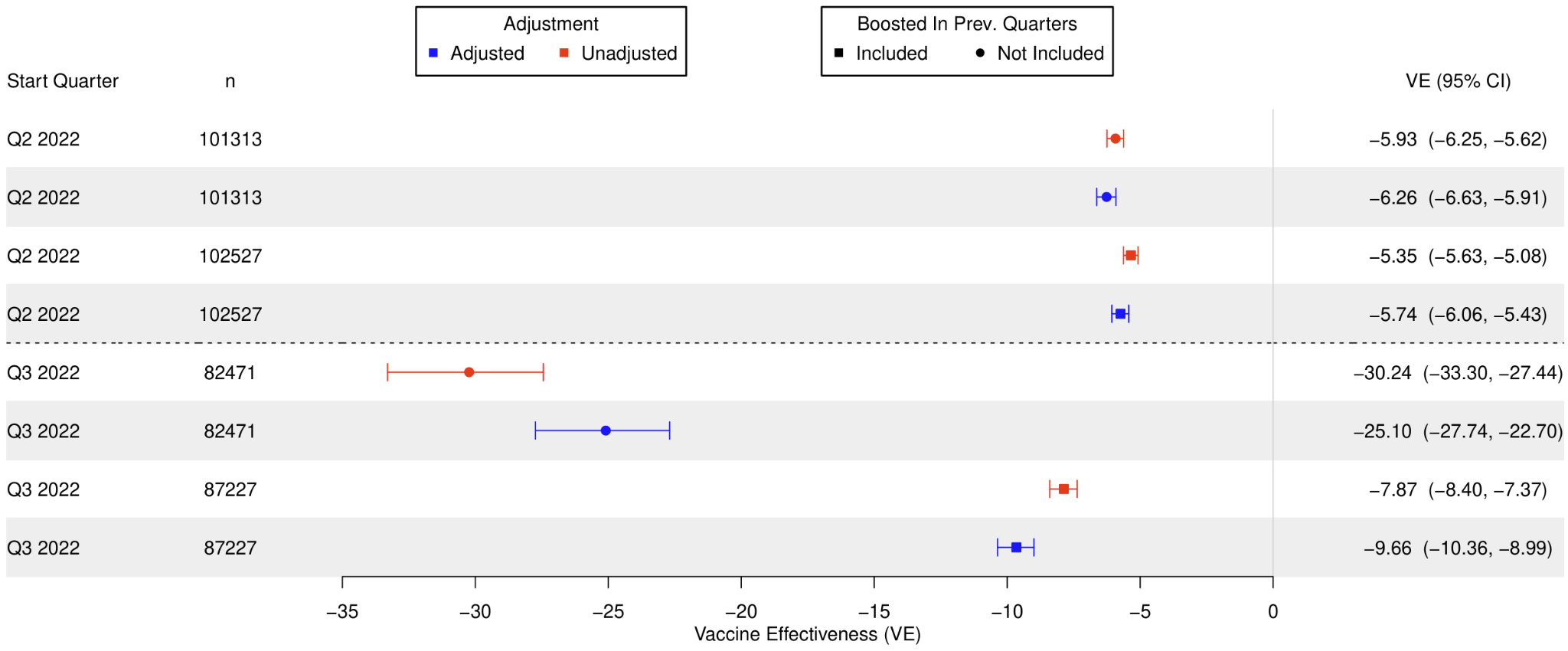

**Figure A26**: Cohort double boosted or more (4 or more doses) vs single boosted (3 doses) VE against infection, with various study start (index) dates and with an end quarter of Q4 2022.

Abbreviations: Boost in prior Q, if design includes in the second boosted group only those boosted in the start quarter (N) or those boosted before or during the start quarter (Y).

**Cohort Severe Disease – Two or More Boosters vs. One Booster**

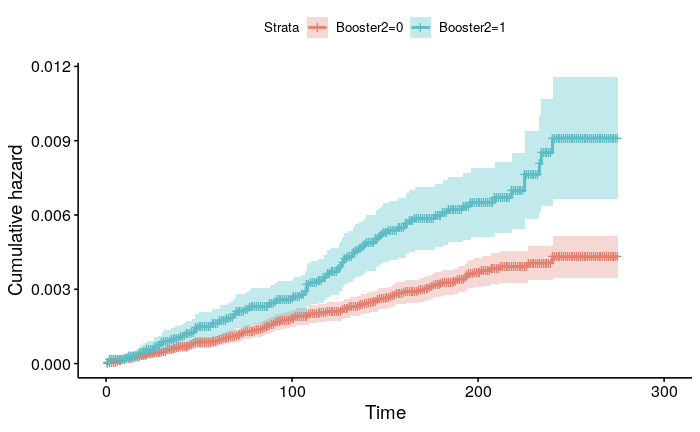

**Figure A27:** Kaplan-Meier curves for cohort double boosted (4 or more doses) vs single boosted (3 doses) VE against severe disease during a study period of April 1, 2022 to December 31, 2022.

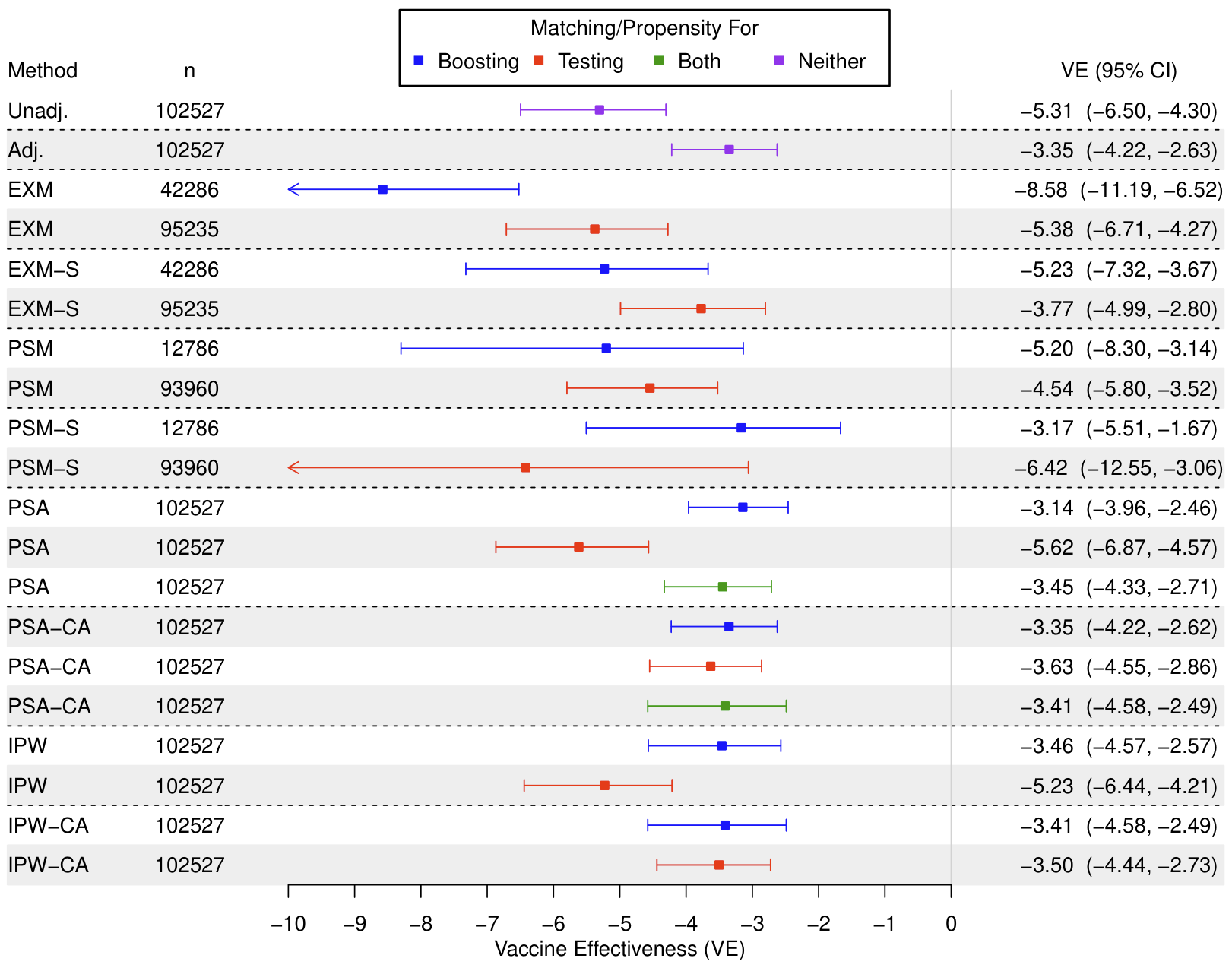

**Figure A28**: Cohort double boosted (4 or more doses) vs single boosted (3 doses) VE against severe disease, with various matching or propensity score designs during a study period of April 1, 2022 to December 31, 2022.

Abbreviations: Unadj., unadjusted; Adj., adjusted for covariates; EXM, exact matching; EXM-S, exact matching with matching strata stratification in Cox regression; PSM, propensity score matching with a 0.2 caliper; PSM-S, propensity score matching with a 0.2 caliper and matching strata stratification in Cox regression; PSA; adjustment for propensity score; PSA-CA, adjustment for propensity score and for covariates; IPW, inverse probability weighting of propensity score; IPW-CA, inverse probability weighting of propensity score also adjusting for covariates.

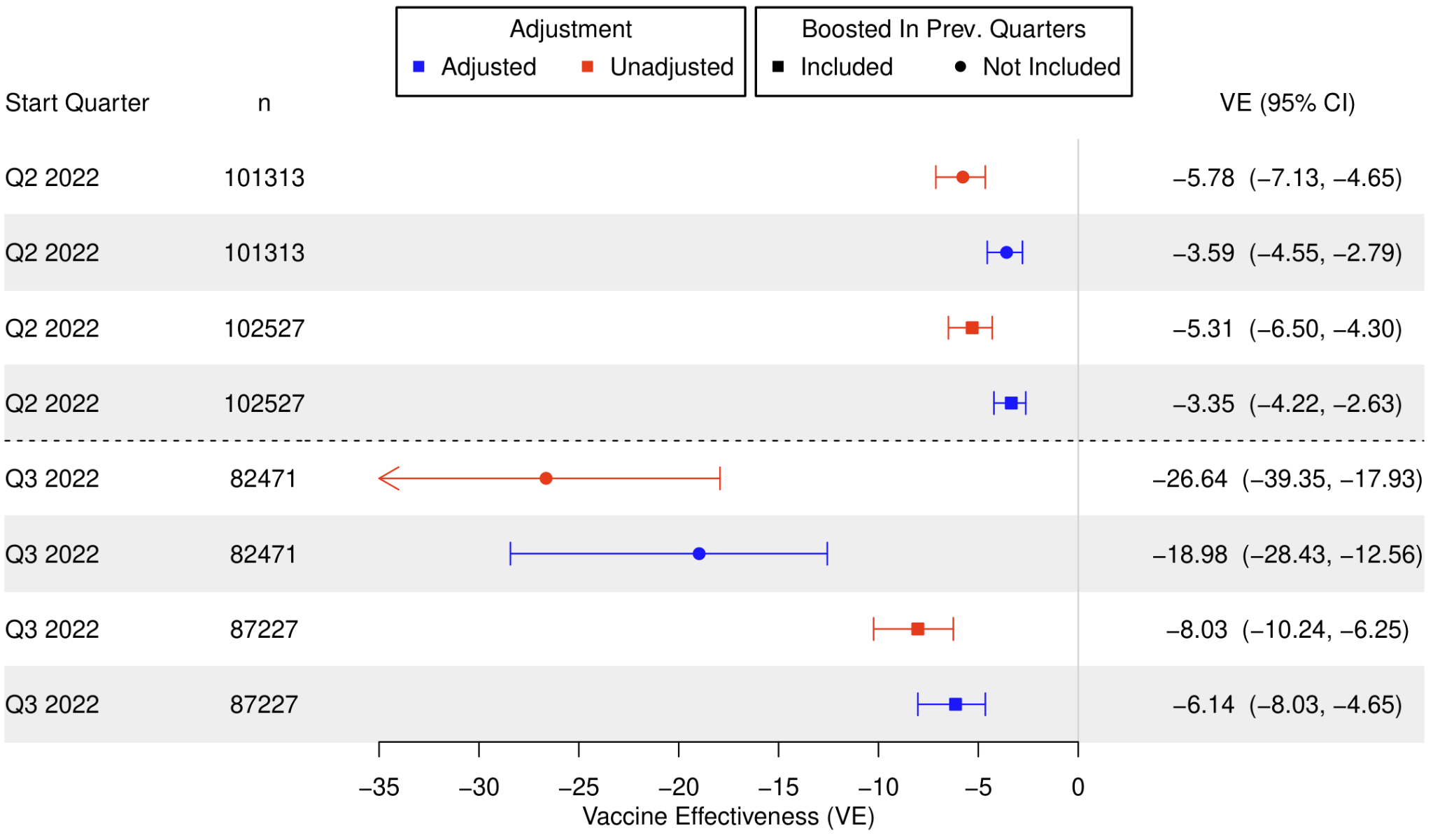

**Figure A29**: Cohort double boosted (4 or more doses) vs single boosted (3 doses) VE against severe disease, with various study start (index) dates and choices of control group and with an end quarter of Q4 2022.

Abbreviations: Boost in prior Q, if design includes in the second boosted group only those boosted in the start quarter (N) or those boosted before or during the start quarter (Y).

**Test-Negative Infection – Two or More Boosters vs. One Booster**

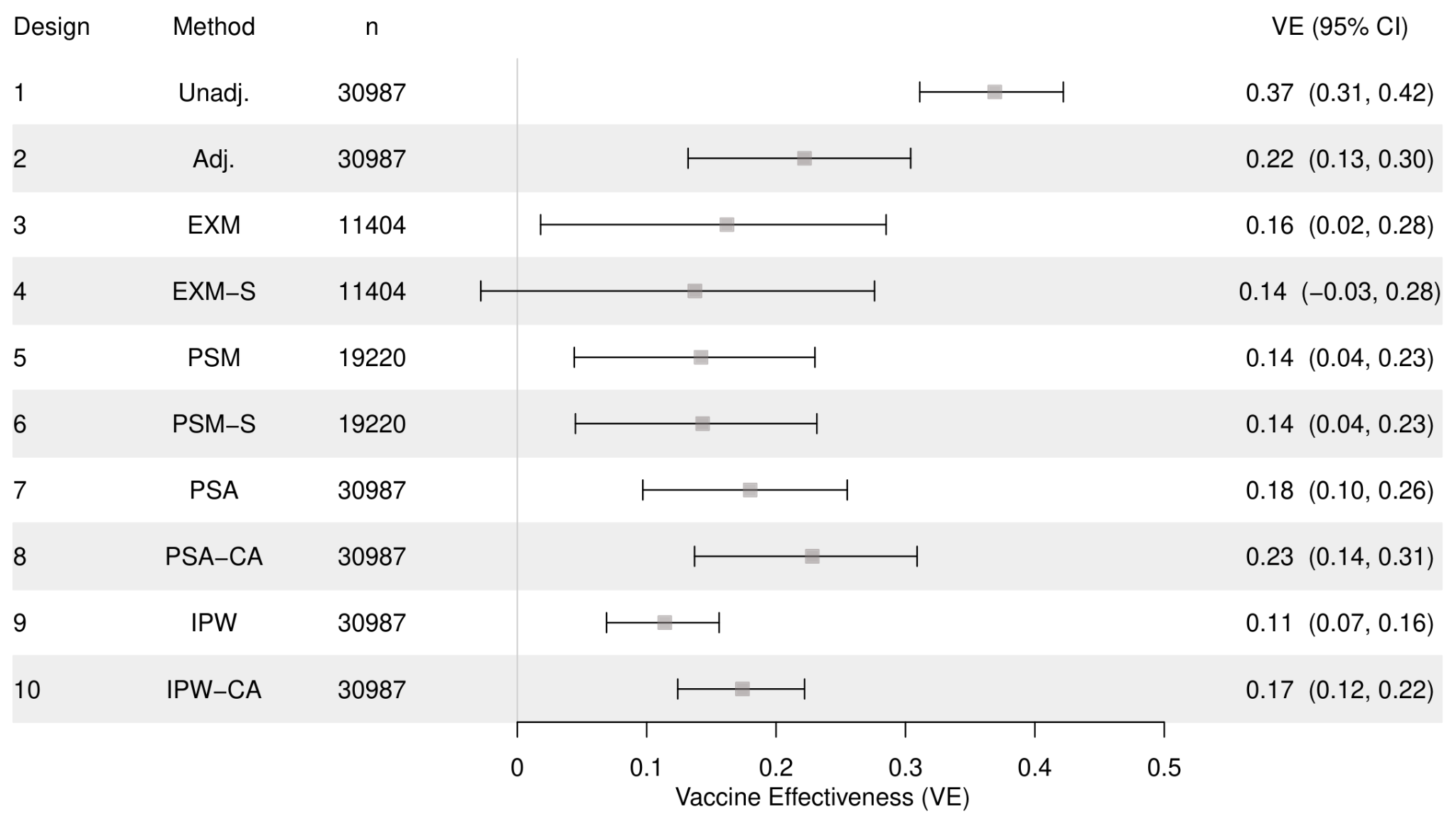

**Figure A30:** Test-negative double boosted or more (4 or more doses) vs single boosted (3 doses) VE against infection, with various matching designs during a study period of April 1, 2022 to December 31, 2022.

Abbreviations: Unadj., unadjusted; Adj., adjusted for covariates; EXM, exact matching; EXM-S, exact matching accounting for matching strata in conditional logistic regression; PSM, propensity score matching with a 0.2 caliper; PSM-S, propensity score matching with a 0.2 caliper accounting for matching strata in conditional logistic regression; PSA; adjustment for propensity score; PSA-CA, adjustment for propensity score and for covariates; IPW, inverse probability weighting of propensity score; IPW-CA, inverse probability weighting of propensity score also adjusting for covariates.

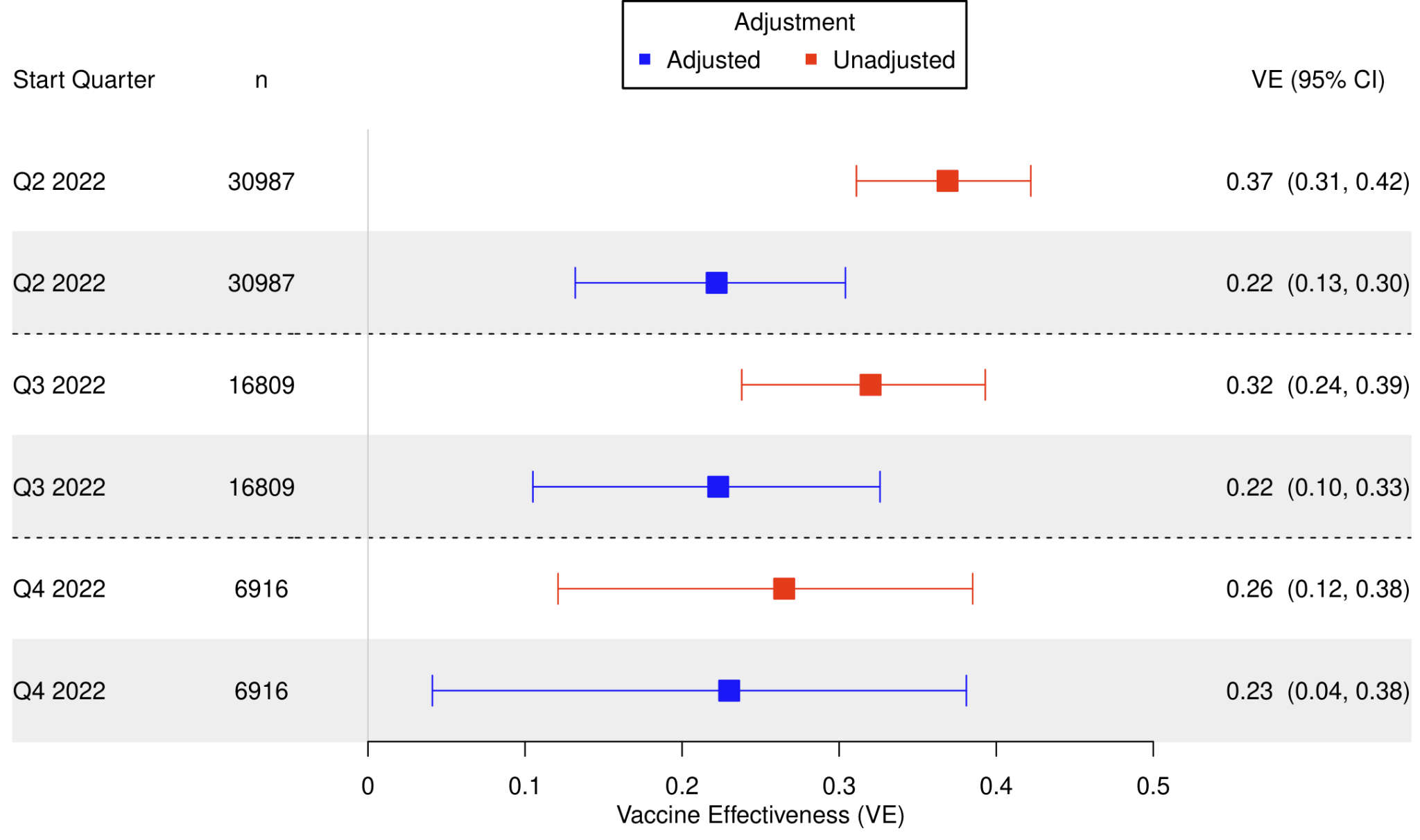

**Figure A31**: Test-negative double boosted or more (4 or more doses) vs single boosted (3 doses) VE against infection, with various study start (index) dates and with an end quarter of Q4 2022.

**Test-Negative Severe Disease – Two or More Boosters vs. One Booster**

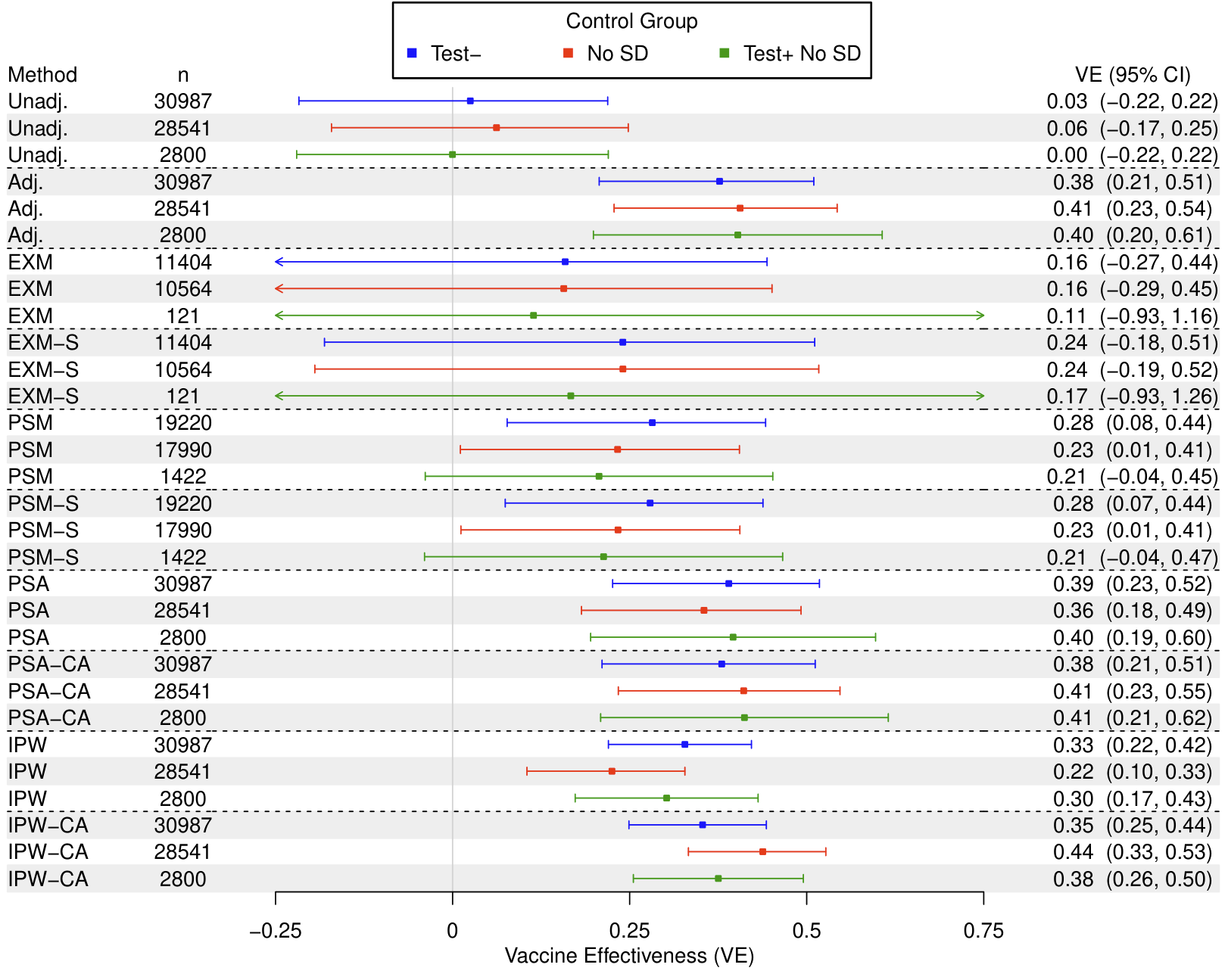

**Figure A32**: Test-negative double boosted (4 or more doses) vs single boosted (3 doses) VE against severe disease, with various matching or propensity score designs during a study period of April 1, 2022 to December 31, 2022.

Control Groups: (1) Test-, test-negatives; (2) No SD, no severe disease; (3) Test+ No SD, test-positives without severe disease.

Abbreviations: Unadj., unadjusted; Adj., adjusted for covariates; EXM, exact matching; EXM-S, exact matching accounting for matching strata in conditional logistic regression; PSM, propensity score matching with a 0.2 caliper; PSM-S, propensity score matching with a 0.2 caliper accounting for matching strata in conditional logistic regression; PSA; adjustment for propensity score; PSA-CA, adjustment for propensity score and for covariates; IPW, inverse probability weighting of propensity score; IPW-CA, inverse probability weighting of propensity score also adjusting for covariates.

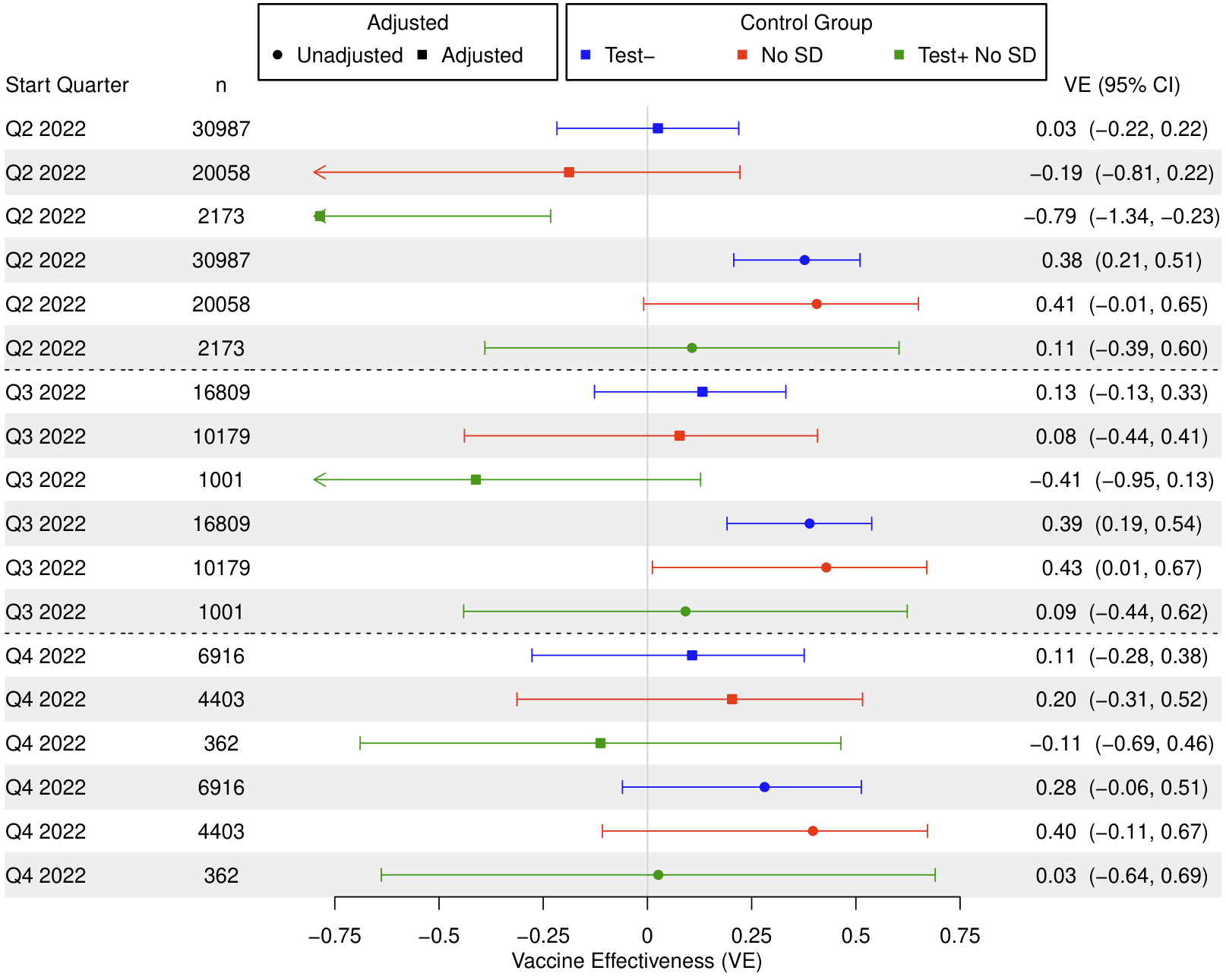

**Figure A33**: Double boosted (4 or more doses) vs single boosted (3 doses) VE against severe disease, with various study start (index) dates and choices of control group and with an end quarter of Q4 2022.

Control Groups: (1) Test-, test-negatives; (2) No SD, no severe disease; (3) Test+ No SD, test-positives without severe disease.
